# Do the effects of interventions aimed at the prevention of childhood obesity reduce inequities? A re-analysis of randomized trial data from two Cochrane reviews

**DOI:** 10.1101/2024.06.10.24308372

**Authors:** Jennifer C Palmer, Annabel L Davies, Francesca Spiga, Berit L Heitmann, Russell Jago, Carolyn D Summerbell, Julian PT Higgins, Inequity in Obesity Prevention Trialists Collaborative Group

## Abstract

**Background:** Public health attempts to prevent obesity in children and young people should aim to minimize health inequalities. We aimed to assess whether there were differences in the effectiveness of the interventions included in two Cochrane reviews according to the eight PROGRESS inequity factors.

**Methods:** We collected data on change in BMI (standardized or unstandardized), subgrouped by baseline measures of PROGRESS factors, for intervention and control groups, from trial authors. We calculated the intervention effect per subgroup (mean difference), then contrasted these to estimate interactions between intervention and the baseline factors. We combined interaction estimates for each factor across trials using standard procedures for meta-analysis.

**Findings:** Our analysis of interventions from 81 trials found no substantial differences in effectiveness for different subgroups in most scenarios. However, in the younger age group (5-11 years), the effect of interventions on standardized BMI appeared to be higher in boys.

**Interpretation:** The Cochrane reviews found that interventions promoting physical activity (only) have a beneficial effect on BMI (compared with a control group) for children and young people aged 5-18 years, as well as interventions promoting physical activity alongside healthy eating for 5-11 year olds. Although these beneficial effects were small, when delivered at scale, they may have the potential to contribute meaningfully to reducing the prevalence of childhood obesity. Our findings suggest that those responsible for public health can promote these beneficial interventions without major concerns about increasing inequalities. Because many of the interventions studied, including school-based interventions, provide building blocks of ‘whole systems approaches’, the findings are relevant to policy and practice.

**Funding:** National Institute for Health and Care Research (NIHR).

**Research in context:** *Evidence before this study:* A core principle of any public health guidance is to minimize health inequalities. Two previous studies of the effects of interventions aiming to prevent obesity in children and young people, by promoting physical activity or a healthy diet, found that such interventions do not increase health inequalities. However, these studies used secondary data published in trial reports, limiting the data available for analysis. Two recent Cochrane systematic reviews and meta-analyses of over 200 randomized trials of interventions to prevent obesity in children and young people, found, *on average*, small beneficial effects of physical activity interventions in 5-18 year olds on (standardized and unstandardized) BMI and of combined physical activity and dietary interventions in 5-11 year olds. A previous modelling study found that small beneficial benefits such as these, when delivered at scale, have the potential to contribute meaningfully to reducing the prevalence of childhood obesity. However, average effects may mask differential effects on health equity. Our objective was to collect primary trial results (not previously reported) to examine whether effects of interventions vary according to factors related to inequity as represented by the PROGRESS acronym: place, race/ethnicity, occupation, gender/sex, religion, education, socio-economic status, and social capital.

*Added value of this study:* To the best of our knowledge, this is the first large-scale meta-analysis to assess the impact of interventions to prevent obesity in children and young people on health equity using primary data from randomized trials. Data from 81 trials were included, collected directly from the trialists as aggregate data by intervention and by subgroup, and combined in meta-analyses. We found no substantial impact of the interventions on inequalities, although in the younger age group (5-11 years), the effect of interventions (n=45) on standardized BMI was greater in boys.

*Implications of all the available evidence:* Those responsible for public health can be confident in promoting the types of interventions included in this meta-analysis to prevent obesity in children and young people (5-18 years), knowing they are unlikely to increase inequalities. One exception was that interventions for younger children may benefit from being equally engaging and enjoyable for females and males. We regard ‘whole systems approaches’ to comprise separate interventions (components) interconnected via a programme theory and logic model, including the types of interventions included in this meta-analysis. As such, our findings are relevant to those providing guidance on a whole systems approach to reducing the prevalence of obesity in children and young people alongside promoting health equity.

## Introduction

Population levels of overweight and obesity in childhood are a significant global challenge.^1^ From 1990 to 2022, age-standardized prevalence of obesity increased in girls in 186 countries and in boys in 195 countries. In most countries, obesity more than doubled.^2^ Children and adolescents living with obesity are more likely to experience reduced health-related quality of life and, for adolescents, comorbidities including type 2 diabetes mellitus, fatty liver disease and poor mental health.^3^ The primary prevention of childhood obesity is therefore important not only to promote good long term physical and mental health but also to help children realise their full life-time potential.^4^

Inequalities in the prevalence of childhood obesity are widening in the UK and other high-income countries.^5^ There are unfair differences, or inequities, between population subgroups categorized by shared characteristics. The strongest evidence is for socioeconomic status (SES). In high-income countries, higher rates of obesity are present in those with lower SES,^6,7^ whereas the opposite relationship is observed in most middle-income countries,^7^ and in low-income countries the relationship varies.^8^ Prevalence of childhood obesity is also linked to place of residence,^9^ race and ethnicity^10^ and gender.^11^ There is less evidence for a link with religion or social capital in children, although a connection has been observed in adults.^12,13^

It is important that attempts to prevent obesity in children recognize these unfair differences. Population-level impacts of interventions can hide differences in effects between subgroups. Even if benefits are seen in all subgroups of the population, interventions may lead to greater inequalities.^14^ Interventions are needed that lead to population-level increases in health and wellbeing while also reducing inequities. It is therefore important to understand whether the effectiveness of interventions varies by inequity factors.^15,16^

Characteristics of intervention content, delivery and implementation have been suggested as reducing inequalities or driving intervention-generated inequalities. Targeted (rather than universal) interventions are proposed as a preferred way to address health disparities.^17^ There is evidence that interventions operating within a higher domain (upstream) of the socio-ecological model^18^ and those on the higher steps (requiring less individual agency) of the Nuffield intervention ladder^19^ are more likely to minimize intervention-generated inequalities in SES.^20,21^

Two recent Cochrane systematic reviews and meta-analyses identified over 200 randomized trials of interventions to prevent obesity in children and young people aged 5-11 and 12-18 years, respectively.^22,23^ The findings suggested that a range of physical activity interventions, alone or in combination with dietary interventions, can have a modest beneficial effect, on average, on preventing obesity.

This paper describes a re-analysis of this evidence base to examine whether there were differences in the effectiveness of these interventions according to the eight inequity factors identified in the PROGRESS framework: place (of residence), race/ethnicity, occupation (of parents), gender/sex, religion, education (of parents), socio-economic status and social capital.^15^ Examination of the published trial reports revealed that many of these PROGRESS inequity factors had been collected at baseline, although subgroup analyses were seldom reported. Therefore, we collected subgroup data from as many trials as possible and used meta-analysis to examine differences with higher statistical power than would be possible in any individual trial. We focus on body mass index (BMI, both standardized by age and sex and unstandardized) as this is the most widely measured outcome in obesity prevention trials.

## Methods

### Selection of included trials

We identified trials through the two recent Cochrane systematic reviews on interventions for preventing obesity in children aged 5 to 18 years.^24,25^ In brief, these reviews sought studies that: (i) were individually- or cluster-randomized trials; (ii) recruited children with a mean age between 5 and 18; (iii) measured BMI or standardized BMI z-score (zBMI) assessed at baseline and at least 12 weeks after baseline; (iv) examined an intervention whose main aim was to change at least one from: diet, physical activity, sedentary behaviour, sleep, play or structured exercise to help prevent childhood obesity; and (v) published primary results in 1990 or later. We sought only comparisons of active interventions against a control group.

### Data collection

We extracted subgroup outcome data from publications where they were reported. Where relevant subgroup data had not been reported, the corresponding authors of the trials were emailed to request these data. The emails were tailored to each recipient to include details of the main published report for the trial in question and any information we had already extracted from the trial reports about the impact of baseline PROGRESS factors. The email included a link to the protocol for our investigation^26^ and access to a data collection table into which outcome data suitable for our analyses could be entered. Emails were followed up selectively, with reminders written to authors of trials that (i) we knew had collected data on PROGRESS characteristics, (ii) were published within the last 15 years, and (iii) included at least 200 participants.

We sought data subgrouped by baseline measures of PROGRESS factors. For each PROGRESS factor, trialists were asked to divide the trial participants into exactly two subgroups as described in Table 1. Since our primary interest was in the direction of differences between subgroups, the precise cut off for dichotomization was not critical. Our preference was for inequity factors to be measured and dichotomized at the individual child level, but we accepted group-level categorizations for each child (e.g. at school-level) if that is how the factor was measured. Data were also requested about baseline weight status and we will report the results of our investigation into baseline weight status elsewhere.

**Table 1:**
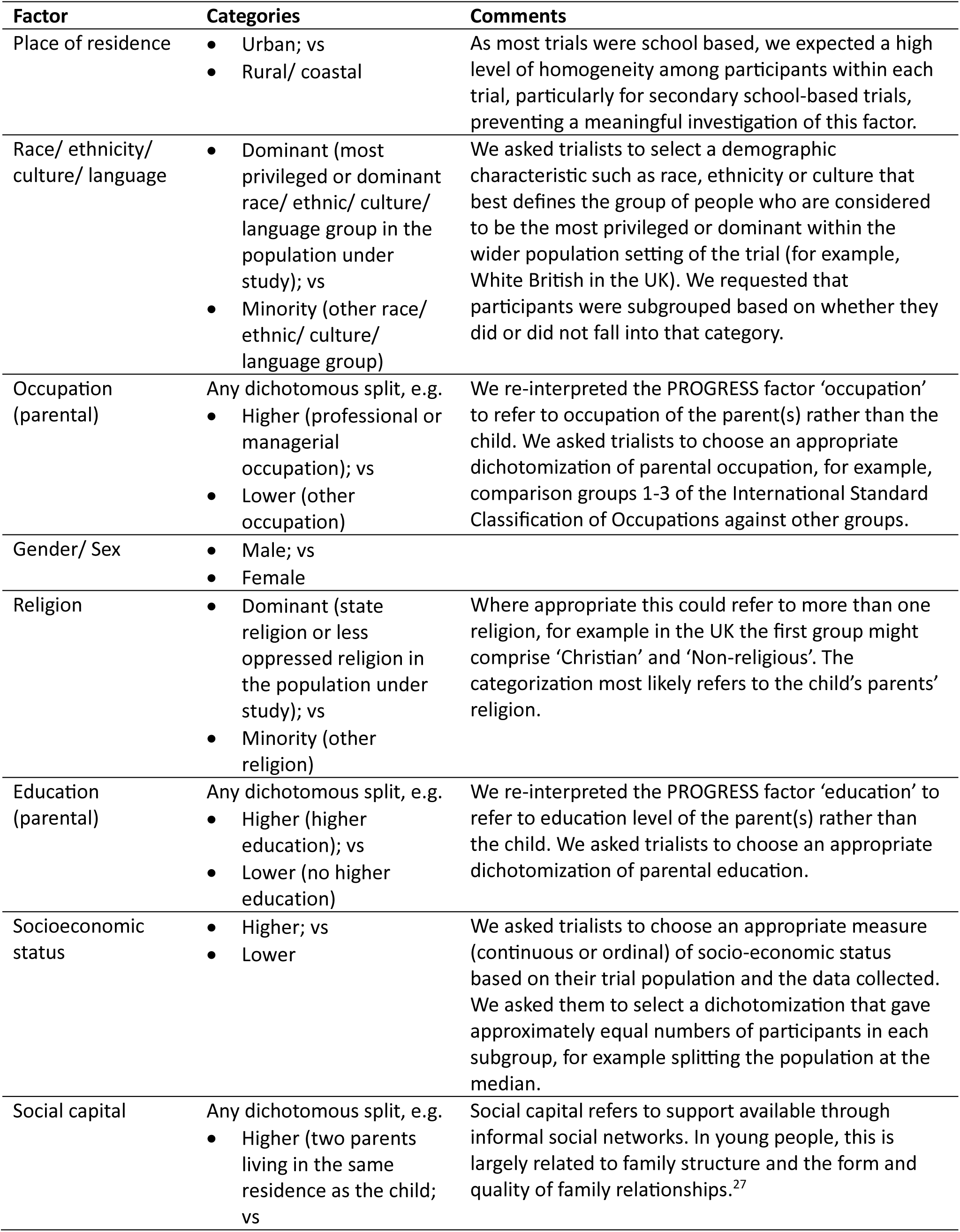
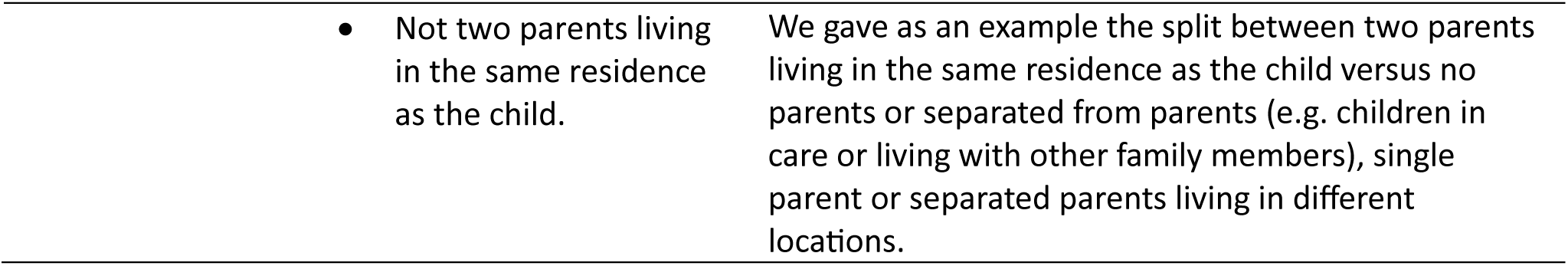
Dichotomization of baseline PROGRESS factors into subgroups.

| Factor | Categories | Comments |
| --- | --- | --- |
| Place of residence | • Urban; vs<br>• Rural/ coastal | As most trials were school based, we expected a high level of homogeneity among participants within each trial, particularly for secondary school-based trials, preventing a meaningful investigation of this factor. |
| Race/ ethnicity/ culture/ language | • Dominant (most privileged or dominant race/ ethnic/ culture/ language group in the population under study); vs<br>• Minority (other race/ ethnic/ culture/ language group) | We asked trialists to select a demographic characteristic such as race, ethnicity or culture that best defines the group of people who are considered to be the most privileged or dominant within the wider population setting of the trial (for example, White British in the UK). We requested that participants were subgrouped based on whether they did or did not fall into that category. |
| Occupation (parental) | Any dichotomous split, e.g.<br>• Higher (professional or managerial occupation); vs<br>• Lower (other occupation) | We re-interpreted the PROGRESS factor 'occupation' to refer to occupation of the parent(s) rather than the child. We asked trialists to choose an appropriate dichotomization of parental occupation, for example, comparison groups 1-3 of the International Standard Classification of Occupations against other groups. |
| Gender/ Sex | • Male; vs<br>• Female |  |
| Religion | • Dominant (state religion or less oppressed religion in the population under study); vs<br>• Minority (other religion) | Where appropriate this could refer to more than one religion, for example in the UK the first group might comprise 'Christian' and 'Non-religious'. The categorization most likely refers to the child's parents' religion. |
| Education (parental) | Any dichotomous split, e.g.<br>• Higher (higher education); vs<br>• Lower (no higher education) | We re-interpreted the PROGRESS factor 'education' to refer to education level of the parent(s) rather than the child. We asked trialists to choose an appropriate dichotomization of parental education. |
| Socioeconomic status | • Higher; vs<br>• Lower | We asked trialists to choose an appropriate measure (continuous or ordinal) of socio-economic status based on their trial population and the data collected. We asked them to select a dichotomization that gave approximately equal numbers of participants in each subgroup, for example splitting the population at the median. |
| Social capital | Any dichotomous split, e.g.<br>• Higher (two parents living in the same residence as the child; vs | Social capital refers to support available through informal social networks. In young people, this is largely related to family structure and the form and quality of family relationships.<sup>27</sup> |

For each subgroup, raw means and standard deviations (SDs) were collected from each intervention arm of each trial (unadjusted for clustering or other variables). In order of preference, the data collection form asked for (i) mean change from baseline, (ii) baseline and follow-up means with the corresponding correlation coefficient (where available), or (iii) follow-up means. It also requested information about clustering (cluster sizes and intraclass correlation coefficients (ICCs)) so that adjustments for clustering could be made in the analyses. Trialists were asked to provide both zBMI and BMI data where available; zBMI was our preferred outcome for the analysis although many trials had collected only BMI. For trials that measured outcomes at multiple follow-up times, data were requested only for the follow-up time closest to 12 months.

We additionally coded individual trials for characteristics shown to be associated with reducing inequalities or driving intervention-generated inequalities: whether the intervention was targeted or universal; the domain of the socio-ecological model it addresses;^18^ the step on the Nuffield intervention ladder it addresses;^19^ whether it included an explicit component aiming to change the structural environment of the child; and the degree of public engagement and involvement in its development.

### Assessment of risk of bias

The Cochrane review had used the RoB 2 tool^28^ to assess risk of bias in each result. Because the biases addressed by RoB 2 may be expected to apply similarly to each subgroup, we would expect many of them to cancel out in our comparisons of subgroups. We therefore supplemented the RoB 2 results with two additional assessments focussing on potential biases in the comparison of two subgroups (i) completeness of data and the extent to which sought results were available (i.e. bias due to missing subgroup data that were extracted from the main analysis papers or provided by trialists) and (ii) classification of participants into subgroups according to thresholds determined by trialists (bias in selection of the subgroup analysis result). Risk-of-bias assessments were undertaken by researchers at the University of Bristol who were not involved in any of the trials.

### Data analysis

Our main analyses were two-stage meta-analyses. For each baseline factor of interest and each trial, we first calculated the intervention effect per subgroup. These were obtained as mean differences (MD) in zBMI (or BMI) where, depending on the data available, the mean from each intervention group was (i) change from baseline provided by the trialists, (ii) change from baseline calculated from baseline and follow-up means and a correlation coefficient provided by the trialists, (iii) change from baseline calculated from baseline and follow-up means and an imputed correlation coefficient, (iv) follow-up means. Based on observed correlations between baseline and follow-up in trials in our original dataset,^22,23^ we imputed a value of 0.9 for scenario (iii). We then calculated the difference in intervention effects between the two subgroups. This estimates the interaction between the intervention and the factor defining the subgroup. Since the subgroups are independent, an estimate of the variance of the interaction is given by the sum of variances of the subgroup-specific effect estimates.

Next, we combined the interaction estimates across trials using standard meta-analysis procedures.^29^ We performed a random-effects meta-analysis to allow for heterogeneity in the estimated interaction parameters. The null hypothesis for each meta-analysis was that the subgroup covariate has no impact on the intervention effect (i.e. the interaction parameter is zero). Using the summary estimate and its standard error from the meta-analysis, we performed a simple Z test of this null hypothesis. The point estimate from the meta-analysis quantifies the average extent to which the intervention effect is impacted by the covariate. Compared with the test of the null hypothesis, practical interpretation of this result requires stronger assumptions about the similarity of relationships across trials. We used the I^2^ statistic to measure the extent to which results were consistent across trials and the P value (P_het_) from a chi-squared test to examine strength of evidence of heterogeneity in the interactions across trials.^30^ Evidence of heterogeneity indicates that the impact of the factor is importantly different in different contexts.

For cluster-randomized trials, we adjusted the standard error of the mean differences to account for clustering using methods described previously.^31^ Where available, this adjustment made use of the ICCs reported by the trialists. Where these were not provided, we used an imputed value of 0.02 based on ICCs reported in other trials, and we performed sensitivity analyses with ICCs of 0 and 0.04. For multi-arm trials, we combined intervention groups following the methods described in the Cochrane Handbook.^32^

We performed a supplementary analysis undertaking meta-analyses separately for the subsets of trials from high-income countries and low- or middle-income countries (according to the World Bank classification), because the association of some PROGRESS factors with obesity may differ between settings (for example, lower socioeconomic status is associated with more obesity in high income countries but often with less obesity in other countries^7^). We additionally performed subset analyses (that were not specified in the protocol) for gender/sex according to whether the intervention targeted diet, physical activity (physical activity, sedentary behaviour or sleep, play or structured exercise) or a combination of diet and physical activity.

## Results

### Response rates and included trials

We summarize the trial selection process in Figure 1. From the 244 trials included in the two Cochrane reviews, we excluded five that performed only head-to-head comparisons without a control group, leaving 239 eligible trials. From our attempts to obtain subgrouped outcome data from the trialists, a response was received from the corresponding (or senior) author of 138 trials (58% response rate for contact). We obtained subgrouped outcome data eligible for inclusion in our analysis from the authors of 64 trials (27% response rate for data collection). We were able to extract subgrouped outcome data from 20 publications, including three trials for which authors provided additional data. We therefore included 81 trials (34% of eligible) in the analyses presented below.

**Figure 1:**
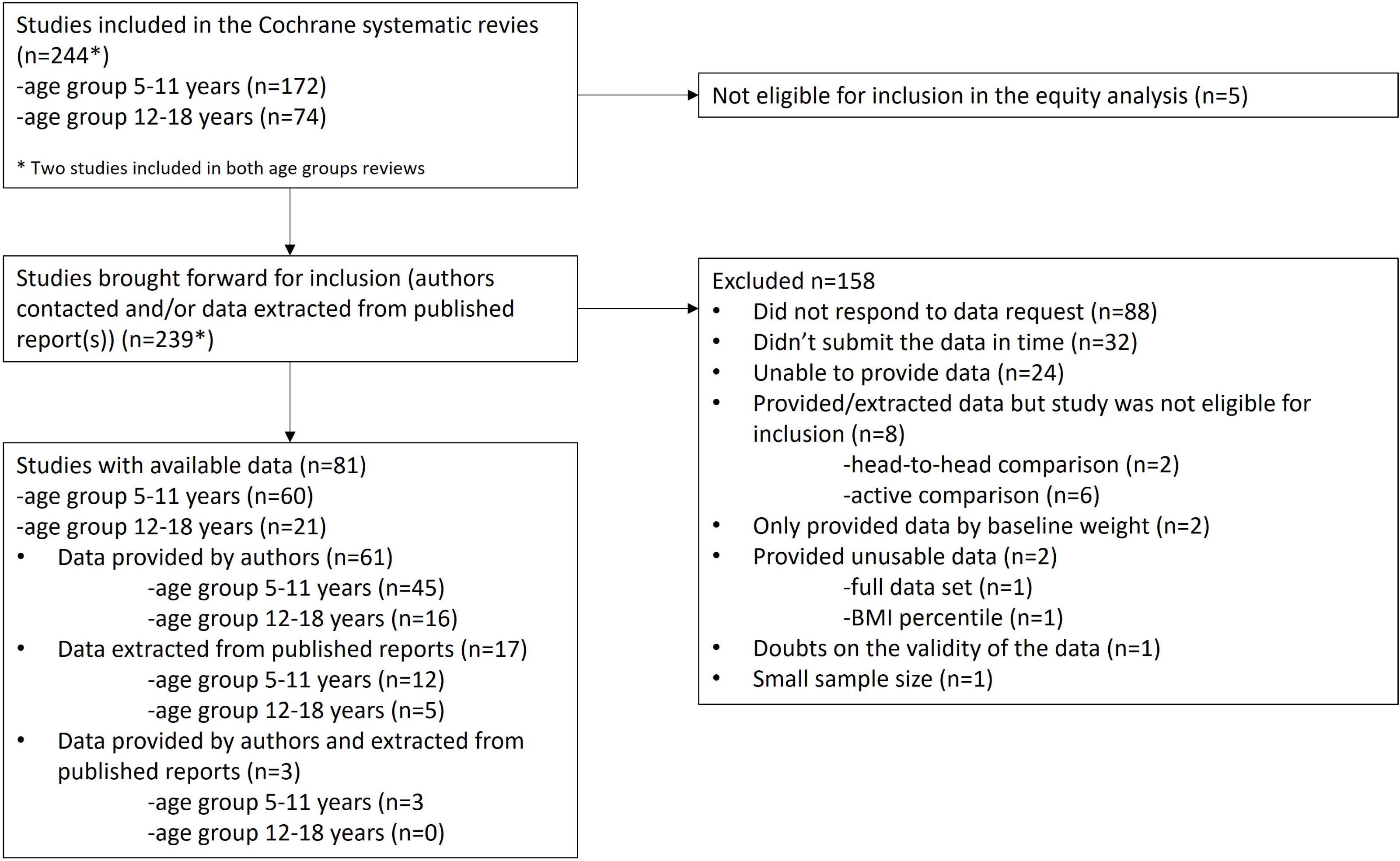
Trial selection process

### Characteristics of included trials

Brief characteristics of the 81 included trials are provided in Supplementary Table 1. Sixty trials (74%) were conducted in children aged 5-11 years and 21 in children and adolescents aged 12-18 years. Sixty-five trials (80%) randomized clusters of children and 16 (20%) randomized individuals. Sixty-seven (83%) were conducted in high-income and 14 (17%) in middle-income countries (11 in upper-middle-income countries and 3 in lower-middle-income countries). Trial locations were the United States for 23 trials (28%), Australia for 15 (19%), China for seven (9%), United Kingdom for six (7%) and the rest of Europe for 15 (19%). The remining 15 locations included one in Egypt and two in Lebanon. In 39 trials (48%), the interventions aimed to change both dietary and activity behaviours, in 30 (37%) physical activity behaviours only and in ten (12%) dietary behaviours only; two trials (3%) had multiple intervention arms and implemented different types of interventions. Most of the trials (62, 77%) implemented the intervention entirely or mainly at school (including after school programs) and six (7%) were implemented entirely or mainly at home. Three (4%) were implemented within a clinical setting and the remaining 10 (12%) in the community or other setting. Three interventions included telehealth.

The coding of individual trials for characteristics associated with inequalities or intervention-generated inequalities is provided in Supplementary Table 2. Most interventions, in total 60 (74%), were universal and 21 (26%) were targeted; some universal interventions were conducted in relatively low-income areas. Nearly all interventions (74/81) operated in the organization or community domains of the socio-ecological model; none in the society or public policy domains and seven in the interpersonal domain. Most interventions (76/81) guided or enabled behaviour change; four included significant restriction (relating to types of foods and beverages available at school) and one provided information via a brief counselling session on healthy weight (at a dental check-up). Around half of the interventions (39/81) involved a change in the school structural environment (for example, changing the catering or school shop layout). Most (56/81) did not report any evidence of public involvement and engagement; 15 reported some degree of consultation and a further ten reported some degree of consultation that included consultation with children. None of the interventions reported using co-production or were user controlled.

Availability of data from individual trials for each PROGRESS characteristic is summarized in Table 2. The mean follow-up time for these outcomes was 10 months (SD 5 months), with the shortest follow-up time being 3 months and the longest 24 months.

**Table 2:**
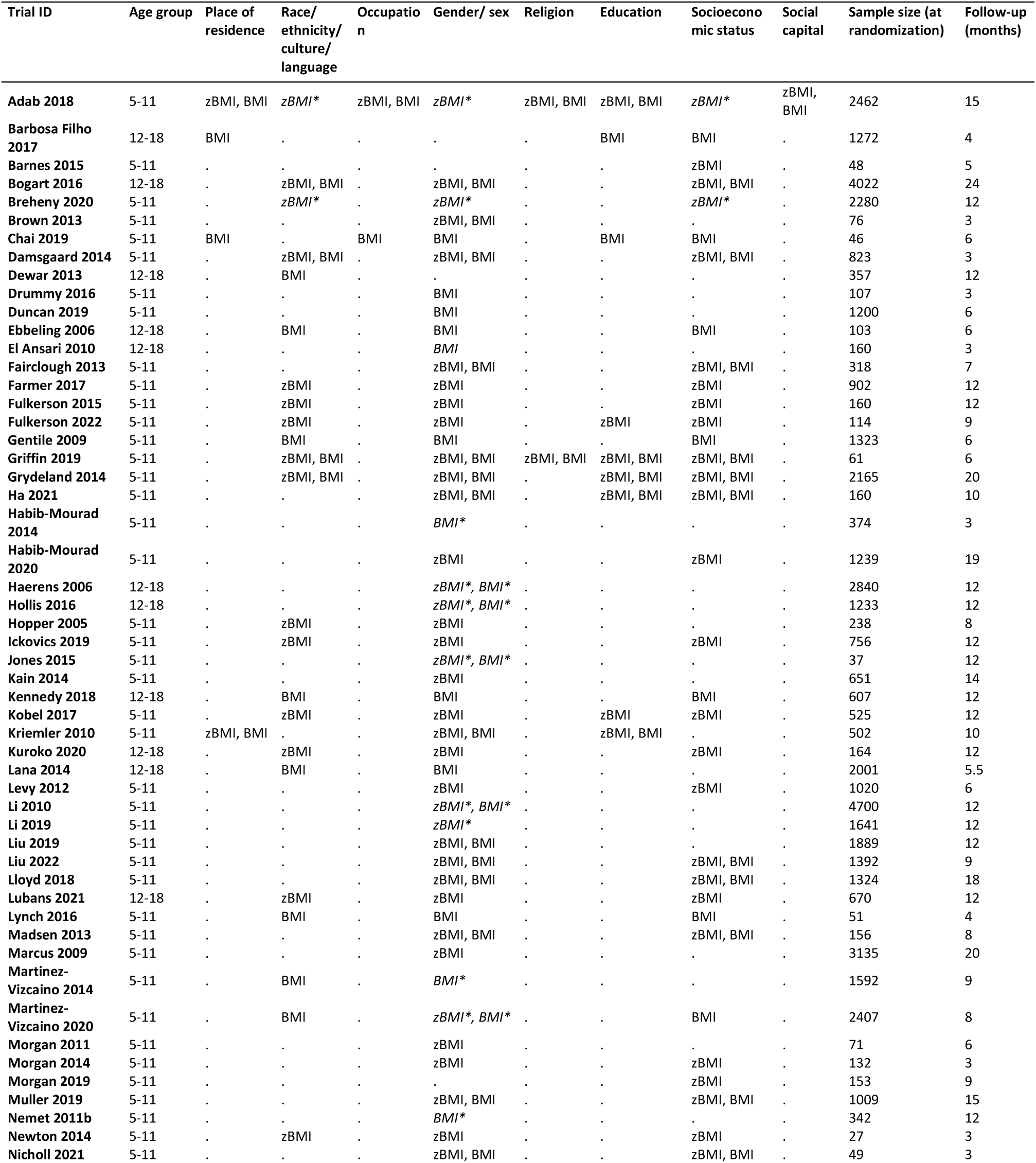
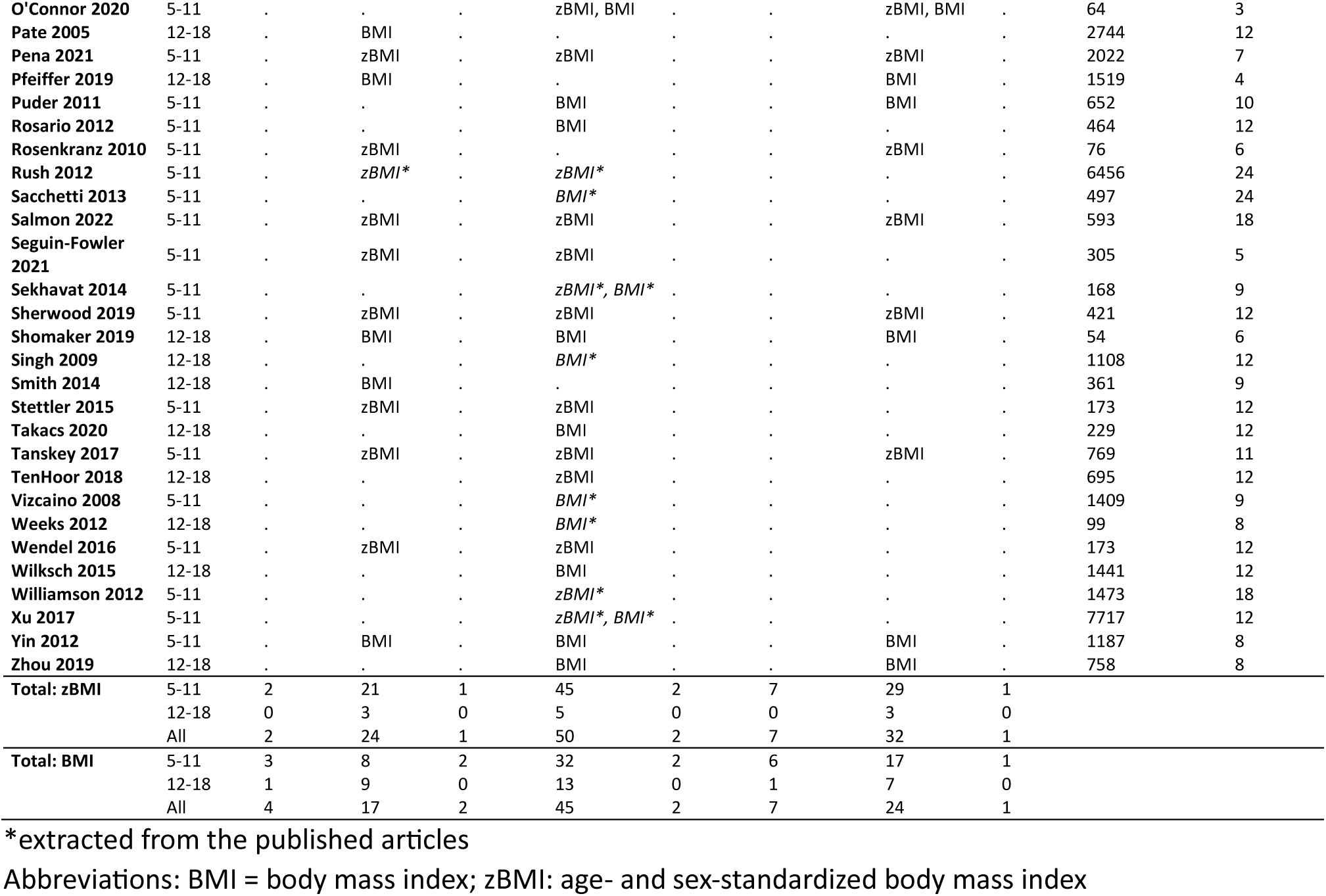
Outcome data obtained from each trial.

| Trial ID | Age group | Place of residence | Race/ethnicity/culture/language | Occupation | Gender/sex | Religion | Education | Socioeconomic status | Social capital | Sample size (at randomization) | Follow-up (months) |
| --- | --- | --- | --- | --- | --- | --- | --- | --- | --- | --- | --- |
| Adab 2018 | 5-11 | zBMI, BMI | zBMI* | zBMI, BMI | zBMI* | zBMI, BMI | zBMI, BMI | zBMI* | zBMI, BMI | 2462 | 15 |
| Barbosa Filho 2017 | 12-18 | BMI | . | . | . | . | BMI | BMI | . | 1272 | 4 |
| Barnes 2015 | 5-11 | . | . | . | . | . | . | zBMI | . | 48 | 5 |
| Bogart 2016 | 12-18 | . | zBMI, BMI | . | zBMI, BMI | . | . | zBMI, BMI | . | 4022 | 24 |
| Breheny 2020 | 5-11 | . | zBMI* | . | zBMI* | . | . | zBMI* | . | 2280 | 12 |
| Brown 2013 | 5-11 | . | . | . | zBMI, BMI | . | . | . | . | 76 | 3 |
| Chai 2019 | 5-11 | BMI | . | BMI | BMI | . | BMI | BMI | . | 46 | 6 |
| Damsgaard 2014 | 5-11 | . | zBMI, BMI | . | zBMI, BMI | . | . | zBMI, BMI | . | 823 | 3 |
| Dewar 2013 | 12-18 | . | BMI | . | . | . | . | . | . | 357 | 12 |
| Drummy 2016 | 5-11 | . | . | . | BMI | . | . | . | . | 107 | 3 |
| Duncan 2019 | 5-11 | . | . | . | BMI | . | . | . | . | 1200 | 6 |
| Ebbeling 2006 | 12-18 | . | BMI | . | BMI | . | . | BMI | . | 103 | 6 |
| El Ansari 2010 | 12-18 | . | . | . | BMI | . | . | . | . | 160 | 3 |
| Fairclough 2013 | 5-11 | . | . | . | zBMI, BMI | . | . | zBMI, BMI | . | 318 | 7 |
| Farmer 2017 | 5-11 | . | zBMI | . | zBMI | . | . | zBMI | . | 902 | 12 |
| Fulkerson 2015 | 5-11 | . | zBMI | . | zBMI | . | . | zBMI | . | 160 | 12 |
| Fulkerson 2022 | 5-11 | . | zBMI | . | zBMI | . | zBMI | zBMI | . | 114 | 9 |
| Gentile 2009 | 5-11 | . | BMI | . | BMI | . | . | BMI | . | 1323 | 6 |
| Griffin 2019 | 5-11 | . | zBMI, BMI | . | zBMI, BMI | zBMI, BMI | zBMI, BMI | zBMI, BMI | . | 61 | 6 |
| Grydeland 2014 | 5-11 | . | zBMI, BMI | . | zBMI, BMI | . | zBMI, BMI | zBMI, BMI | . | 2165 | 20 |
| Ha 2021 | 5-11 | . | . | . | zBMI, BMI | . | zBMI, BMI | zBMI, BMI | . | 160 | 10 |
| Habib-Mourad 2014 | 5-11 | . | . | . | BMI* | . | . | . | . | 374 | 3 |
| Habib-Mourad 2020 | 5-11 | . | . | . | zBMI | . | . | zBMI | . | 1239 | 19 |
| Haerens 2006 | 12-18 | . | . | . | zBMI*, BMI* | . | . | . | . | 2840 | 12 |
| Hollis 2016 | 12-18 | . | . | . | zBMI*, BMI* | . | . | . | . | 1233 | 12 |
| Hopper 2005 | 5-11 | . | zBMI | . | zBMI | . | . | . | . | 238 | 8 |
| Ickovics 2019 | 5-11 | . | zBMI | . | zBMI | . | . | zBMI | . | 756 | 12 |
| Jones 2015 | 5-11 | . | . | . | zBMI*, BMI* | . | . | . | . | 37 | 12 |
| Kain 2014 | 5-11 | . | . | . | zBMI | . | . | . | . | 651 | 14 |
| Kennedy 2018 | 12-18 | . | BMI | . | BMI | . | . | BMI | . | 607 | 12 |
| Kobel 2017 | 5-11 | . | zBMI | . | zBMI | . | zBMI | zBMI | . | 525 | 12 |
| Kriemler 2010 | 5-11 | zBMI, BMI | . | . | zBMI, BMI | . | zBMI, BMI | . | . | 502 | 10 |
| Kuroko 2020 | 12-18 | . | zBMI | . | zBMI | . | . | zBMI | . | 164 | 12 |
| Lana 2014 | 12-18 | . | BMI | . | BMI | . | . | . | . | 2001 | 5.5 |
| Levy 2012 | 5-11 | . | . | . | zBMI | . | . | zBMI | . | 1020 | 6 |
| Li 2010 | 5-11 | . | . | . | zBMI*, BMI* | . | . | . | . | 4700 | 12 |
| Li 2019 | 5-11 | . | . | . | zBMI* | . | . | . | . | 1641 | 12 |
| Liu 2019 | 5-11 | . | . | . | zBMI, BMI | . | . | . | . | 1889 | 12 |
| Liu 2022 | 5-11 | . | . | . | zBMI, BMI | . | . | zBMI, BMI | . | 1392 | 9 |
| Lloyd 2018 | 5-11 | . | . | . | zBMI, BMI | . | . | zBMI, BMI | . | 1324 | 18 |
| Lubans 2021 | 12-18 | . | zBMI | . | zBMI | . | . | zBMI | . | 670 | 12 |
| Lynch 2016 | 5-11 | . | BMI | . | BMI | . | . | BMI | . | 51 | 4 |
| Madsen 2013 | 5-11 | . | . | . | zBMI, BMI | . | . | zBMI, BMI | . | 156 | 8 |
| Marcus 2009 | 5-11 | . | . | . | zBMI | . | . | . | . | 3135 | 20 |
| Martinez-Vizcaino 2014 | 5-11 | . | BMI | . | BMI* | . | . | . | . | 1592 | 9 |
| Martinez-Vizcaino 2020 | 5-11 | . | BMI | . | zBMI*, BMI* | . | . | BMI | . | 2407 | 8 |
| Morgan 2011 | 5-11 | . | . | . | zBMI | . | . | . | . | 71 | 6 |
| Morgan 2014 | 5-11 | . | . | . | zBMI | . | . | zBMI | . | 132 | 3 |
| Morgan 2019 | 5-11 | . | . | . | . | . | . | zBMI | . | 153 | 9 |
| Muller 2019 | 5-11 | . | . | . | zBMI, BMI | . | . | zBMI, BMI | . | 1009 | 15 |
| Nemet 2011b | 5-11 | . | . | . | BMI* | . | . | . | . | 342 | 12 |
| Newton 2014 | 5-11 | . | zBMI | . | zBMI | . | . | zBMI | . | 27 | 3 |
| Nicholl 2021 | 5-11 | . | . | . | zBMI, BMI | . | . | zBMI, BMI | . | 49 | 3 |
| O'Connor 2020 | 5-11 | . | . | . | zBMI, BMI | . | . | zBMI, BMI | . | 64 | 3 |
| Pate 2005 | 12-18 | . | BMI | . | . | . | . | . | . | 2744 | 12 |
| Pena 2021 | 5-11 | . | zBMI | . | zBMI | . | . | zBMI | . | 2022 | 7 |
| Pfeiffer 2019 | 12-18 | . | BMI | . | . | . | . | BMI | . | 1519 | 4 |
| Puder 2011 | 5-11 | . | . | . | BMI | . | . | BMI | . | 652 | 10 |
| Rosario 2012 | 5-11 | . | . | . | BMI | . | . | . | . | 464 | 12 |
| Rosenkranz 2010 | 5-11 | . | zBMI | . | . | . | . | zBMI | . | 76 | 6 |
| Rush 2012 | 5-11 | . | zBMI* | . | zBMI* | . | . | . | . | 6456 | 24 |
| Sacchetti 2013 | 5-11 | . | . | . | BMI* | . | . | . | . | 497 | 24 |
| Salmon 2022 | 5-11 | . | zBMI | . | zBMI | . | . | zBMI | . | 593 | 18 |
| Seguin-Fowler 2021 | 5-11 | . | zBMI | . | zBMI | . | . | . | . | 305 | 5 |
| Sekhavat 2014 | 5-11 | . | . | . | zBMI*, BMI* | . | . | . | . | 168 | 9 |
| Sherwood 2019 | 5-11 | . | zBMI | . | zBMI | . | . | zBMI | . | 421 | 12 |
| Shomaker 2019 | 12-18 | . | BMI | . | BMI | . | . | BMI | . | 54 | 6 |
| Singh 2009 | 12-18 | . | . | . | BMI* | . | . | . | . | 1108 | 12 |
| Smith 2014 | 12-18 | . | BMI | . | . | . | . | . | . | 361 | 9 |
| Stettler 2015 | 5-11 | . | zBMI | . | zBMI | . | . | . | . | 173 | 12 |
| Takacs 2020 | 12-18 | . | . | . | BMI | . | . | . | . | 229 | 12 |
| Tanskey 2017 | 5-11 | . | zBMI | . | zBMI | . | . | zBMI | . | 769 | 11 |
| TenHoor 2018 | 12-18 | . | . | . | zBMI | . | . | . | . | 695 | 12 |
| Vizcaino 2008 | 5-11 | . | . | . | BMI* | . | . | . | . | 1409 | 9 |
| Weeks 2012 | 12-18 | . | . | . | BMI* | . | . | . | . | 99 | 8 |
| Wendel 2016 | 5-11 | . | zBMI | . | zBMI | . | . | . | . | 173 | 12 |
| Wilksch 2015 | 12-18 | . | . | . | BMI | . | . | . | . | 1441 | 12 |
| Williamson 2012 | 5-11 | . | . | . | zBMI* | . | . | . | . | 1473 | 18 |
| Xu 2017 | 5-11 | . | . | . | zBMI*, BMI* | . | . | . | . | 7717 | 12 |
| Yin 2012 | 5-11 | . | BMI | . | BMI | . | . | BMI | . | 1187 | 8 |
| Zhou 2019 | 12-18 | . | . | . | BMI | . | . | BMI | . | 758 | 8 |
| Total: zBMI | 5-11 | 2 | 21 | 1 | 45 | 2 | 7 | 29 | 1 |  |  |
|  | 12-18 | 0 | 3 | 0 | 5 | 0 | 0 | 3 | 0 |  |  |
|  | All | 2 | 24 | 1 | 50 | 2 | 7 | 32 | 1 |  |  |
| Total: BMI | 5-11 | 3 | 8 | 2 | 32 | 2 | 6 | 17 | 1 |  |  |
|  | 12-18 | 1 | 9 | 0 | 13 | 0 | 1 | 7 | 0 |  |  |
|  | All | 4 | 17 | 2 | 45 | 2 | 7 | 24 | 1 |  |  |

\*extracted from the published articles
Abbreviations: BMI = body mass index; zBMI: age- and sex-standardized body mass index

### Risk of bias

Risk of bias in main trial results (as reported in the Cochrane reviews) was ‘Low’ for seven trials (9%), ‘Some concerns’ for 46 (57%), and ‘High’ for 28 (35%) (see Supplementary Table 3). Assessment of the completeness of data (i.e. bias due to missing subgroup data that were extracted from the main analysis papers or were provided by trialists) resulted in ‘Low’ risk of bias for 69 trials (85%), and ‘Some concerns’ for five trials (6%). Of the remaining trials, six trials (7%) were judged differentially for different subgroups results: four trials were ‘Low’ or ‘Some concerns’, and two trials were ‘Low’ or ‘High’. We were not able to assess one trial for completeness of data as the sample size of the overall results were not reported in the main article. Assessment of risk of bias arising from classification of participants into subgroups according to thresholds determined by trialists (bias in selection of the subgroup result) resulted in all trials being judged at ‘Low’ risk of bias.

### Meta-analyses

Forest plots summarizing the main results for the eight PROGRESS factors are provided in Figure 2 for zBMI and Figure 3 for BMI. We present results according to the total number of trials providing data for analysis. Sensitivity analyses using different ICCs in the adjustment for clustering yielded no meaningful differences; confidence intervals were, as is to be expected, slightly narrower for ICC = 0 and slightly wider for ICC = 0.4.

**Figure 2:**
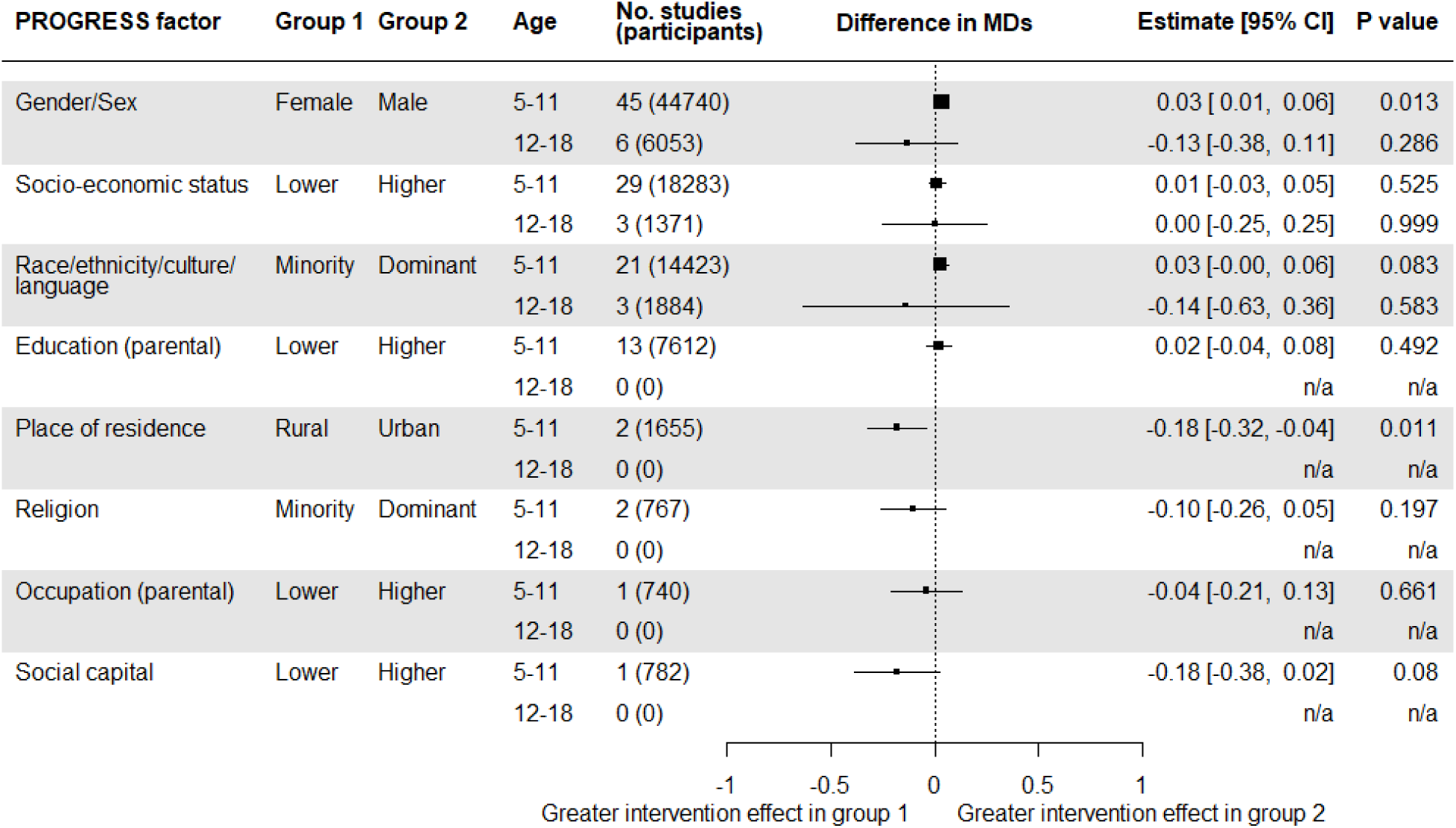
Summary of results for age- and sex-standardized BMI (zBMI): estimates of interaction, expressed as difference in intervention effect (itself expressed as a mean difference) between two inequity factor-based subgroups

**Figure 3:**
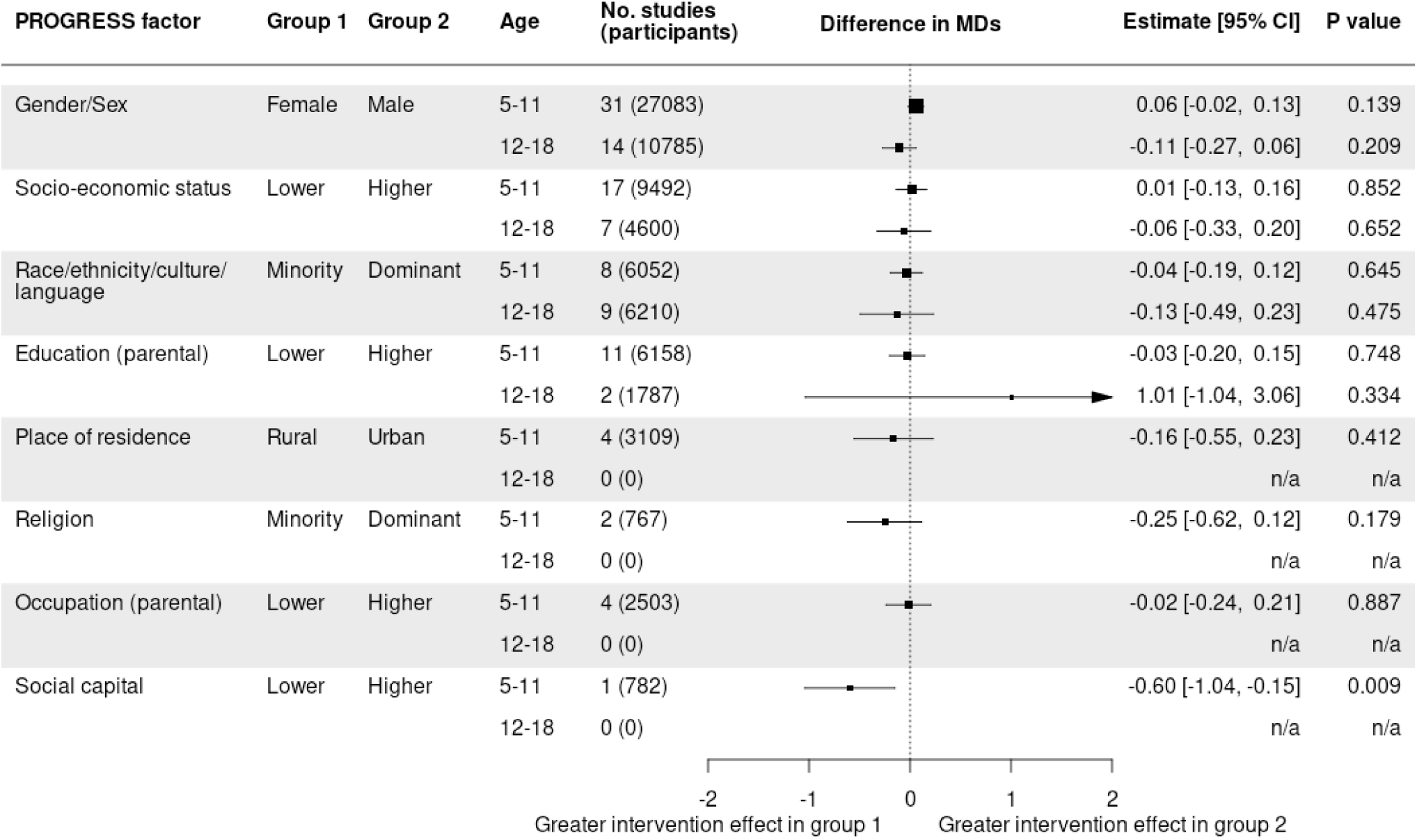
Summary of results for BMI: estimates of interaction, expressed as difference in intervention effect (itself expressed as a mean difference) between two inequity factor-based subgroups

#### Gender/sex

In the younger age group (5-11 years), we included 45 trials (44,740 participants) with zBMI data and 31 trials (27,083 participants) with BMI data. The test for interaction provided evidence of a differential effect of interventions between males and females on zBMI (P=0.01), although not on BMI (P=0.1). In both analyses the intervention effect was greater in males than females. The average magnitude of the difference in MDs was 0.03 (95% CI: 0.01 to 0.06; I^2^=8%, P_het_=0.7) for zBMI, and 0.06 kg/m^2^ (95% CI: −0.02 to 0.13; I^2^=2%, P_het_=0.7) for BMI. Results from individual trials are reported in Supplementary Figure 1 and Supplementary Figure 2 for zBMI and BMI, respectively.

Results of subset analyses provided no evidence that the impact of gender/sex differed for different types of interventions (diet, activity or diet and activity) in the younger age group (test for subset differences P=0.4 for zBMI and P=0.5 for BMI) or for different income country statuses (P=0.8 for zBMI and P=0.9 for BMI) (Supplementary Table 4).

In the older age group (12-18 years), we included 6 trials (6,053 participants) with zBMI data (Figure 2) and 14 trials (10,758 participants) with BMI data (Figure 3). The test for interaction provided little evidence of a differential effect of interventions on zBMI (P=0.3) and BMI (P=0.2), between males and females. The average magnitude of the difference in MDs was −0.13 (95% CI: −0.38 to 0.11; I^2^ 81%, P_het_<0.0001) for zBMI, and −0.11 kg/m^2^ (95% CI: −0.27 to 0.06; I^2^=0%, P_het_=0.4) for BMI. Results from individual trials are reported in Supplementary Figure 3 and Supplementary Figure 4 for zBMI and BMI, respectively.

Results of subset analyses provided some evidence in the older age group that the impact of gender/sex differed for different type of interventions for zBMI as an outcome but not on unstandardized BMI (test for subset differences P=0.001 for zBMI, P=0.8 for BMI), with interventions targeting both diet and activity being more effective in males. There was little evidence that the impact of gender/sex differed by country income status (P=0.4 for BMI). We were not able to perform a subset analysis for zBMI because all trials in the older age group that provided zBMI data by sex were from high income countries (Supplementary Table 5).

#### Socioeconomic status

We included 29 trials (18,283 participants) with zBMI data (Figure 2) and 17 trials (9,492 participants) with BMI data (Figure 3) in the younger age group. There was no evidence of a differential effect of interventions between children with higher and lower socioeconomic status (P=0.5 for zBMI; P=0.9 for BMI) and the average magnitude of the difference in MDs was near to zero: 0.01 (95% CI: −0.03 to 0.05; I^2^=46%, P_het_=0.008) for zBMI and 0.01 kg/m^2^ (95% CI: −0.13 to 0.16; I^2^=49%, P=0.003) for BMI. Results from individual trials are reported in Supplementary Figure 8 for zBMI and Supplementary Figure 6 for BMI. Subset analyses provided no evidence that the impact of socioeconomic status differed for different income country status (P=0.9 for zBMI and P=0.2 for BMI): see Supplementary Table 4.

In the older age group (12-18 years), we included 3 trials (1,308 participants) with zBMI (Figure 2) and 7 trials (4,600 participants) with BMI data (Figure 3). Again, the test for interaction provided no evidence of a differential effect of interventions between participants with higher socioeconomic status and participants with lower socioeconomic status (P=1 for zBMI; P=0.7 for BMI). The average magnitude of the difference in MDs was 0.0001 (95% CI: −0.25 to 0.25; I^2^=28%, P_het_=0.3) for zBMI, and −0.06 kg/m^2^ (95% CI: −0.33 to 0.20; I^2^=0%, P_het_=1.0) for BMI. Results from individual trials are reported in Supplementary Figure 7 for zBMI and Supplementary Figure 8 for BMI. Subset analyses provided little evidence that the impact of socioeconomic status differed for different income country status (P=0.8 for BMI). We were not able to perform a subset analysis for zBMI because all trials in the older age group that provided zBMI data by socioeconomic status were from high income countries. Full results of the subset analyses are reported in Supplementary Table 5.

#### Race/ethnicity/culture/language

In the younger age group (5-11 years), we included 21 trials (14,423 participants) with zBMI (Figure 2) and 8 trials (6,052 participants) with BMI data (Figure 3). The test for interaction provided little evidence of a differential effect of interventions on zBMI (P=0.08) and BMI (P=0.6), between participants in the dominant ethnic group and participants in the minority ethnic group. The average magnitude of the difference in MDs was 0.03 (95% CI: −0.004 to 0.06; I^2^=2%, P_het_=0.5) for zBMI, and −0.04 kg/m^2^ (95% CI: −0.19 to 0.12; I^2^=0%, P_het_=0.6) for BMI. Results from individual trials are reported in Supplementary Figure 9 and Supplementary Figure 10 for zBMI and BMI, respectively.

In the older age group (12-18 years) we included 3 trials (1,884 participants) with zBMI (Figure 2) and 9 trials (6,210 participants) with BMI data (Figure 3). The test for interaction provided no evidence of a differential effect of interventions on zBMI (P=0.6) and BMI (P=0.5), between participants in the dominant ethnic group and participants in the minority ethnic group. The average magnitude of the difference in MDs was −0.14 (95% CI: −0.63 to 0.36; I^2^=74%, P_het_=0.01) for zBMI, and −0.13 kg/m^2^ (95% CI: −0.49 to 0.23; I^2^=20%, P_het_=0.2) for BMI. Results from individual trials are reported in Supplementary Figure 11 and Supplementary Figure 12 for zBMI and BMI, respectively.

#### Education (parental)

Thirteen trials (7,216 participants) with zBMI (Figure 2) and 11 trials (6,158 participants) with BMI (Figure 3) provided data on parental education status for the younger age group. The test for interaction did not indicate a differential effect of interventions on zBMI (P=0.5) and BMI (P=0.7), between participants in the higher parental education group and participants in the lower parental education group. The average magnitude of the difference in MDs was 0.02 (95% CI: −0.04 to 0.08; I^2^ =24%, P_het_=0.2) for zBMI, and −0.03 kg/m^2^ (95% CI: −0.20 to 0.15; I^2^=43%, P_het_=0.02) for BMI. Results from individual trials are reported in Supplementary Figure 13 and Supplementary Figure 14 for zBMI and BMI, respectively. Subset analyses did not provide evidence that impact of parental education differed for different income country status (P=0.3 for zBMI and P=0.1 for BMI). Full results of the subset analyses are reported in Supplementary Table 4.

In the older age group (12-18 years), two trials (1,718 participants) provided BMI data (Figure 3) and no trials zBMI data. The test for interaction showed no evidence of a differential effect of interventions on BMI (P=0.3), between participants in the higher parental education group and participants in the lower parental education group. The average magnitude of the difference in MDs was 1.01 kg/m^2^ (95% CI: −1.04 to 3.06; I^2^=73%, P_het_=0.06). Results from individual trials are reported in Supplementary Figure 15. Results of subset analyses support little evidence that the impact of parental education differed for different income country status (P=0.1 for BMI). Full results of the subset analyses are reported in Supplementary Table 5.

#### Place of residence

For place of residence (urban versus rural), we included two trials (1655 participants) in the younger age group with zBMI data (Figure 2) and four trials (3109 participants) in the younger age group with BMI data Figure 3). The test for interaction provides some evidence of a differential effect of interventions between participants in urban locations and participants living in rural locations on zBMI (P=0.01) but this was not replicated for BMI (P=0.4). The intervention appeared to be more effective in participants living in rural locations, with the average magnitude of the difference being −0.18 (95% CI: −0.32 to −0.04; I^2^=0%, P_het_=0.8) for zBMI, and −0.16 kg/m^2^ (95% CI: −0.55 to 0.23; I^2^=60%, P_het_=0.1) for BMI. Results from individual trials are reported in Supplementary Figure 16 and Supplementary Figure 17 for zBMI and BMI, respectively. No trials provided subgrouping by place of residence for the older age group. All trials that provided data by place of residence were from high income countries, so we were not able to perform a subset analysis on this factor.

#### Religion

We included two trials (767 participants) in the younger age group with zBMI (Figure 2) and BMI data (Figure 3). The test for interaction provided no evidence of a differential effect of interventions on zBMI (P=0.2) and BMI (P=0.2) between participants in the dominant religion group and participants in minority religion group. The average magnitude of the difference in MDs was −0.10 (95% CI: −0.26 to 0.05; I^2^=0%, P=0.8) for zBMI, and −0.25 kg/m^2^ (95% CI: −0.62 to 0.12; I^2^=0%, P_het_=0.4) for BMI. Results from individual trials are reported in Supplementary Figure 18 and Supplementary Figure 19 for zBMI and BMI, respectively. No trials provided results by religion in the older age group. Both trials that provided data by religion were from high income countries, so we were not able to perform a subset analysis on this factor.

#### Occupation (parental)

In the younger age group (5−11 years) we included 1 trial (740 participants) with zBMI (Figure 2) and 4 trials (2,503 participants) with BMI data (Figure 3). There was no evidence of a differential effect of interventions on zBMI (P=0.7) and BMI (P=0.9), between participants in the higher occupation group and participants in the lower occupation group. The average magnitude of the difference in MDs was −0.02 (95% CI: −0.24 to 0.21) for zBMI, and −0.04 kg/m^2^ (95% CI: −0.21 to 0.13) for BMI. Results from individual trials are reported in Supplementary Figure 20 and Supplementary Figure 21 for zBMI and BMI, respectively. No trials provided results by occupation in the older age group. All trials providing data by parental occupation were from high income countries, so we were not able to perform a subset analysis on this factor.

#### Social capital

We included only a single trial (782 participants) in the younger age group and none in the older age group for the factor social capital. In this trial, children were divided into a subgroup in which their parents had a spouse in the same house and a subgroup with a different family structure. The test for interaction provided no evidence of a differential effect of intervention on zBMI (P=0.7; Figure 2) and BMI (P=0.9; Figure 3), between participants in the higher social capital group and participants in the lower social capital group. The magnitude of the difference in MDs was −0.18 (95% CI: −0.38 to 0.02) for zBMI, and −0.6 kg/m^2^ (95% CI: −1.04 to −0.15) for BMI (Supplementary Figure 22 and Supplementary Figure 23). This trial was conducted in a high-income country.

## Discussion

To the best of our knowledge, this is the first meta-analysis using primary data from randomized trials of interventions to prevent obesity in children and young people to assess the potential impact of these interventions on health equity. Our findings stem from re-analysis of data, mostly provided directly by the trialists, from 81 of the eligible 244 trials included in two recent Cochrane reviews.^24,25^

We found that there was no substantial impact of the interventions on inequalities as measured by the eight PROGRESS factors. We did, however, observe that interventions had more beneficial impacts on zBMI in younger boys (5-11 years) than in younger girls (based on data from 45 trials), although such a difference was not observed for older children (12-18 years). This supports the findings of previous trials in primary schools of multi-component physical activity interventions.^33,34^ In the UK and many other countries, physical education at primary school is taught in mixed groups of boys and girls, while for older children there is more variation in the use of single or mixed gender physical education. Our findings suggest that interventions for younger children may benefit from being equally engaging and enjoyable for females and males. However, in the older age group, where increased BMI may correspond with decreased percentage body fat in males but not females,^35^ a more beneficial effect of interventions in males compared with females could be masked by examining BMI only. More evidence is required from studies that use better proxies of percentage body fat.

Two other signals were based on very small numbers of trials, so should be interpreted more cautiously. The first of these was that in the younger age group, the intervention effects on zBMI were greater for those living in a rural community compared with an urban community. One of the two trials contributing to this analysis included significant involvement from the local premier team sports (football) club, role models and family campaigns. The second was that, again in the younger age group and based on the same trial with local premier team sports club involvement, the effect of intervention on BMI was greater in children from families with low social capital.

Our results concur with previous work addressing the same question, using secondary data.^36,37^ Our findings suggest that policy makers, commissioners and providers can be confident in promoting the types of interventions to prevent obesity in children and young people included in the two Cochrane reviews that were found to have a beneficial effect on BMI or zBMI independently of PROGRESS inequity factors, thus not increasing inequities. These reviews found beneficial interventions for interventions that promoted physical activity (only) over the medium-term (9-15 months) and long-term (>15 months) (mean difference around −0.30 kg/m^2^ BMI) for adolescents aged 12-18 years and over the medium term (−0.1 kg/m^2^ BMI, −0.05 zBMI) for children aged 5-11 years. Beneficial effects were also found for interventions that promoted physical activity alongside healthy eating in the short-term (3-9 months) and medium-term (−0.11 kg/m^2^ BMI, around −0.04 zBMI) for 5-11 year olds. Although these beneficial effects are small, when delivered at scale, the effects of these preventive interventions have shown through modelling (for England) to have the potential to contribute meaningfully to reducing the prevalence of childhood obesity.^38^

Most of the interventions included in the two Cochrane reviews were operating in the ‘organizational’ or ‘community’ domains of the socio-ecological model and were on the ‘guide’ or ‘enable’ steps of the Nuffield intervention ladder.^19^ There is therefore a theoretical basis for assuming these types of interventions may increase health inequity. We see no clear indication that the few trials observing reductions in inequalities differed from other trials in the characteristics (or combinations of characteristics) we coded that are proposed to reduce inequalities or drive intervention-generated inequalities. That said, our coding method was relatively blunt and there was little variation between trials on some characteristics. Many of the interventions that we coded as ‘Universal’ rather than ‘Targeted’ were set in urban areas of large cities with relatively high levels of deprivation in parts. Similarly, most interventions operated in the same domains of the socio-ecological model and on the same steps of the Nuffield intervention ladder. Our limited findings do not support those of Moore et al^39^ in suggesting that interventions including a change in the school physical environment may reduce SES inequalities, noting that the majority of changes identified in the trial were in the school setting and focussed either on the food environment or both the food environment and the built environment.

Strengths of our study include the collection of subgrouped trial data directly from the trialists. Our response rates of trials 58% for successful contact and 28% for data collection were much higher than we anticipated, allowing sizeable sample sizes for some of the meta-analyses. We suspect that our personalized emails and convenient data collection methods may have contributed partly to this. We had originally planned to seek individual participant data from the trials, which would have been a much more laborious process, potentially with a lower response rate and certainly with more administrative burden through data sharing agreements. A further strength of our results is the lack of discernible heterogeneity in the interaction estimates across studies, providing some justification for the crude combination of findings from trials with very different intervention strategies.

The study is not without limitations. While response rates were high for this kind of research, we still included only a minority of the trials identified by the Cochrane reviews, and numbers of trials for some of the PROGRESS factors were very small indeed. Baseline inequity factors were dichotomized to facilitate straightforward meta-analysis of interactions, which is simplistic and may disguise more nuanced associations. Moreover, different dichotomizations were used in different trials, and most were selected by the trialists, although we did not judge that this introduced a high risk of bias. None of the trials evaluated a ‘whole systems approach’. Finally, we planned to examine factors related to inequity as represented by the PROGRESS framework and did not explore other dimensions such as intervention effects related to baseline weight status; we will report the results of our investigation into this elsewhere.

Most organizations responsible for public health guidance on how best to prevent childhood obesity and reduce heath inequalities advocate a ‘whole systems approach’, including the World Health Organization and National Institute for Health and Care Excellence (NICE) in the UK, as do commissioners of local services such as Local Authorities in England. Although randomized trials of whole systems approaches are few, some trials are adapting traditional methods and providing important findings, such as the ‘Healthy Together Victoria’ study.^40^ We encourage all trialists undertaking this pioneering work to measure PROGRESS equity factors at baseline and assess the effect of their intervention on inequalities. In practice, whole systems approaches comprise separate interventions (components) interconnected via a programme theory and logic model. Many of the interventions included in the two Cochrane reviews that are the basis for the work presented in this paper are representative of such components. As such, the findings of this health equity analysis are relevant to those providing guidance on, or commissioning, a whole systems approach to preventing obesity in children and young people alongside promoting health equity.

## Inequity in Obesity Prevention Trialists Collaborative Group

Trialists: Arne Astrup, Valter Cordeiro Barbosa Filho, Mark E Benden, Lynne Boddy, Laura M Bogart, Blakely Brown, Angela Carlin, Diana P Pozuelo Carrascosa, Li Kheng Chai, Clare Drummy, Scott Duncan, Cara Ebbeling, Eva Martos, Stuart Fairclough, Jayne Fulkerson, Douglas A Gentile, Mary B Gruber, May Grydeland, Amy S Ha, Carla Habib Mourad, Kate Gilstad-Hayden, Douglas L Hill, Gill ten Hoor, Kiya Hurley, Alison Hurst, Nahla Hwalla, Jeannette R Ickovics, Kate Jolly, Juliana Kain, Susanne Kobel, Viktoria Anna Kovacs, Susi Kriemler, Sarahmarie Kuroko, Alberto Lana, Teresa Shamah Levy, Mairena Sánchez-López, David Lubans, Brian Lynch, Kristine A Madsen, Claude Marcus, Méndez-Gómez Humarán, Carmen Morales-Ruan, Philip Morgan, Ivan Müller, Robert Newton, Analise Nicholl, Teresia O’Connor, Russell R Pate, Sebastián Peña, Lorraine B Robbins, Jardena J Puder, Thomas Robinson, Rafaela Rosário, Richard Rosenkranz, Jennifer Sacheck, Jo Salmon, Rebecca A Seguin-Fowler, Nancy E Sherwood, Hajnalka Takacs, Rachael Taylor, Haixue Wang, Haijun Wang, Robin Whittemore, Simon Wilksch, Zenong Yin, Zhixiong Zhou.

Other project team members: Katie Breheny, Deborah M Caldwell, Sarah Dawson, Yang Gao, Frances Hillier-Brown, Rebecca K Hodder, Sofus C Larsen, Theresa HM Moore, James D Nobles, Sophie M Phillips, Jelena Savović, Fanney Thorsteinsdottir, Eve Tomlinson, Luke Wolfenden.

### Affiliations

Department of Nutrition, Exercise and Sports, University of Copenhagen, Denmark (Astrup A); Universidade Estadual do Ceará, Brazil (Barbosa Filho VC); Texas A&M School of Public Health, Texas A&M University, TX, Usa (Benden ME); The Physical Activity Exchange, Research Institute for Sport and Exercise Sciences, Liverpool John Moores University, UK (Boddy L); RAND Corporation, Santa Monica, CA, USA (Bogart LM); School of Public and Community Health Sciences, University of Montana, Missoula, MT, USA (Brown B); Centre for Exercise Medicine, Physical Activity and Health, Sports and Exercise Sciences Research Institute, Ulster University, Belfast, UK (Carlin A); School of Human Movement and Nutrition Sciences, The University of Queensland, Brisbane, QLD, Australia (Chai LK); Southern Health and Social Care Trust, Southern College of Nursing, Craigavon Area Hospital, Portadown, UK (Drummy C); School of Sport and Recreation, Auckland University of Technology, Auckland, New Zealand (Duncan S); New Balance Foundation Obesity Prevention Center, Boston Children’s Hospital, Boston, MA, USA; Department of Pediatrics, Harvard Medical School, Boston, MA, USA (Ebbeling C); University of Physical Education, Budapest, Hungary (Eva E, Takacs H); Sport and Physical Activity Department, Hedge Hill University, UK (Fairclough S); Clinical and Translational Science Institute, University of Minnesota, MN, USA (Fulkerson J); Department of Psychology, Iowa State University, IA, USA (Gentile DA); Yale School of Public Health and Professor of Psychology, Yale University, CT, USA (Gilstad-Hayden K, Ickovics JR); California State Polytechnic University Humboldt, Arcata, CA, USA (Gruber MB); Department of Physical Performance, Norwegian School of Sport Sciences, Oslo, Norway (Grydeland M); Department of Sports Science and Physical Education, The Chinese University of Hong Kong, China (Ha AS); Department of Nutrition and Food Sciences, Faculty of Agriculture and Food Sciences, American University of Riad El-Solh, Beirut, Lebanon (Habib Mourad C, Hwalla N); The Children’s Hospital of Philadelphia, PA, USA (Hill DL); School of Geography, Earth and Environmental Sciences, University of Birmingham, Uk (Hurley K); University of Exeter Medical School, Exeter, UK (Hurst A); Institute of Applied Health Research, University of Birmingham, UK (Jolly K); Instituto de Nutrición y Tecnología de Alimentos (INTA), University of Chile (Kain J); Ulm University Hospital, Division of Sports- and Rehabilitation Medicine, Ulm, Germany (Kobel S); Division of Nutrition Physiology and Epidemiology, National Institute of Pharmacy and Nutrition, Budapest, Hungary (Kovacs VA); Epidemiology, Biostatistics and Prevention Institute, University of Zurich, Switzerland (Kriemler S); Department of Women’s and Children’s Health, University of Otago, New Zealand (Kuroko S); Department of Preventive Medicine and Public Health. University of Oviedo/ISPA, Spain (Lana A); Centro de Investigación en Evaluación y Encuestas, Instituto Nacional de Salud Pública, Cuernavaca, Morelos, México (Levy TS, Morales-Ruan C); Centre for Active Living and Learning, College of Human and Social Futures, University of Newcastle, Newcastle, NSW, Australia; Faculty of Sport and Health Sciences, University of Jyväskylä, Jyväskylä, Finland; Hunter Medical Research institute, New Lambton Heights, NSW, Australia (Lubans D); Mayo Clinic Rochester, Rochester, MN, USA (Lynch B); UC Berkeley, School of Public Health, Berkeley, CA, USA (Madsen KA); Department of Clinical Science, Intervention and Technology, Karolinska Institutet, Sweden (Marcus C); Centro de Investigación en Matemáticas A.C., Unidad Aguascalientes, Aguascalientes, México (Méndez-Gómez H); Centre for Active Living and Learning, College of Human and Social Futures, School of Education, University of Newcastle, Callaghan, NSW, Australia (Morgan P); Department of Sport, Exercise and Health, University of Basel, Switzerland (Müller I); Pennington Biomedical Research Centre, Baton Rouge, LA, USA (Newton R); Exercise Medicine Research Institute, School of Medicine and Health Sciences, Edith Cowan University, Australia (Nicholl A); USDA/ARS Children’s Nutrition Research Center, Huston, TX, USA (O’Connor T); Department of Exercise Science, University of South Carolina, Columbia, SC, USA (Pate RR); Finnish Institute for Health and Welfare, Mannerheimintie 166 Helsinki, Finland (Peña S); MSU College of Nursing, Michigan State University, MI, USA (Pfeiffer LB); University of Castilla-La Mancha, Faculty of Nursing, Cuenca, Spain (Pozuelo Carrascosa DP); Obstetric service, Department Woman-Mother-Child, Lausanne University Hospital, Lausanne, Switzerland (Puder JJ); Department of Pediatrics, Stanford University, USA (Robinson T); School of Nursing, Health Sciences Research Unit: Nursing(UICISA: E), Nursing School of Coimbra(ESEnfC), University of Minho, Coimbra, Portugal. (Rosário R); Kinesiology and Nutrition Sciences, University of Nevada, Las Vegas, NE, USA (Rosenkranz R); Department of Exercise and Nutrition Sciences, Milken Institute School of Public Health, The George Washington University, DC, USA (Sacheck J); Institute for Physical Activity and Nutrition, Deakin University, VIC, Australia (Salmon J); Universidad de Castilla-La Mancha, School of Education, Ciudad Real, Spain and Health and Social Research Center, Cuenca, Spain (Sánchez-López M); Institute for Advancing Health through Agriculture, Texas A&M University System, College Station, TX, USA (Seguin-Fowler RA); Division of Epidemiology and Community Health, School of Public Health, University of Minnesota, MN, USA (Sherwood NE); Department of Medicine, University of Otago, New Zealand (Taylor R); Dept. Work and Social Psychology, Faculty of Psychology and Neuroscience, Maastricht University, The Netherlands (Ten Hoor G); Peking University Health Science Center, China (Wang H, Wang H); Yale School of Nursing, Yale University, CT, USA (Whittemore R); College of Education, Psychology & Social Work, Flinders University, South Australia, Australia, Advanced Psychology Services, Adelaide, SA, Australia (Wilksch S); Institute for Sport Performance and Health Promotion, Capital University of Physical Education and Sports, Beijing, China (Yin Z, Zhou Z); Population Health Sciences, Bristol Medical School, University of Bristol, Bristol, UK (Breheny K, Caldwell DM, Dawson S, Moore THM, Savović J, Tomlinson E); Department of Sport, Physical Education and Health, Hong Kong Baptist University, Kowloon, Hong Kong (Gao Y); Human Nutrition Research Centre and Population Health Sciences Institute, University of Newcastle, Newcastle, UK (Hillier-Brown F); School of Medicine and Public Health, The University of Newcastle, Australia (Hodder RK, Wolfenden G); Faculty of Medical and Health Sciences, University of Copenhagen, Copenhagen, Denmark (Larsen S, Thorsteinsdottir F); Health, Nutrition & Environment, Leeds Beckett University, UK (Nobles JD), Department of Sport and Exercise Science, Durham University, Durham, UK and Child Health and Physical Activity Laboratory, School of Occupational Therapy, Western University, London, Ontario, Canada (Phillips SM).

## Contributions

- Conceptualization: JPT Higgins, CD Summerbell and BL Heitmann
- Formal analysis: AL Davies and JC Palmer, with lesser contributions from F Spiga and JPT Higgins
- Funding acquisition: JPT Higgins and CD Summerbell
- Investigation: generation and contribution of trial data: all members of the *Inequity in Obesity Prevention Trialists Collaborative Group*; data collection and processing: JC Palmer and AL Davies; coding intervention characteristics: CD Summerbell; interpretation: AL Davies, F Spiga, BL Heitmann, R Jago, CD Summerbell and JPT Higgins
- Methodology: JPT Higgins and AL Davies
- Project administration: JC Palmer
- Supervision: JPT Higgins
- Writing – original draft preparation: AL Davies, F Spiga, CD Summerbell and JPT Higgins
- Writing – review & editing: JC Palmer, AL Davies, F Spiga, BL Heitmann, R Jago, CD Summerbell and JPT Higgins

## Data Availability

All data produced in the present study are available upon reasonable request to the authors

## Acknowledgements

This work was funded by the National Institute for Health and Care Research (NIHR) Public Health Programme (grant number NIHR131572). FS and JPTH were supported in part by the NIHR Bristol Evidence Synthesis Group. JPTH and RJ were supported in part by the NIHR Applied Research Collaboration West (ARC West) at University Hospitals Bristol and Weston NHS Foundation Trust. JPTH is a NIHR Senior Investigator (grant number NIHR203807). The views expressed in this paper are those of the authors and do not necessarily reflect those of the NIHR or Department of Health and Social Care.

## Data sharing statement

The protocol for this work was published at https://fundingawards.nihr.ac.uk/award/NIHR131572 as an appendix to the April 2023 version of the wider project protocol. Only aggregated data by subgroup were collected from the trialists. These data are presented in the figures in the supplementary materials to the paper and are available by request from the authors.

