## Supplementary Material for "Do the effects of interventions aimed at the prevention of childhood obesity reduce inequities? A re-analysis of randomized trial data from two Cochrane reviews"

Supplementary Figure 15: Estimates of intervention effect for separate subgroups (left) and differences in intervention effect between subgroups (interactions; right) for factor **(parental) education** and outcome **BMI** in the **older age group** (12-18 years) 47

Supplementary Figure 20: Estimates of intervention effect for separate subgroups (left) and differences in intervention effect between subgroups (interactions; right) for factor **place of residence** and outcome **zBMI** in the **younger age group** (5-11 years)48

Supplementary Figure 21: Estimates of intervention effect for separate subgroups (left) and differences in intervention effect between subgroups (interactions; right) for factor **place of residence** and outcome **BMI** in the **younger age group** (5-11 years) 49

Supplementary Table 1: Characteristics of included trials

| **Study ID**<br>**Country (income of the country)** | **Name of study (if reported)**<br>**Study design (unit of allocation)**<br>**Study setting and location** | **N participants (intervention(s); control)** | **Age (mean years)** | **Gender/Sex** | **Intervention type**<br>**Comparator type**<br>**Setting of intervention**<br>**Duration of intervention** |
|---|---|---|---|---|---|
| Adab 2018<br>United Kingdom<br>(high income) | WAVES study (West Midlands ActiVe lifestyle and healthy Eating in School children study)<br>cluster RCT (school)<br>Fifty-four state primary schools in the West Midlands | 2462 (1134; 1328) | 6.3 (SD 0.3) | 51.1% boys | Dietary and activity intervention<br>No active intervention<br>12 months<br>School + community |
| Barbosa Filho 2017<br>Brazil<br>(upper middle income) | Fortaleça sua Saúde<br>cluster RCT (school)<br>Six full-time schools of the city that were linked to the national program School Health Program in Fortaleza | 1272 (639; 633) | Age range: 11–13 years 52.9%; age range 14–18 years 47.1% | 51.5% boys | Activity intervention<br>No active intervention<br>4 months<br>School |
| Barnes 2015<br>Australia<br>(high income) | MADE4Life Program<br>cluster RCT (mother + ≥ 1 daughter)<br>An Australian community | 48 (25; 23) | 8.5 (SD 1.7) | 100% girls | Activity intervention<br>No active intervention<br>8 weeks<br>Community |
| Bogart 2016<br>United States<br>(high income) | SNaX (Students for Nutrition and Exercise)<br>cluster RCT (school)<br>Ten schools in Los Angeles Unified School District (LAUSD), California | 4022 (1954; 2068) | 12.2 (SD 0.68) | 49.1% boys | Dietary and activity intervention<br>No active intervention<br>5 weeks<br>School + home |
| Breheny 2020<br>United Kingdom<br>(high income) | Daily Mile<br>cluster RCT (school)<br>Fourty trimary schools in the South of Birmingham | 2280 (1153; 1127) | 8.9 (SD 1) | 52.4% boys | Activity intervention<br>No active intervention<br>12 months<br>School |
| Brown 2013<br>United States<br>(high income) | Journey to Native Youth Health<br>RCT (individual)<br>Two American Indian reservations in north-central and southwestern Montana | 76 (38; 38) | 11.4 (SD 1.1) | 50% boys | Dietary and activity intervention<br>Attention control<br>12 weeks<br>Community |
| Chai 2019<br>Australia<br>(high income) | Back2Basics (Family telehealth consultations)<br>RCT (parent/child dyad)<br>Communities in New South Wales, New Castle, Tamworth, Armidale | 46 (Back2Basics family intervention (telehealth): 16<br>Back2Basics family intervention (telehealth + SMS): 15; 15) | 9 (SD 2.3) | 59% boys | Dietary intervention<br>No active intervention<br>12 weeks<br>Telehealth |

| | | | | | |
|---|---|---|---|---|---|
| Damsgaard 2014<br>Denmark<br>(high income) | OPUS (The Optimal Well-Being, Development and Health for Danish Children through a Healthy New Nordic Diet (OPUS) School Meal Study)<br>cluster RCT (school)<br>Nine primary schools in Zealand and Lolland-Falster | 823 (398; 425) | 10 (SD 0.6) | 52.1% boys | Dietary intervention<br>No active intervention<br>3 months<br>School |
| Dewar 2013<br>Australia<br>(high income) | NEAT Girls (Nutrition and Enjoyable Activity for Teen Girls)<br>cluster RCT (school)<br>Twelve government secondary schools in the Hunter Region and Central Coast areas in New South Wales | 357 (178; 179) | 13.2 (SD 0.5) | 100% girls | Dietary and activity intervention<br>No active intervention<br>12 months<br>School |
| Drummy 2016<br>United Kingdom<br>(high income) | NR<br>cluster RCT (classroom)<br>Seven primary schools in Northern Ireland | 107 (54; 53) | 9.5 | NR | Activity intervention<br>No active intervention<br>12 weeks<br>School |
| Duncan 2019<br>New Zealand<br>(high income) | Healthy Homework<br>cluster RCT (school)<br>Sixteen primary schools from Auckland and Dunedin | 1200 (600; 600) | Intervention: 8.71 (SD 0.99)<br>Control: 8.74 (SD 1.04) | 48.3% boys | Dietary and activity intervention<br>No active intervention<br>8 weeks<br>School |
| Ebbeling 2006<br>United States<br>(high income) | BASH - Beverages and Student Health<br>RCT (individual)<br>Homes | 103 (53; 50) | Intervention: 16 (SD 1.1)<br>Control: 15.8 (SD 1.1) | Intervention: 45% boys<br>Control 46% boys | Dietary Intervention<br>No active intervention<br>25 weeks<br>Home + telehealth |
| El Ansari 2010<br>Egypt<br>(lower middle income) | NR<br>RCT (individual)<br>One secondary school with both indoor and outdoor sport facilities and sport equipment in Mansoura City | 160 (80; 80) | Intervention: 15.7 (SD 1.8)<br>Control: 15.4 (SD 1.6) | 43.75% boys | Activity intervention<br>No active intervention<br>3 months<br>School (after school programme) |
| Fairclough 2013<br>United Kingdom<br>(high income) | CHANGE! (Children's health, Activity and Nutrition: Get Educated!)<br>cluster RCT (school)<br>Twelve primary schools in the Wigan Borough in northwest England | 318 (166; 152) | Intervention: 10.6 (SD 0.3)<br>Control: 10.7 (SD 0.3) | NR | Dietary and activity intervention<br>No active intervention<br>20 weeks<br>School |
| Farmer 2017<br>New Zealand<br>(high income) | PLAY<br>cluster RCT (school)<br>Sixteen state primary schools in the Otago region and Waitakere City (within the Auckland region) | 902 (458; 444) | Intervention: 8.0 (SD 1.2)<br>Control: 7.9 (SD 1.1) | 53.6% boys | Activity intervention<br>No active intervention<br>1 year<br>School |
| Fulkerson 2015<br>United States | HOME Plus (Healthy Home Offerings via the Mealtime Environment Plus Study) | 160 (81; 79) | 10.3 (SD 1.4) | 53% boys | Dietary intervention<br>Attention control |

| | | | | | |
|---|---|---|---|---|---|
| (high income) | RCT (staggered-cohort design - see notes)<br>(parent/child dyad)<br>Minneapolis | | | | 10 months<br>Home + community |
| Fulkerson 2022<br>United States<br>(high income) | NU-HOME (New Ulm at HOME - Healthy Home Offerings via the Mealtime Environment)<br>RCT (staggered-cohort design - see notes)<br>(parent/child dyad)<br>New Ulm or Sleepy Eye communities, Minnesota | 114 (58; 56) | 9 (SD 1.1) | 41.2% boys | Dietary and activity intervention<br>No active intervention<br>7 months<br>Home + community |
| Gentile 2009<br>United States<br>(high income) | Switch programme (Switch what you do, view, and chew)<br>cluster RCT (school)<br>Ten elementary schools in Lakeville, Minnesota and Cedar Rapids, Iowa; | 1323 (670; 653) | 9.6 (SD 0.6) | 47% boys | Dietary and activity intervention<br>No active intervention<br>8 months<br>School + home + community |
| Griffin 2019<br>United Kingdom<br>(high income) | HDHK-UK (Healthy Dads, Healthy Kids, United Kingdom)<br>cluster RCT (father + ≥ 1 daughter)<br>Two urban local authority areas of the West Midlands | 61 (42; 19) | 7.7 (SD 2.1) | 100% boys | Dietary and activity intervention<br>Attention control<br>9 weeks<br>Community |
| Grydeland 2014<br>Norway<br>(high income) | HEIA (HEalth In Adolescents)<br>cluster RCT (school)<br>Thirty-seven schools in the largest towns/municipalities in seven counties surrounding Oslo | 2165 (784; 1381) | Intervention: 11.2 (SD 0.3)<br>Control: 11.2 (SD 0.3) | 51.4% boys | Dietary and activity intervention<br>No active intervention<br>20 months<br>School |
| Ha 2021<br>China<br>(upper middle income) | Active 1 + Fun<br>cluster RCT (parent + ≥ 1 child)<br>Families from eight local primary schools in Hong Kong | 160 (83 (at baseline); 77 (at baseline)) | 10 | 59.6 % boys | Activity intervention<br>No active intervention<br>6 months<br>School |
| Habib-Mourad 2014<br>Lebanon<br>(lower middle income) | Health-E-PALS<br>cluster RCT (school)<br>Eight private and public schools in Beirut | 374 (193; 181) | Intervention: 10.3 (SD 0.9)<br>Control: 10.1 (SD 1) | 54.5% boys | Dietary and activity intervention<br>No active intervention<br>12 weeks<br>School |
| Habib-Mourad 2020<br>Lebanon<br>(lower middle incoml) | Ajyal Salima Program<br>cluster RCT (school)<br>Private and public schools in Beirut | 1239 (698; 541) | 9.95 (SE 1.13) | 46.3% boys | Dietary and activity intervention<br>No active intervention<br>2 years<br>School |
| Haerens 2006<br>Belgium<br>(high income) | NR<br>cluster RCT (school)<br>Fifteen schools with technical and vocational education in West Flanders | 2840 (Intervention + parents involvement: 1226<br>Intervention only: 1006; 759) | 13.06 (SD 0.81) | 63.4% boys | Dietary and activity intervention<br>No active intervention<br>2 school years (9 months/year)<br>School |

| | | | | | |
|---|---|---|---|---|---|
| Hollis 2016<br>Australia<br>(high income) | PA4E (Physical Activity 4 Everyone)<br>cluster RCT (school)<br>Ten secondary schools in New South Wales | 1233 (NR; NR) | Median: 12 | Intervention: 48% boys<br>Control: 49% boys | Activity Intervention<br>No active intervention<br>7-8 school terms (19-24 months)<br>School + community + home |
| Hopper 2005<br>United states<br>(high income) | Family Fitness<br>cluster RCT (school)<br>Six elementary schools in Humboldt County, California | 238 (142 (at baseline); 96 (at baseline)) | 8.57 (SD 0.63) | 51% boys | Dietary and activity intervention<br>No active intervention<br>20 weeks<br>School |
| Ickovics 2019<br>United States<br>(high income) | School-Based Policies intervention<br>cluster RCT (2×2 factorial design) (school)<br>Twelve schools (kindergarten through eighth grade) in New Haven, Conecticut | 756 (Policy interventions related to nutrition: 202<br>Policy interventions related to physical activity: 176<br>Policy interventions related to nutrition and physical activity: 237 ; 141) | 10.9 (SD 0.62) | 46.2% boys | Dietary/activity/dietary and activity intervention (multi-arm)<br>Attention control<br>3 years<br>School |
| Jones 2015<br>Australia<br>(high income) | The Wollongong SPORT<br>RCT (individual)<br>Communities in low-income areas of Wollongong | 37 (19; 18) | Girls: 9.6 (SD 0.9); boys: 9.9 (SD 0.8) | 54% boys | Activity intervention<br>Attention control<br>7 months<br>School |
| Kain 2014<br>Chile<br>(high income) | NR<br>cluster RCT (school)<br>Nine primary public schools in Ñuñoa, a district of Santiago | 651 (651 (at baseline); 823 (at baseline)) | 6.6 (SD 1.07) | 53.4% boys | Dietary and activity intervention<br>No active intervention<br>12 months<br>School |
| Kennedy 2018<br>Australia<br>(high income) | Resistance Training for Teens<br>cluster RCT (school)<br>Sixteen government secondary schools in Hunter, Central Coast and Sydney regions of New South Wales | 607 (353; 254) | 14.1 (SD 0.5) | 49.9% boys; | Activity intervention<br>No active intervention<br>6 months<br>School + web |
| Kobel 2017<br>Germany<br>(high income) | Join the Healthy Boat (Baden-Wurttemberg Study)<br>cluster RCT (classroom)<br>Ninety-one primary schools of the state of Baden-Württemberg | 525 (318; 207) | 7.1 (SD 0.7) | 48.6% boys | Dietary and activity intervention<br>No active intervention<br>12 months<br>School |
| Kriemler 2010<br>Switzerland<br>(high income) | KISS<br>cluster RCT (school)<br>Fifteen schools in Aargau and Baselland provinces | 502 (297; 205) | 6.9 (SD 0.3) | 48.8% boys | Activity intervention<br>No active intervention<br>9 months<br>School |

| | | | | | |
|---|---|---|---|---|---|
| Kuroko 2020<br>New Zealand<br>(high income) | COOK (Create Our Own Kai)<br>RCT (individual)<br>Local educational facilities' teaching kitchens and homes in Dunedin | 164 (109; 55) | 13.6 (SD 0.8) | 35.6% boys | Dietary intervention<br>No active intervention<br>7 weeks<br>School (after school programme) + home + web |
| Lana 2014<br>Mexico (78% of participants); Spain (22% of participants)<br>(upper middle income (Mexico); high income (Spain)) | PREVENCANADOL program<br>RCT (individual)<br>Secondary education schools | 2001 (1014; 987) | NR | 45.2% boys | Dietary intervention<br>No active intervention<br>9 months<br>School + web |
| Levy 2012<br>Mexico<br>(upper middle income) | Nutrition on the go<br>cluster RCT (school)<br>Sixty schools in different municipalities of the State of Mexico | 1020 (510; 510) | % of age 10:<br>Intervention: 78.6%;<br>control: 75.3% | Intervention: 48.4% boys<br>Control: 50.3% boys | Dietary and activity intervention<br>No active intervention<br>6 months<br>School |
| Li 2010<br>China<br>(upper middle income) | Happy 10 program<br>cluster RCT (school)<br>Twenty primary schools from DongCheng and ChongWen disctricts (Beijing) | 4700 (2329; 2371) | 9.3 (SD 0.7) | 52.3% boys | Activity intervention<br>No active intervention<br>12 months<br>School |
| Li 2019<br>China<br>(upper middle income) | CHIRPY DRAGON<br>cluster RCT (school)<br>Fourty non-boarding, state-funded primary schools in traditional urban districts of Guangzhou | 1641 (832; 809) | Intervention: 6.15 (SD 0.36)<br>Control: 6.14 (SD 0.35) | 54.5% boys | Dietary and activity intervention<br>No active intervention<br>12 months<br>School |
| Liu 2019<br>China<br>(upper middle income) | NR<br>cluster RCT (school)<br>Twelve schools from Dongcheng District, a central districts in the east of Beijing | 1889 (930; 959) | 9 (SD 0.67) | 51.7% boys | Dietary and activity intervention<br>No active intervention<br>1 year<br>School |
| Liu 2022<br>China<br>(upper middle income) | DECIDE - Children (Diet, Exercise and Cardiovascular Health)<br>cluster RCT (school)<br>Twenty-four schools from three socioeconomically distinct Chinese areas: Beijing, Changzhi of Shanxi Province, and Urumuqi of Xinjiang Province | 1392 (705; 687) | Intervention: 9.6 (0.4)<br>Control: 9.6 (0.4) | 51.5% boys | Dietary and activity intervention<br>No active intervention<br>9 months<br>School |
| Lloyd 2018<br>United Kingdom<br>(high income) | HeLP (Healthy Lifestyles Programme)<br>cluster RCT (school)<br>Thirty-two state-run primary and junior schools in Devon and Plymouth | 1324 (676; 648) | 9.7 (SD 0.3) | 48.7% boys | Dietary and activity intervention<br>No active intervention |

| | | | | | |
|---|---|---|---|---|---|
| | | | | | 3 school terms (the spring and summer term of Year 5 and the autumn term of Year 6)<br>School |
| Lubans 2021<br>Australia<br>(high income) | B2L (Burn 2 Learn)<br>cluster RCT (school)<br>Twenty government secondary schools with senior school students in New South Wales | 670 (337; 333) | 16 (SD 0.4) | 55.4% boys | Activity intervention<br>No active intervention<br>20 weeks<br>School + web |
| Lynch 2016<br>United States<br>(high income) | Let's Go! 5-2-1-0<br>cluster RCT (classroom)<br>A local elementary school in Rochester, Minnesota | 51 (29; 22) | Intervention: 8 (IQR 7-8)<br>Control: 8 (IQR 7-9) | 51% boys | Dietary and activity intervention<br>No active intervention<br>4 months<br>School |
| Madsen 2013<br>United States<br>(high income) | Modified SCORES program<br>cluster RCT (school)<br>Seven schools in San Francisco, California | 156 (82; 74) | 9.8 (SD 0.6) | 60% boys | Activity intervention<br>No active intervention<br>2 school terms (12 weeks in the fall sessions and 12 weeks in the spring sessions)<br>School |
| Marcus 2009<br>Sweden<br>(high income) | STOPP<br>cluster RCT (school)<br>Ten primary schools in the Stockholm county area | 3135 (1670; 1465) | Intervention: 7.4 (SD 1.3)<br>Control: 7.5 (SD 1.3) | 50.8% boys | Dietary and activity intervention<br>No active intervention<br>1-4 years<br>School |
| Martinez-Vizcaino 2014<br>Spain<br>(high income) | MOVI-2<br>cluster RCT (school)<br>Twenty schools in 20 towns in the Province of Cuenca | 1592 (769; 823) | 9.5 (SD 0.5) | 48.6 boys | Activity intervention<br>No active intervention<br>9 months<br>School |
| Martinez-Vizcaino 2020<br>Spain<br>(high income) | MOVI-KIDS<br>cluster RCT (cross-over) (school)<br>Twenty-one pre-school and primary schools in Cuenca and Ciudad Real provinces in the Castilla-La Mancha region | 2407 (1299; 1108) | Intervention boys: 5.32 (SD 0.62); intervention girls: 5.38 (SD 0.64)<br>Control boys: 5.31 (SD 0.59); control girls: 5.39 (SD 0.62) | 50.1% boys | Activity intervention<br>No active intervention<br>8 months<br>School + home |
| Morgan 2011<br>Australia<br>(high income) | HDHK (Healthy Dads, Healthy Kids)<br>cluster RCT (father + ≥ 1 child)<br>Communities in Newcastle, New South Wales | 71 (39; 32) | 8.2 (SD2.0) | 53.5% boys | Dietary and activity intervention<br>No active intervention<br>3 months<br>Community |

| | | | | | |
|---|---|---|---|---|---|
| Morgan 2014<br>Australia<br>(high income) | HDHK (Healthy Dads, Healthy Kids)<br>cluster RCT (father + ≥ 1 child)<br>Communities in the Singleton and Maitland local government areas of the Hunter region | 132 (72; 60) | 8.1 (SD 2.1) | 55% boys | Dietary and activity intervention<br>No active intervention<br>7 weeks<br>Community |
| Morgan 2019<br>Australia<br>(high income) | DADEE<br>cluster RCT (family (father + ≥ 1 daughter) )<br>Communities in Newcastle, New South Wales | 153 (74; 79) | 7.7 (SD 1.8) | 100% girls | Activity intervention<br>No active intervention<br>8 weeks<br>Community |
| Muller 2019<br>South Africa<br>(upper middle income) | DASH (Disease, Activity and School children's Health)<br>cluster RCT (school)<br>Eight primary schools in Gqeberha (formerly Port Elizabeth) in the Eastern Cape province | 1009 (Physical activity (PA) intervention: 119 Physical activity + health and hygiene education (PA + HE) intervention: 181 Physical activity + health and hygiene education + nutritional education intervention (PA + HE + NU): 99 Health and hygiene education + nutritional education intervention (HE + NU): 140 ; No intervention: 470 (note: the analysis compared schools with physical activity intervention (n=337) vs schools without physical activity intervention (n=610))) | 10.0 (SD 0.9) | 51.1% boys | Activity intervention<br>No active intervention<br>1 school year (10 months; 2 x 10 week intervention periods)<br>School |
| Nemet 2011b<br>Israel<br>(high income) | NR<br>cluster RCT (school)<br>Schools in Central Israel | 342 (154; 188) | Intervention: 5.36 (SE 0.03)<br>Control: 5.4 (SE 0.04) | Intervention: 58% boys<br>Control: 55% boys | Dietary and activity intervention<br>No active intervention<br>1 school year<br>School |
| Newton 2014<br>United States<br>(high income) | Parent-Targeted Mobile Phone Intervention<br>RCT (parent/child dyad)<br>Communities in Baton Rouge, Louisiana | 27 (13; 14) | 8.7 (SD 1.4) | 44% boys | Activity intervention<br>Attention control<br>12 weeks<br>Home |

| | | | | | |
|---|---|---|---|---|---|
| Nicholl 2021<br>Australia<br>(high income) | Milky Way Study<br>RCT (individual)<br>Communities in Perth, Western Austraia | 49 (24; 25) | Intervention: 5.2 (SD 0.9)<br>Control: 5.2 (SD 0.9) | 53.1% boys | Dietary intervention<br>No active intervention<br>12.3 (SD 0.9) weeks (range: 11.5-15 weeks)<br>Home |
| O'Connor 2020<br>United States<br>(high income) | PSNS (Papa´s Saludables Nin˜os Saludables(<br>cluster RCT (father + ≤ 3 children)<br>One of the Texas Children's Health Plan (TCHP) Center for Children and Women clinics in Houston, Texas | 64 (31 (at baseline); 33 (at baseline)) | 8.5 (SD 2.12) | 43.8% boys | Dietary and activity intervention<br>No active intervention<br>10 weeks<br>Clinical setting |
| Pate 2005<br>United States<br>(high income) | LEAP (Lifestyle Education for Activity Program)<br>cluster RCT (school)<br>Twenty-four high schools in 14 counties in South Carolina | 2744 (1523; 1221) | Intervention: 13.6 (SD 0.6)<br>Control: 13.6 (SD 0.6) | 100% girls | Activity intervention<br>No active intervention<br>12 months<br>School + community + home |
| Pena 2021<br>Chile<br>(high income) | Juntos Santiago trial<br>cluster RCT (school)<br>Twenty-four public, private-subsidized, and private schools in the municipalities of Santiago and Estación Central in Santiago | 2022 (1611; 411) | Intervention: 11.1 (SD 0.8)<br>Control: 11.2 (SD 0.8) | 66.8% boys | Dietary and activity intervention<br>No active intervention<br>7 months<br>School |
| Pfeiffer 2019<br>United States<br>(high income) | Girls on the Move<br>cluster RCT (school)<br>Eight schools in Michigan | 1519 (753; 766) | Intervention: 12.05 (SD 0.99)<br>Control: 12.05 (SD 1.02) | 100% girls | Activity intervention<br>No active intervention<br>17 weeks<br>School + web |
| Puder 2011<br>Switzerland<br>(high income) | Ballabeina study<br>cluster RCT (classroom)<br>Forthy public preschool classes in the German (city of St Gallen) and the French (urban surroundings of Lausanne, canton Vaud speaking regions of Switzerland | 652 (342 (at baseline); 310 (at baseline)) | 5.1 (SD 0.7) | 50% boys | Dietary and activity intervention<br>No active intervention<br>10 months<br>School |
| Rosario 2012<br>Portugal<br>(high income) | NR<br>cluster RCT (school)<br>Seven Santos Simões public elementary public schools in Guimarães, Braga | 464 (233; 231) | 8.3 (SD 1.2) | 48.5% boys | Dietary and activity intervention<br>No active intervention<br>6 months<br>School |
| Rosenkranz 2010<br>United States<br>(high income) | SNAP (Scouting Nutrition & Activity Program)<br>cluster RCT (nested cohort design) (girl scout troops)<br>Communities in three Midwestern towns, Kansas | 76 (34; 42) | Intervention: 10.5 (SD 1.1)<br>Control: 10.5 (SD 1.3) | 100% girls | Dietary and activity intervention<br>No active intervention<br>4 months<br>Community |
| Rush 2012<br>New Zealand | Project Energize<br>cluster RCT (school) | 6456 (3263; 3193) | 5 and 10 | 50.4% boys | Dietary and activity intervention<br>No active intervention |

| | | | | | |
|---|---|---|---|---|---|
| (high income) | One-hundred four primary schools in the Waikato district | | | | 2 years<br>School |
| Sacchetti 2013<br>Italy<br>(high income) | NR<br>cluster RCT (classroom)<br>Twenty-six 3rd-grade classes of primary schools in a province of the Emilia Romagna region | 497 (247 (at baseline); 250 (at baseline)) | Range 8-9 | 51.5% boys | Activity intervention<br>No active intervention<br>2 years<br>School |
| Salmon 2022<br>Australia<br>(high income) | Transform-Us!<br>cluster RCT (2×2 factorial design) (school)<br>Twenty government, catholic and independent co-educational primary schools within 50 km of the Melbourne Central Business District | 593 (Physical activity intervention (PA-I): 161 Sedentary behaviour intervention (SB-I): 124 Physical activity + sedentary behaviour intervention (PA-I + SB-I): 159 (at baseline); 149 (at baseline)) | Range 8-9 | 44.2% boys | Activity intervention<br>No active intervention<br>30 months<br>School |
| Seguin-Fowler 2021<br>United states<br>(high income) | F3HK (Farm Fresh Foods for Healthy Kids)<br>RCT (caregiver/child dyad)<br>Farm communities in New York, North Carolina, Vermont, Washington | 305 (148; 157) | Intervention: 6.1 (SD 3)<br>Control: 6.2 (SD 3) | Intervention: 43.9% boys<br>Control: 51.6% boys | Dietary Intervention<br>No active intervention<br>2 years<br>Community + home |
| Sekhavat 2014<br>Canada<br>(high income) | NR<br>RCT (individual)<br>Undergraduate pediatric dentistry clinic at the University of Toronto's Faculty of Dentistry, Toronto | 168 (87; 81) | 8.97 (SD 1.52) | 52.4% boys | Dietary and activity intervention<br>No active intervention<br>5-10 minute counseling session during initial dental visit<br>Clinical setting |
| Sherwood 2019<br>United States<br>(high income) | Healthy Homes/Healthy Kids 5-10<br>RCT (parent/child dyad)<br>Community in the Greater Minneapolis-St. Paul area | 421 (212; 209) | 6.6 (SD 1.7) | 50.6% boys | Dietary and activity intervention<br>Attention control<br>12 months<br>Clinical setting + telehealth |
| Shomaker 2019<br>United States<br>(high income) | Learning to BREATHE<br>RCT (individual)<br>an outpatient, pediatric research laboratory at Colorado State University in Colorado | 54 (29; 25) | Intervention: 13.97 (SD 1.42)<br>Control: 14.49 (SD 1.72) | Intervention: 45% boys<br>Control: 44% boys | Dietary Intervention<br>Attention control<br>6 weeks<br>Home + community |
| Singh 2009<br>Netherlands<br>(high income) | DOiT (Dutch Obesity Intervention in Teenagers)<br>cluster RCT (school)<br>Eighteen prevocational secondary schools | 1108 (632; 476) | Intervention boys: 12.8 (SD 0.5); intervention girls: 12.6 (SD 0.5); | 49.55% boys | Dietary and activity intervention<br>No active intervention<br>8 months<br>School |

| | | | | | |
|---|---|---|---|---|---|
| | | | Control boys 12.9 (SD 0.5); control girls 12.7 (SD 0.5) | | |
| Smith 2014<br>Australia<br>(high income) | ATLAS (Active Teen Leaders Avoiding Screen-time)<br>cluster RCT (school)<br>Fourteen secondary schools in New South Wales | 361 (181; 180) | 12.7 (SD 0.5) | 100% boys | Activity intervention<br>No active intervention<br>8 months<br>School + web |
| Stettler 2015<br>United States<br>(high income) | Smart Steps<br>cluster RCT (clinical practice)<br>Clinical practices in Philadelphia | 173 (Smart Steps - beverage-only: 77<br>Smart Steps - multiple behavior: 63 ; 33) | Beverage-only intervention: 10.8 (SD 1.4)<br>Multiple behaviour intervention: 10.7 (SD 1.3)<br>Control: 10.8 (SD 1.4) | Beverage-only intervention: 46% boys<br>Multiple behaviour intervention: 43% boys<br>Control: 55% boys | Dietary/dietary and activity intervention (multi-arm)<br>Attention control<br>12 months<br>Clinical setting |
| Takacs 2020<br>Hungary<br>(high income) | NR<br>cluster RCT (classroom)<br>Two state-owned primary schools in Budaors-Pest County | 229 (117; 112) | 12.6 (SD 0.1) | 44.5% boys | Dietary intervention<br>No active intervention<br>9 months<br>School + after school programme + web |
| Tanskey 2017<br>United states<br>(high income) | FLEX (Fueling Learning through Exercise) Study<br>cluster RCT (school)<br>Sixteen schools in Massachusetts | 769 (100 Miles club: 261<br>Just Move: 249 (at baseline); 259 (at baseline)) | 8.7 (SD 0.7) | 44% boys | Activity intervention<br>No active intervention<br>2 years<br>School |
| TenHoor 2018<br>Netherlands<br>(high income) | Focus on Strength<br>cluster RCT (school)<br>Nine Dutch secondary schools | 695 (353; 342) | 12.97 (SD 0.54) | 50.36% boys | Activity intervention<br>No active intervention<br>12 months<br>School |
| Vizcaino 2008<br>Spain<br>(high income) | MOVI<br>cluster RCT (school)<br>Twenty schools in 20 towns in the Province of Cuenca | 1409 (691; 718) | Intervention boys: 9.4 (SD 0.7); intervention girls: 9.4 (SD 0.7);<br>Control boys : 9.5 (SD 0.7); | 50.6% boys | Activity intervention<br>No active intervention<br>24 weeks<br>School |

| | | | | | |
|---|---|---|---|---|---|
| | | | control girls:<br>9.4 (SD 0.6) | | |
| Weeks 2012<br>Australia<br>(high income) | POWER PE (Preventing Osteoporosis With Exercise Regimes in Physical Education)<br>RCT (individual)<br>One high school in the Gold Coast, Queensland | 99 (52; 47) | 13.8 (SD 0.4) | 46.5% boys | Activity intervention<br>No active intervention<br>8 months<br>School |
| Wendel 2016<br>United States<br>(high income) | NR<br>cluster RCT (classroom)<br>Twenty-four schools in Texas | 173 (101 (at baseline) ; 72 (at baseline) ) | 8.8 | 49.7% boys | Activity intervention<br>No active intervention<br>2 years<br>School |
| Wilksch 2015<br>Australia<br>(high income) | Life Smart<br>cluster RCT (classroom)<br>Twelve schools in South Australia, Victoria, Western Australia | 1441 (347; 473) | 13.21 (SD 0.68) | 36% boys | Dietary and activity intervention<br>No active intervention<br>5 weeks<br>School |
| Williamson 2012<br>United States<br>(high income) | Louisiana (LA) Health<br>cluster RCT (school)<br>Twenty three school systems in Louisiana | 1473 (Primary prevention intervention: 713<br>Primary + secondary prevention intervention: 760 ; 587) | 10.5 (SD 1.2 ) | 41.5% boys | Dietary and activity intervention<br>Attention control<br>28 months<br>School |
| Xu 2017<br>China<br>(upper middle income) | NISCOC (Nutrition-based Intervention Study on Childhood Obesity in China)<br>cluster RCT (school)<br>Thirty schools from Shanghai, Chongqing, Guangzhou, Jinan and Harbin | 7717 (3773; 3944) | 9 (SD 1.4) | Intervention: 50.9% boys<br>Control: 50.6% boys | Dietary and activity intervention<br>No active intervention<br>9 months<br>School |
| Yin 2012<br>United States<br>(high income) | Fitkid - Georgia Fitkid Project<br>cluster RCT (school)<br>Eighteen schools in Augusta, Richmond County, Georgia | 1187 (603; 584) | 8.7 (SD 0.5) | 47% boys | Activity intervention<br>No active intervention<br>3 years<br>School |
| Zhou 2019<br>China<br>(upper middle income) | CHAMPS (Childhood Health; Activity and Motor Performance Study)<br>cluster RCT (school)<br>Twelve middle schools in Beijing, Wuhu, Anhui Province, Weifang, Shandong Province | 758 (School physical education (SPE) intervention: 204<br>After school program (ASP) intervention: 200<br>School physical education Intervention + after school program intervention (SPE + ASP): 178; 176) | 12.66 (SD 0.56) | 53.4% boys | Dietary and activity intervention<br>No active intervention<br>8 months<br>School + after school programme |

https://doi.org/10.3390/nu12092687

https://res.mdpi.com/d_attachment/nutrients/nutrients-12-02687/article_deploy/nutrients-12-02687.pdf.

Haerens LD, B. Maes, L. Stevens, V. Cardon, G. De Bourdeaudhuij, I. Body mass effects of a physical activity and healthy food intervention in middle schools. Obesity. 2006;14(5):847-54.

Hollis JLS, R. Campbell, L. Morgan, P. J. Lubans, D. R. Nathan, N. Wolfenden, L. Okely, A. D. Davies, L. Williams, A. Cohen, K. E. Oldmeadow, C. Gillham, K. Wiggers, J. Effects of a 'school-based' physical activity intervention on adiposity in adolescents from economically disadvantaged communities: secondary outcomes of the 'Physical Activity 4 Everyone' RCT. International Journal of Obesity. 2016;40(10):1486-93.

Hopper CA, Munoz KD, Gruber MB, Nguyen KP. The Effects of a Family Fitness Program on the Physical Activity and Nutrition Behaviors of Third-Grade Children. Research Quarterly for Exercise and Sport. 2005;76(2):130-39.

Ickovics JR, Duffany KO, Shebl FM, Peters SM, Read MA, Gilstad-Hayden KR, et al. Implementing School-Based Policies to Prevent Obesity: Cluster Randomized Trial. American Journal of Preventive Medicine. 2019;56(1):e1-11.

Jones RA, Kelly J, Cliff DP, Batterham M, Okely AD. Acceptability and Potential Efficacy of Single-Sex After-School Activity Programs for Overweight and At-Risk Children: The Wollongong SPORT RCT. Pediatric Exercise Science. 2015;27(4):535-45.

Kain J, Concha F, Moreno L, Leyton B. School-based obesity prevention intervention in Chilean children: effective in controlling, but not reducing obesity. Journal of Obesity. 2014;2014:618293.

Kennedy SG, Smith JJ, Morgan PJ, Peralta LR, Hilland TA, Eather N, et al. Implementing Resistance Training in Secondary Schools: A Cluster Randomized Controlled Trial. Med Sci Sports Exerc. 2018;50(1):62-72.

Kobel S, Lammle C, Wartha O, Kesztyus D, Wirt T, Steinacker JM. Effects of a Randomised Controlled School-Based Health Promotion Intervention on Obesity Related Behavioural Outcomes of Children with Migration Background. Journal of Immigrant and Minority Health. 2017;19(2):254-62.

Kriemler S, Zahner L, Schindler C, Meyer U, Hartmann T, Hebestreit H, et al. Effect of school based physical activity programme (KISS) on fitness and adiposity in primary schoolchildren: cluster randomised controlled trial. BMJ (Clinical research ed). 2010;340:c785.

Kuroko S, Black K, Chryssidis T, Finigan R, Hann C, Haszard J, et al. Create Our Own Kai: a Randomised Control Trial of a Cooking Intervention with Group Interview Insights into Adolescent Cooking Behaviours. Nutrients. 2020;12(3).

Lana AF-O, G. Lopez, M. L. Impact of a web-based intervention supplemented with text messages to improve cancer prevention behaviors among adolescents: results from a randomized controlled trial. Preventive medicine. 2014;59:54-9.

Levy S, Morales Ruan T, Amaya Castellanos C, Salazar Coronel C, Jimenez Aguilar A, Mendez Gomez A, et al. Effectiveness of a diet and physical activity promotion strategy on the prevention of obesity in Mexican school children. BMC Public Health. 2012;12:152.

Li B, Pallan M, Liu WJ, Hemming K, Frew E, Lin R, et al. The CHIRPY DRAGON intervention in preventing obesity in Chinese primary-school--aged children: A cluster-randomised controlled trial. PLoS Medicine. 2019;16(11):e1002971.

Li YP, Hu XQ, Schouten EG, Liu AL, Du SM, Li LZ, et al. Report on childhood obesity in China (8): effects and sustainability of physical activity intervention on body composition of Chinese youth. Biomedical and Environmental Sciences. 2010;23(3):180-7.

Liu Z, Gao P, Gao AY, Lin Y, Feng XX, Zhang F, et al. Effectiveness of a Multifaceted Intervention for Prevention of Obesity in Primary School Children in China: A Cluster Randomized Clinical Trial. JAMA Pediatrics. 2022;176(1):e214375.

Liu Z, Li Q, Maddison R, Ni Mhurchu C, Jiang Y, Wei DM, et al. A School-Based Comprehensive Intervention for Childhood Obesity in China: A Cluster Randomized Controlled Trial. Childhood Obesity. 2019;15(2):105-15.

Lloyd J, Creanor S, Logan S, Green C, Dean SG, Hillsdon M, et al. Effectiveness of the Healthy Lifestyles Programme (HeLP) to prevent obesity in UK primary-school children: a cluster randomised controlled trial. The Lancet Child and Adolescent Health. 2018;2(1):35-45.

Lubans DR, Smith JJ, Eather N, Leahy AA, Morgan PJ, Lonsdale C, et al. Time-efficient intervention to improve older adolescents' cardiorespiratory fitness: findings from the 'Burn 2 Learn' cluster randomised controlled trial. British Journal of Sports Medicine. 2021;55(13):751.

Lynch BA, Gentile N, Maxson J, Quigg S, Swenson L, Kaufman T. Elementary School-Based Obesity Intervention Using an Educational Curriculum. Journal of Primary Care & Community Health. 2016;7(4):265-71.

Madsen K, Thompson H, Adkins A, Crawford Y. School-community partnerships: a cluster-randomized trial of an after-school soccer program. JAMA Pediatrics. 2013;167(4):321-26.

Marcus C, Nyberg G, Nordenfelt A, Karpmyr M, Kowalski J, Ekelund U. A 4-year, cluster-randomized, controlled childhood obesity prevention study: STOPP. International Journal of Obesity2009. p. 408-17.

Martinez-Vizcaino V, Pozuelo-Carrascosa DP, Garcia-Prieto JC, Cavero-Redondo I, Solera-Martinez M, Garrido-Miguel M, et al. Effectiveness of a school-based physical activity intervention on adiposity, fitness and blood pressure: MOVI-KIDS study. British Journal of Sports Medicine. 2020;54(5):279-85.

Martinez-Vizcaino VS-L, M. Notario-Pacheco, B. Salcedo-Aguilar, F. Solera-Martínez, M. Franquelo-Morales, P. L, et a. Gender differences on effectiveness of a school-based physical activity intervention for reducing cardiometabolic risk: A cluster randomized trial. International Journal of Behavioral Nutrition and Physical Activity. 2014;11:154.

Morgan PJ, Collins CE, Plotnikoff RC, Callister R, Burrows T, Fletcher R, et al. The 'Healthy Dads, Healthy Kids' community randomized controlled trial: a community-based healthy lifestyle program for fathers and their children. Preventive Medicine. 2014;61:90-9.

Morgan PJ, Young MD, Barnes AT, Eather N, Pollock ER, Lubans DR. Engaging Fathers to Increase Physical Activity in Girls: The "Dads And Daughters Exercising and Empowered" (DADEE) Randomized Controlled Trial. Annals of Behavioral Medicine. 2019;53(1):39-52.

Morgan PJL, D. R. Callister, R. Okely, A. D. Burrows, T. L. Fletcher, R. Collins, C. E. The 'Healthy Dads, Healthy Kids' randomized controlled trial: efficacy of a healthy lifestyle program for overweight fathers and their children. International journal of obesity. 2011;35(3):436-47.

Muller I, Schindler C, Adams L, Endes K, Gall S, Gerber M, et al. Effect of a Multidimensional Physical Activity Intervention on Body Mass Index, Skinfolds and Fitness in South African Children: Results from a Cluster-Randomised Controlled Trial. International Journal of Environmental Research & Public Health [Electronic Resource]. 2019;16(2):15.

Nemet D, Geva D, Pantanowitz M, Igbaria N, Meckel Y, Eliakim A. Health promotion intervention in Arab-Israeli kindergarten children. Journal of Pediatric Endocrinology. 2011;24(11-12):1001-7.

Newton RLJ, Marker AM, Allen HR, Machtmes R, Han H, Johnson WD, et al. Parent-Targeted Mobile Phone Intervention to Increase Physical Activity in Sedentary Children: Randomized Pilot Trial. Journal of Medical Internet Research mHealth uHealth. 2014;2(4):e48.

Nicholl A, Deering KE, Evelegh K, Lyons-Wall P, Lawrence D, Mori TA, et al. Whole-fat dairy products do not adversely affect adiposity or cardiometabolic risk factors in children in the Milky Way Study: a double-blind randomized controlled pilot study. American Journal of Clinical Nutrition. 2021;114(6):2025-42.

O'Connor TM, Beltran A, Musaad S, Perez O, Flores A, Galdamez-Calderon E, et al. Feasibility of Targeting Hispanic Fathers and Children in an Obesity Intervention: Papás Saludables Niños Saludables. Childhood Obesity. 2020;16(6):379-92.

Pate RRW, D. S. Saunders, R. P. Felton, G. Dishman, R. K. Dowda, M. Promotion of physical activity among high-school girls: a randomized controlled trial. American Journal of Public Health. 2005;95(9):1582-7.

Pena S, Carranza M, Cuadrado C, Parra DC, Villalobos Dintrans P, Castillo C, et al. Effectiveness of a Gamification Strategy to Prevent Childhood Obesity in Schools: A Cluster Controlled Trial. Obesity. 2021;17:17.

Pfeiffer KA, Robbins LB, Ling J, Sharma DB, Dalimonte-Merckling DM, Voskuil VR, et al. Effects of the Girls on the Move randomized trial on adiposity and aerobic performance (secondary outcomes) in low-income adolescent girls. Pediatric Obesity. 2019:e12559.

Puder JJ, Marques-Vidal P, Schindler C, Zahner L, Niederer I, Bürgi F, et al. Effect of multidimensional lifestyle intervention on fitness and adiposity in predominantly migrant preschool children (Ballabeina): cluster randomised controlled trial. BMJ. 2011;343:d6195.

Rosario R, Oliveira B, Araujo A, Lopes O, Padrao P, Moreira A, et al. The impact of an intervention taught by trained teachers on childhood overweight. International Journal of Environmental Research & Public Health. 2012;9(4):1355-67.

Rosenkranz RRB, T. K. Dzewaltowski, D. A. A group-randomized controlled trial for health promotion in Girl Scouts: healthier troops in a SNAP (Scouting Nutrition & Activity Program). BMC Public Health. 2010;10:81.

Rush E, Reed P, McLennan S, Coppinger T, Simmons D, Graham D. A school-based obesity control programme: Project Energize. Two-year outcomes. British Journal of Nutrition. 2012;107(4):581-7.

Sacchetti R, Ceciliani A, Garulli A, Dallolio L, Beltrami P, Leoni E. Effects of a 2-year school-based intervention of enhanced physical education in the primary school. Journal of School Health. 2013;83(9):639-46.

Salmon J, Arundell L, Cerin E, Ridgers ND, Hesketh KD, Daly RM, et al. Transform-Us! cluster RCT: 18-month and 30-month effects on children's physical activity, sedentary time and cardiometabolic risk markers. British Journal of Sports Medicine. 2023;57(5):311-19.

Seguin-Fowler RA, Hanson KL, Jilcott Pitts SB, Kolodinsky J, Sitaker M, Ammerman AS, et al. Community supported agriculture plus nutrition education improves skills, self-efficacy, and eating behaviors among low-income caregivers but not their children: a randomized controlled trial. International Journal of Behavioral Nutrition and Physical Activity. 2021;18(1):112.

Sekhavat A. Efficacy of nutrition and physical activity counseling for pre-adolescent children in a dental setting : a randomized clinical trial 2014.

Sherwood NE, Levy RL, Seburg EM, Crain AL, Langer SL, JaKa MM, et al. The Healthy Homes/Healthy Kids 5-10 Obesity Prevention Trial: 12 and 24-month outcomes. Pediatric Obesity. 2019;14(8):e12523.

Shomaker LB, Berman Z, Burke M, Annameier SK, Pivarunas B, Sanchez N, et al. Mindfulness-based group intervention in adolescents at-risk for excess weight gain: A randomized controlled pilot study. Appetite. 2019;140:213-22.

Singh ASCAP, M. J. Brug, J. Van Mechelen, W. Dutch obesity intervention in teenagers: effectiveness of a school-based program on body composition and behavior. Archives of Pediatrics & Adolescent Medicine2009. p. 309-17.

Smith JJM, P. J. Plotnikoff, R. C. Dally, K. A. Salmon, J. Okely, A. D. Finn, T. L. Lubans, D. R. Smart-phone obesity prevention trial for adolescent boys in low-income communities: the ATLAS RCT. Pediatrics. 2014;134(3):e723-e31.

Stettler N, Wrotniak BH, Hill DL, Kumanyika SK, Xanthopoulos MS, Nihtianova S, et al. Prevention of excess weight gain in paediatric primary care: beverages only or multiple lifestyle factors. The Smart Step Study, a cluster-randomized clinical trial. Pediatric Obesity. 2015;10(4):267-74.

Takacs H, Martos E, Kovacs VA. Effects of a practice-focused nutrition intervention in Hungarian adolescents. Annali dell'Istituto superiore di sanita. 2020;56(1):99-106.

Tanskey LA. An Investigation of Summer Weight Change in Elementary School Students [Ph.D.]. Ann Arbor: Tufts University, Gerald J. and Dorothy R. Friedman School of Nutrition Science and Policy; 2017.

Ten Hoor GA, Rutten GM, Van Breukelen GJP, Kok G, Ruiter RAC, Meijer K, et al. Strength exercises during physical education classes in secondary schools improve body composition: a cluster randomized controlled trial. International Journal of Behavioral Nutrition & Physical Activity. 2018;15(1):92.

Vizcaíno VM, Aguilar FS, Gutiérrez RF, Martínez MS, López MS, Martínez SS, et al. Assessment of an after-school physical activity program to prevent obesity among 9- to 10-year-oldchildren: a cluster randomized trial. International Journal of Obesity. 2008;32:12-22.

Weeks BKB, B. R. Twice-weekly, in-school jumping improves lean mass, particularly in adolescent boys. Pediatric Obesity. 2012;7(3):196-204.

Wendel ML, Benden ME, Zhao H, Jeffrey C. Stand-Biased Versus Seated Classrooms and Childhood Obesity: A Randomized Experiment in Texas. American Journal of Public Health. 2016;106(10):1849-54.

Wilksch SMP, S. J. Byrne, S. M. Austin, S. B. McLean, S. A. Thompson, K. M. Dorairaj, K. Wade, T. D. Prevention across the spectrum: A randomized controlled trial of three programs to reduce risk factors for both eating disorders and obesity. Psychological Medicine. 2015;45(9):1811-23.

Williamson DA, Champagne CM, Harsha DW, Han H, Martin CK, Newton RlJ, et al. Effect of an environmental school-based obesity prevention program on changes in body fat and body weight: a randomized trial. Obesity. 2012;20(8):1653-61.

Xu H, Li Y, Zhang Q, Hu XL, Liu A, Du S, et al. Comprehensive school-based intervention to control overweight and obesity in China: a cluster randomized controlled trial. Asia Pacific Journal of Clinical Nutrition. 2017;26(6):1139-51.

Yin Z, Yin Z. The impact of a 3-year after-school obesity prevention program in elementary school children. Childhood Obesity. 2012;8(1):60-70.

Zhou Z, Li S, Yin J, Fu Q, Ren H, Jin T, et al. Impact on Physical Fitness of the Chinese CHAMPS: A Clustered Randomized Controlled Trial. International Journal of Environmental Research & Public Health [Electronic Resource]. 2019;16(22):11.

Supplementary Table 2: Coding of interventions against characteristics suggested to reduce inequalities or drive intervention-generated inequalities

| Study ID | **Targeted (and inequity factor(s) targeted) or Universal**[a] (coding taken from Cochrane reviews) | **Socioecological model (SEM) domain (main)**[b] (Public policy; Society; Community; Organization; Interpersonal; Individual)[2] | **Step on Nuffield Ladder**[c] (Eliminate; Restrict; Guide or Enable choice; Provide information) | **Intervention included (Yes/No) an explicit component aiming to change the physical environment**[d] **for the child** (coding taken from Cochrane reviews) | **Degree of public involvement (and inequity factor(s)**[e]**:** User-controlled; Co-production; Consultation including child PPI; Consultation; none reported. |
|---|---|---|---|---|---|
| Adab 2018 | **Universal** | **Organizational** School (+ a community and a family opportunity for involvement, via the school) | Guide/Enable | No | **Consultation** |
| Barbosa Filho 2017 | **Targeted (low SES)** (All schools were in areas with a low Human Development Index) | **Organizational** School | Guide/Enable | **Yes** (sports/games equipment) | **Consultation** |
| Barnes 2015 | **Universal** (for girls only) | **Community** Community | Guide/Enable | No | None reported |
| Bogart 2016 | **Targeted (low SES)** (All schools had >50% of students eligible for the National Free School Lunch Program) | **Organizational** School + home | Guide/Enable | **Yes** (school-wide changes including free chilled filtered water) | **Consultation** (used a community-based participatory research approach; intervention included student peer-leaders) |
| Breheny 2020 | **Universal** | **Organizational** School | Guide/Enable | **Yes** (identification of walking/running paths for Daily Mile) | None reported (However, this intervention is a policy recommendation) |
| Brown 2013 | **Targeted (Race)** (American Indian reservations) | **Community** Community | Guide/Enable | No | **Consultation** (used a community-based participatory research approach) |
| Chai 2019 | **Universal** | **Interpersonal** Home + Telehealth | Guide/Enable | No | None reported |
| Damsgaard 2014 | **Universal** | **Organizational** School | **Restrict** | **Yes** (change in school food offering) | None reported |
| Dewar 2013 | **Targeted (low SES)** | **Organizational** School | Guide/Enable | No | None reported |

| | | | | | |
|---|---|---|---|---|---|
| | (Only schools in lower 50% of the Socio-Economic Indexes for Areas index)<br>(girls only) | | | | |
| Drummy 2016 | **Universal** | **Organizational**<br>School | Guide/Enable | No | None reported |
| Duncan 2019 | **Universal** | **Organizational**<br>School (and home) | Guide/Enable | No | **Consultation**<br>(intersectoral steering group) |
| Ebbeling 2006 | **Universal** | **Interpersonal**<br>Home + telehealth | Guide/Enable<br>(significant incentive) | **Yes**<br>(home deliveries of free noncaloric beverages to displace SSBs) | None reported |
| El Ansari 2010 | **Universal** | **Organizational**<br>School (after school programme) | Guide/Enable | No | None reported |
| Fairclough 2013 | **Targeted (low SES)**<br>Conducted in an area of high deprivation and, within it, relatively high and low SES schools were randomized. | **Organizational**<br>School | Guide/Enable | No | **Consultation including child PPI** |
| Farmer 2017 | **Universal** | **Organizational**<br>School | Guide/Enable | **Yes**<br>(change in playground environment) | **Consultation**<br>(to develop a playground action plan – *child involvement not explicitly mentioned*) |
| Fulkerson 2015 | **Universal** | **Interpersonal**<br>Home + community | Guide/Enable | No | **Consultation** |
| Fulkerson 2022 | **Universal**<br>(in rural communities) | **Interpersonal**<br>Home + community | Guide/Enable | No | **Consultation** |
| Gentile 2009 | **Universal** | **Organizational**<br>School (+ home + community) | Guide/Enable | **Yes** | None reported |
| Griffin 2019 | **Targeted (SES)**<br>(Areas selected were ranked in the most deprived 20% in the UK) | **Community**<br>Community | Guide/Enable | No | **None reported**<br>(although initial intervention was culturally adapted and further adapted after pilot study using parents' feedback) |
| Grydeland 2014 | **Universal** | **Organizational**<br>School | Guide/Enable | **Yes** | **Consultation including child PPI** |

| | | | | | |
|---|---|---|---|---|---|
| | | | | (various including school policy, free food, activity equipment) | (through focus groups with children and parents). |
| Ha 2021 | **Universal** | **Organizational**<br>School (+ home) | Guide/Enable | **Yes**<br>(sports equipment) | None reported |
| Habib-Mourad 2014 | **Universal** | **Organizational**<br>School | Guide/Enable | **Yes**<br>(school policy, including promotion (and availability) of healthy foods & beverages available/allowed in school shop and lunch boxes, respectively) | None reported |
| Habib-Mourad 2020 | **Universal** | **Organizational**<br>School | Guide/Enable | **Yes**<br>(school policy, including promotion (and availability) of healthy foods & beverages available/allowed in school shop and lunch boxes, respectively) | None reported<br>(used a whole school approach) |
| Haerens 2006 | **Universal**<br>(Only schools offering technical or vocational training) | **Organizational**<br>School | Guide/Enable | **Yes**<br>(physical activity environment; extra sports material; compulsory 10 min cycle) | None reported |
| Hollis 2016 | **Targeted (low SES)**<br>schools from socio-economically disadvantaged communities (in lower 50% in district). | **Organizational**<br>School + community + home | Guide/Enable | **Yes**<br>(physical activity environment; school policy; active school breaks; provision of equipment) | None reported |
| Hopper 2005 | **Universal** | **Organizational**<br>School | Guide/Enable | No | None reported |
| Ickovics 2019 | **Targeted (low SES)**<br>Eligibility for free lunches was particularly high overall in the area, exceeding 60% in all schools (mean=71.4%) | **Organizational**<br>School (+ home) | Guide/Enable | **Yes**<br>(school policy, changes relating to diet and physical activity) | None reported<br>(used a whole school approach) |
| Jones 2015 | **Targeted (low SES)**<br>Children were recruited from low-income areas (single-sex intervention) | **Organizational**<br>School (+ home) | Guide/Enable | **Yes** (school physical environment) | None reported |

| | | | | | |
|---|---|---|---|---|---|
| Kain 2014 | **Targeted (low SES)**<br>Schools in low-income areas | **Organizational**<br>School | Guide/Enable | No | None reported |
| Kennedy 2018 | **Universal** | **Organizational**<br>School + web | Guide/Enable | **Yes** (physical activity environment) | None reported |
| Kobel 2017 | **Universal** | **Organizational**<br>School | Guide/Enable | No | None reported |
| Kriemler 2010 | **Universal** | **Organizational**<br>School | Guide/Enable | **Yes** (physical activity environment) | None reported |
| Kuroko 2020 | **Universal** | **Organizational**<br>School (after school programme, + home + web) | Guide/Enable | No | None reported<br>(although initial intervention was culturally adapted and further adapted after pilot study using participant feedback) |
| Lana 2014 | **Universal** | **Organizational**<br>School + web | Guide/Enable | No<br>(although it did impact online social environment) | None reported<br>(although does mention a focus group with students was conducted to assess their preferences regarding website design) |
| Levy 2012 | **Universal** | **Organizational**<br>School | Guide/Enable | **Yes**<br>(diet and water) | **Consultation**<br>(although the campaign was developed with the 'school population', involvement with school children was not specifically mentioned) |
| Li 2010 | **Universal** | **Organizational**<br>School | Guide/Enable | No | None reported |
| Li 2019 | **Universal** | **Organizational**<br>School | Guide/Enable | **Yes**<br>(school lunches; school policy on physical activity) | **Consultation**<br>(process evaluation did include school children) |
| Liu 2019 | **Universal** | **Organizational**<br>School | **Restrict** | **Yes**<br>(school policy included SSSBs, unhealthy snacks, electronic devices) | **Consultation including child PPI** |
| Liu 2022 | **Universal** | **Organizational**<br>School | **Restrict** | **Yes** | **Consultation including child PPI** |

| | | | | | |
|---|---|---|---|---|---|
| | | | | (school policy included not selling, eating or buying SSSBs or unhealthy snacks; weighed children weekly) | |
| Lloyd 2018 | **Universal** | **Organizational**<br>School | Guide/Enable | No | **Consultation including child PPI** |
| Lubans 2021 | **Universal** | **Organizational**<br>School + web | Guide/Enable | **Yes**<br>(focussed on compulsory HIIT activity breaks as part of school policy) | None reported |
| Lynch 2016 | **Universal** | **Organizational**<br>School | Guide/Enable | No | None reported<br>(very limited consultation) |
| Madsen 2013 | **Targeted (low SES)**<br>(the after-school program preferentially enrolled students who qualify for free or reduced-price meals) | **Organizational**<br>School<br>(community-based after-school program) | Guide/Enable | No | None reported<br>(although the Company who produce the intervention had refined it based on 20 years of experience working with low SES schools) |
| Marcus 2009 | **Universal** | **Organizational**<br>School | **Restrict**<br>(all sweets & SSBs were eliminated from school) | **Yes**<br>(all sweets & SSBs were eliminated from school) | None reported |
| Martinez-Vizcaino 2014 | **Universal** | **Organizational**<br>School | Guide/Enable | No | None reported |
| Martinez-Vizcaino 2020 | **Universal** | **Organizational**<br>School + home | Guide/Enable | **Yes**<br>(equipment in the playground) | None reported |
| Morgan 2011 | **Universal** | **Community**<br>Community | Guide/Enable | No | None reported |
| Morgan 2014 | **Targeted (Occupation)**<br>The study setting is two local government areas with high rates of mining and shift work-based employment | **Community**<br>Community | Guide/Enable | No | None reported |
| Morgan 2019 | **Universal** | **Community**<br>Community | Guide/Enable | **Yes** | None reported |

| | | | | | |
|---|---|---|---|---|---|
| | | | | (home environment; provision of equipment) | (although the intervention was 'informed by an extensive program of<br>qualitative and quantitative research', no evidence of consultation could be found) |
| Muller 2019 | **Universal** | **Organizational**<br>School | Guide/Enable | **Yes**<br>(school infrastructure enhancements e.g. activity stations) | **Consultation including child PPI**<br>(no details provided, only 'developed in collaboration with education authorities, teachers and students from the participating schools) |
| Nemet 2011b | **Targeted (low SES)**<br>(kindergartens from low socioeconomic status communities) | **Organizational**<br>School (+ home) | Guide/Enable | No | **Consultation**<br>(limited in nature and detail) |
| Newton 2014 | **Universal** | **Interpersonal**<br>Home | Guide/Enable | No | None reported |
| Nicholl 2021 | **Universal** | **Interpersonal**<br>Home | Guide/Enable | **Yes**<br>(low fat dairy products were provided free of charge | **Consultation including child PPI** |
| O'Connor 2020 | **Targeted (Race)**<br>Children of fathers who self-identified as Hispanic or Latino | **Organizational**<br>Clinical setting | Guide/Enable | **Yes**<br>(home environment; free culturally adapted equipment) | **Consultation including child PPI**<br>(informed by a family panel *which we assume includes children*) |
| Pate 2005 | **Universal**<br>(only girls) | **Organizational**<br>School + community + home | Guide/Enable | NO<br>(changed environment but not physical environment) | None reported |
| Pena 2021 | **Universal** | **Organizational**<br>School | Guide/Enable | No | **Consultation including child PPI**<br>(intervention was codesigned with the children) |
| Pfeiffer 2019 | **Targeted (low SES)**<br>(83% of children qualified for free/reduced-price school lunches) (girls only) | **Organizational**<br>School + web | Guide/Enable | No | None reported |

| | | | | | |
|---|---|---|---|---|---|
| Puder 2011 | **Targeted (Race)**<br>(72% of children with at least one parent with migrant status) | **Organizational**<br>School | Guide/Enable | **Yes**<br>(built environment) | **Consultation** |
| Rosario 2012 | **Universal** | **Organizational**<br>School | Guide/Enable | No | None reported |
| Rosenkranz 2010 | **Universal**<br>(girls only) | **Community**<br>Community | Guide/Enable (although consumption of SSBs, sweets and TV watching during girl scouts meetings was prohibited) | No | None reported |
| Rush 2012 | **Universal** | **Organizational**<br>School | Guide/Enable (although pies and cookies were removed from canteen options and replaced with filled rolls, fruit and low-fat yogurt) | **Yes**<br>(canteen 'makeovers') | None reported (intervention development and delivery was contracted to a company) |
| Sacchetti 2013 | **Universal** | **Organizational**<br>School | Guide/Enable | No | None reported |
| Salmon 2022 | **Universal** | **Organizational**<br>School (+ home) | Guide/Enable | **Yes**<br>(e.g. physical activity equipment; school policy) | **Consultation**<br>(and learning from previous trials) |
| Seguin-Fowler 2021 | **Targeted (low SES)**<br>(households were eligible if they met guidelines for low income (<185% federal poverty level) | **Community**<br>Community + home | Guide/Enable | **Yes**<br>(cost-offset (half-price) community supported agriculture | **Consultation including child PPI**<br>(and learning from previous trials) |
| Sekhavat 2014 | **Universal** | **Organizational**<br>Clinical setting (dental) | Provide information | No | None reported |
| Sherwood 2019 | **Universal** | **Organizational**<br>Clinical setting + telehealth | Guide/Enable | No | None reported |
| Shomaker 2019 | **Universal** | **Interpersonal**<br>Home (+ community) | Guide/Enable | **Yes**<br>(given meditation equipment) | None reported |
| Singh 2009 | **Universal** | **Organizational**<br>School | Guide/Enable | **Yes**<br>(e.g. advice for schools on changes in and around school cafeterias) | None reported (although children were involved in logo design and |

| | | | | | |
|---|---|---|---|---|---|
| | | | | | appearance of intervention materials) |
| Smith 2014 | **Targeted (low SES)** (Only schools in lowest 50% of SES areas were eligible) | **Organizational** School + web | Guide/Enable | **Yes** (equipment was provided to some schools depending on requirements) | None reported |
| Stettler 2015 | **Universal** | **Organizational** Clinical setting | Guide/Enable | No | None reported |
| Takacs 2020 | **Universal** | **Organizational** School (after school programme +web) | Guide/Enable | No | None reported |
| Tanskey 2017 | **Targeted (low SES)** Eligible schools had >40% of students qualifying for free or reduced-price lunch and/or had >40% non-Caucasian students | **Organizational** School | Guide/Enable | No (although no mention of school policy, context suggests a 'whole school approach') | None reported (although interventions were those already used and identified for good practize by a nationwide initiative through a contest) |
| TenHoor 2018 | **Universal** | **Organizational** School | Guide/Enable | No (although encouraged school management to purchase additional equipment if provision not adequate). | **Consultation** (teachers were involved in choosing strength exercises) |
| Vizcaino 2008 | **Universal** | **Organizational** School (after school) | Guide/Enable | No (although equipment use was donated to schools at the end of the project) | None reported |
| Weeks 2012 | **Universal** | **Organizational** School | Guide/Enable | No | None reported |
| Wendel 2016 | **Universal** | **Organizational** School | Guide/Enable | **Yes** (stand-biased desks) | None reported (although based on pilot work) |
| Wilksch 2015 | **Universal** | **Organizational** School | Guide/Enable | No | None reported (although interventions chosen had previously been evaluated and this may have included PPI) |
| Williamson 2012 | **Targeted (Place - rural)** (Majority of children were from low income African-American families) | **Organizational** School | Guide/Enable | **Yes** (school environment, various including cafeteria food service and range of options in vending machines) | None reported (although based on their previous research projects) |
| Xu 2017 | **Universal** | **Organizational** School | Guide/Enable | **Yes** (including school lunch cafeteria menu) | None reported |

| | | | | | |
|---|---|---|---|---|---|
| Yin 2012 | **Targeted (SES)**<br>(in a school district where 65% of children qualified for reduced price or free school lunches and 66% African American) | **Organizational**<br>School (after school) | Guide/Enable | No | **Consultation**<br>(intervention was designed in collaboration with school officials and teachers) |
| Zhou 2019 | **Universal** | **Organizational**<br>School (+ after school programme) | Guide/Enable | **Yes**<br>(school policies; provision of equipment) | None reported |

[a]Trials were categorised as Universal for this exploratory analysis if the eligibility criteria for schools, communities or individual children and young people was not related to one of the eight inequity factors (PROGRESS factors) described in this paper. For example, studies that only selected participants who were above a certain BMI percentile, or reported a certain level of physical activity or an average consumption of a particular food or beverage type above or below a certain cut off, or who had a parent who was living with overweight or obesity, were categorised as Universal. Many of the studies in this analysis reported focussing their recruitment generally towards schools or communities within relatively low-income areas/districts, but these were classified as Universal interventions unless there was clear evidence (detail) that specific recruitment criteria relating to SES was involved.
[b]If an intervention was implemented in more than one domain of the SEM, the most upstream domain is listed.
[c]If an intervention was implemented on more than one step of the Nuffield intervention ladder, the highest step is listed.
[d]The physical environment is also sometimes termed as the built environment or the structural environment.
[e]Based on information available in trial reports (including protocols) only.

Supplementary Table 3: Risk of bias assessments for included studies

| | Risk of bias 2 (RoB 2) assessment | | | | | | Extra considerations | |
|---|---|---|---|---|---|---|---|---|
| **Study ID** | **D1. Risk of bias arising from the randomization process** | **D2. Risk of bias due to deviations from the intended interventions** | **D3. Risk of bias due to missing data** | **D4. Risk of bias in measurement of the outcome** | **D5. Risk of bias in selection of the reported result** | **Overall risk of bias judgement** | **Risk of bias due to missing subgroup analysis data (extracted and/or provided by trialists)** | **Risk of bias in selection of the subgroup analysis result** |
| **Adab 2018** | Low | Low | Some concerns | Low | Low | Some concerns | Low (Race/Ethnicity/Culture/Language; Gender/Sex; Socioeconomic status; Place of Residence)<br>Some concerns (Occupation; Religion; Education; Social capital) | Low |
| **Barbosa Filho 2017**[a] | High | Low | n/a | Low | n/a | High | Low | Low |
| **Barnes 2015** | Low | Low | Low | Low | Some concerns | Some concerns | Low | Low |
| **Bogart 2016** | High | Low | High | Low | Some concerns | High | Low (Race/Ethnicity/Culture/Language; Gender/Sex)<br>Some concerns (Socioeconomic status) | Low |
| **Breheny 2020** | Low | Low | Some concerns | Low | Low | Some concerns | Low | Low |
| **Brown 2013** | Some concerns | Low | Some concerns | Low | Some concerns | Some concerns | Low | Low |
| **Chai 2019** | Low | Low | High | Low | Some concerns | High | Some concerns | Low |
| **Damsgaard 2014** | Some concerns | Low | Some concerns | Low | Low | Some concerns | Low | Low |
| **Dewar 2013** | Low | Low | Some concerns | Low | Low | Some concerns | Low | Low |
| **Drummy 2016** | High | Low | Some concerns | Low | Some concerns | High | Low | Low |
| **Duncan 2019** | Some concerns | Low | Some concerns | Low | Some concerns | Some concerns | Low | Low |
| **Ebbeling 2006** | Low | Low | Low | Low | Some concerns | Some concerns | Low | Low |
| **El Ansari 2010** | Some concerns | Low | Some concerns | Low | Some concerns | Some concerns | Low | Low |
| **Fairclough 2013** | Some concerns | Low | High | Low | Some concerns | High | Low | Low |
| **Farmer 2017** | Some concerns | Low | Some concerns | Low | Some concerns | Some concerns | Low (Race/Ethnicity/Culture/Language; Gender/Sex)<br>High (Socioeconomic status) | Low |
| **Fulkerson 2015** | Some concerns | Low | Some concerns | Low | Some concerns | Some concerns | Low | Low |
| **Fulkerson 2022** | Some concerns | Low | High | Low | Some concerns | High | Low | Low |
| **Gentile 2009** | Some concerns | High | High | Low | Some concerns | High | Low (Race/Ethnicity/Culture/Language; Gender/Sex)<br>Some concerns (Socioeconomic status) | Low |

| | | | | | | | | |
|---|---|---|---|---|---|---|---|---|
| **Griffin 2019** | Low | Low | High | Low | Some concerns | High | Some concerns | Low |
| **Grydeland 2014** | Some concerns | High | Some concerns | Low | Some concerns | High | Low | Low |
| **Ha 2021** | Low | Low | Some concerns | Low | Some concerns | Some concerns | Low | Low |
| **Habib-Mourad 2014** | Some concerns | Low | Low | Low | Some concerns | Some concerns | Low | Low |
| **Habib-Mourad 2020** | High | Low | High | Low | Some concerns | High | Low | Low |
| **Haerens 2006** | Some concerns | Low | Some concerns | Low | Some concerns | Some concerns | Low | Low |
| **Hollis 2016** | Low | Low | Low | Low | Low | Low | Low | Low |
| **Hopper 2005** | High | Low | Some concerns | Low | Some concerns | High | Low | Low |
| **Ickovics 2019** | Some concerns | Low | High | Low | Some concerns | High | Low | Low |
| **Jones 2015** | Low | Low | Some concerns | Low | Some concerns | Some concerns | Low | Low |
| **Kain 2014** | High | Low | High | High | Some concerns | High | Low | Low |
| **Kennedy 2018** | Low | Low | Low | Low | Low | Low | Low | Low |
| **Kobel 2017** | Some concerns | Low | High | Low | Some concerns | High | Low | Low |
| **Kriemler 2010** | Low | Low | Low | Low | Low | Low | Low (Gender/Sex; Place of Residence) High (Education) | Low |
| **Kuroko 2020** | Some concerns | Low | Some concerns | Low | Some concerns | Some concerns | Low | Low |
| **Lana 2014**[b] | Some concerns | Low | n/a | High | n/a | High | Low | Low |
| **Levy 2012** | Some concerns | Some concerns | Low | Low | Some concerns | Some concerns | Low | Low |
| **Li 2010** | Some concerns | Low | Some concerns | Low | Some concerns | Some concerns | Low | Low |
| **Li 2019** | Low | Low | Low | Low | Low | Low | Low | Low |
| **Liu 2019** | Some concerns | Low | Low | Low | Some concerns | Some concerns | Low | Low |
| **Liu 2022** | Low | Low | Low | Low | Low | Low | Low | Low |
| **Lloyd 2018** | Low | Low | Low | Low | Low | Low | Low | Low |
| **Lubans 2021** | Some concerns | Low | High | Low | Low | High | Some concerns | Low |
| **Lynch 2016**[c] | Some concerns | Low | High | Low | n/a | High | Some concerns | Low |
| **Madsen 2013**[d] | Some concerns | Low | High | Low | n/a | High | Low | Low |
| **Marcus 2009** | High | Low | Some concerns | Low | Some concerns | High | Low | Low |
| **Martinez-Vizcaino 2014** | Some concerns | Low | Some concerns | Low | Low | Some concerns | Low | Low |
| **Martinez-Vizcaino 2020** | High | Low | Low | Low | Some concerns | High | Low | Low |
| **Morgan 2011** | Low | Low | High | Low | Low | High | Low | Low |
| **Morgan 2014** | Low | Low | Some concerns | Low | Low | Some concerns | Some concerns | Low |
| **Morgan 2019** | Low | Low | Some concerns | Low | Some concerns | Some concerns | Low | Low |
| **Muller 2019** | Some concerns | Low | Some concerns | Low | Low | Some concerns | Low | Low |
| **Nemet 2011b** | Some concerns | Some concerns | Some concerns | Low | Some concerns | Some concerns | Low | Low |
| **Newton 2014** | Low | Low | Low | Low | Some concerns | Some concerns | Low | Low |
| **Nicholl 2021** | Low | Low | Some concerns | Low | Some concerns | Some concerns | Low | Low |
| **O'Connor 2020** | Some concerns | Low | Some concerns | Low | Low | Some concerns | Low | Low |
| **Pate 2005** | Some concerns | Low | Low | Low | Some concerns | Some concerns | Low | Low |
| **Pena 2021** | High | Low | Some concerns | Low | Some concerns | High | Low | Low |

| | | | | | | | | |
|---|---|---|---|---|---|---|---|---|
| **Pfeiffer 2019** | Low | Low | High | Low | Low | High | Low | Low |
| **Puder 2011** | Low | Low | Low | Low | Low | Low | Low | Low |
| **Rosario 2012** | Low | Low | Some concerns | Low | Some concerns | Some concerns | Low | Low |
| **Rosenkranz 2010** | Some concerns | Low | Low | Low | Some concerns | Some concerns | Low | Low |
| **Rush 2012** | High | Some concerns | High | Low | Low | High | Low | Low |
| **Sacchetti 2013** | Some concerns | Some concerns | Some concerns | Low | Some concerns | Some concerns | Low | Low |
| **Salmon 2022** | Low | Low | Some concerns | Low | Low | Some concerns | Low | Low |
| **Seguin-Fowler 2021** | Low | Low | Some concerns | Low | Low | Some concerns | Low (Education) Some concerns (Race/Ethnicity/Culture/Language; Gender/Sex) | Low |
| **Sekhavat 2014** | Some concerns | Low | Some concerns | Low | Some concerns | Some concerns | Low | Low |
| **Sherwood 2019** | Low | Low | Some concerns | Low | Low | Some concerns | Low | Low |
| **Shomaker 2019** | Some concerns | Low | Low | Low | Some concerns | Some concerns | Low | Low |
| **Singh 2009** | Some concerns | Low | Some concerns | Low | Some concerns | Some concerns | Low | Low |
| **Smith 2014** | Low | Low | Some concerns | Low | Low | Some concerns | Low | Low |
| **Stettler 2015** | Some concerns | Low | Some concerns | Low | Some concerns | Some concerns | Low | Low |
| **Takacs 2020** | Some concerns | Low | Some concerns | Low | Some concerns | Some concerns | Low | Low |
| **Tanskey 2017** | High | Low | Low | Low | High | High | Low | Low |
| **TenHoor 2018[b]** | Some concerns | High | n/a | Low | n/a | High | Low | Low |
| **Vizcaino 2008** | Some concerns | Low | Some concerns | Low | Some concerns | Some concerns | Low | Low |
| **Weeks 2012** | Some concerns | Low | Low | Low | Some concerns | Some concerns | Low | Low |
| **Wendel 2016** | High | Some concerns | Some concerns | Low | Some concerns | High | Low | Low |
| **Wilksch 2015** | Some concerns | Low | Some concerns | Low | Some concerns | Some concerns | Low | Low |
| **Williamson 2012** | Some concerns | Low | Some concerns | Low | Low | Some concerns | Low | Low |
| **Xu 2017** | Some concerns | Low | Low | Low | Some concerns | Some concerns | Low | Low |
| **Yin 2012** | Some concerns | Low | High | Low | Some concerns | High | Low | Low |
| **Zhou 2019[b]** | Some concerns | Low | n/a | Low | n/a | Some concerns | NR | Low |

[a]Study not included in the Cochrane review meta-analysis due to BMI measurement at follow-up was planned, but results are not reported and we have no evidence that it was measured.
[b]Study not included in the Cochrane review meta-analysis due to BMI/zBMI being measured at follow-up but results not being reported.
[c]Study not included in the Cochrane review meta-analysis due to data not being usable (results were reported as median)
[d]Study not included in the Cochrane review meta-analysis due to data not being usable (results were reported only narratively)

Abbreviations: n/a = not applicable (the main result was not included in the Cochrane reviews meta-analyses); NR = not reported (sample size of the main results was not reported).

Supplementary Table 4: Results of subset analyses according to study-level subgroups for the younger age group

| Analysis | Inequity subgroup 1 | Inequity subgroup 2 | Study subgroup | N studies | N ptts subgroup 1 | N ptts subgroup 2 | Difference in inequity subgroups | 95% CI | $I^2$ | $P_{het}$ (for heterogeneity) | P for interaction* |
|---|---|---|---|---|---|---|---|---|---|---|---|
| **zBMI** | | | | | | | | | | | |
| Gender/sex by intervention type | Male | Female | Diet | 6 | 763 | 743 | 0.00 | -0.06, 0.07 | 0% | 0.9 | 0.4 |
| | | | Activity | 14 | 6196 | 6020 | 0.01 | -0.04, 0.06 | 0% | 0.5 | |
| | | | Diet and activity | 28 | 15938 | 15308 | 0.04 | 0.01, 0.07 | 0% | 0.6 | |
| Gender/sex by country income status | Male | Female | Higher income status | 36 | 13178 | 12795 | 0.03 | 0.00, 0.06 | 1% | 0.4 | 0.8 |
| | | | Lower income status | 9 | 9603 | 9164 | 0.04 | -0.02, 0.10 | 0% | 0.9 | |
| SES by country income status | Higher SES | Lower SES | Higher income status | 24 | 7450 | 6625 | 0.01 | -0.03, 0.06 | 41% | 0.02 | 0.9 |
| | | | Lower income status | 5 | 2330 | 1878 | 0.01 | -0.11, 0.12 | 60% | 0.04 | |
| Education by country income status | Higher education | Lower education | Higher income status | 11 | 1983 | 2703 | 0.01 | -0.07, 0.08 | 34% | 0.1 | 0.3 |
| | | | Lower income status | 3 | 1768 | 1158 | 0.07 | -0.03, 0.17 | 0% | 0.6 | |
| **BMI** | | | | | | | | | | | |
| Gender/sex by intervention type | Male | Female | Diet | 12 | 5260 | 5072 | 0.05 | -0.09, 0.19 | 0% | 0.8 | 0.5 |
| | | | Activity | 3 | 433 | 406 | -0.02 | -0.19, 0.14 | 0% | 0.9 | |
| | | | Diet and activity | 16 | 8053 | 7859 | 0.10 | -0.02, 0.22 | 11% | 0.3 | |
| Gender/sex by country income status | Male | Female | Higher income status | 24 | 5700 | 5753 | 0.07 | -0.03, 0.16 | 0% | 0.6 | 0.9 |
| | | | Lower income status | 7 | 8046 | 7584 | 0.05 | -0.11, 0.21 | 0% | 0.6 | |
| SES by country income status | Higher SES | Lower SES | Higher income status | 14 | 3678 | 3537 | -0.02 | -0.20, 0.16 | 61% | 0.002 | 0.2 |
| | | | Lower income status | 3 | 1219 | 1058 | 0.15 | -0.07, 0.37 | 0% | 0.5 | |
| Education by country income status | Higher education | Lower education | Higher income status | 9 | 2273 | 2484 | -0.08 | -0.26, 0.11 | 54% | 0.03 | 0.1 |
| | | | Lower income status | 2 | 822 | 579 | 0.23 | -0.03, 0.49 | 0% | 0.4 | |

Abbreviations: SES = socioeconomic status; Ptts = participants.

*Examines difference between the study subgroups, from test for subgroup differences based on random-effects meta-analysis

Supplementary Table 5: Results of subset analyses according to study-level subgroups for the older age group

| Analysis | Inequity subgroup 1 | Inequity subgroup 2 | Study subgroup | N studies | N ptts subgroup 1 | N ptts subgroup 2 | Difference in inequity subgroups | 95% CI | $I^2$ | $P_{het}$ (for heterogeneity) | P for interaction* |
|---|---|---|---|---|---|---|---|---|---|---|---|
| **zBMI** | | | | | | | | | | | |
| Gender/sex by intervention type | Male | Female | Diet | 1 | 42 | 71 | 0.14 | -0.49, 0.78 | n/a | n/a | <0.0001 |
| | | | Activity | 3 | 941 | 1006 | 0.02 | -0.10, 0.15 | 0 | 0.9 | |
| | | | Diet and Activity | 2 | 2326 | 1667 | -0.51 | -0.64, -0.38 | 0 | 0.4 | |
| **BMI** | | | | | | | | | | | |
| Gender/sex by intervention type | Male | Female | Diet | 3 | 158 | 190 | -0.32 | -1.18, 0.55 | 39 | 0.2 | 0.8 |
| | | | Activity | 6 | 1736 | 1760 | -0.05 | -0.30, 0.21 | 20 | 0.3 | |
| | | | Diet and Activity | 5 | 3578 | 3363 | -0.14 | -0.38, 0.10 | 0 | 0.4 | |
| Gender/sex by country income status | Male | Female | Higher income status | 3 | 965 | 937 | -0.36 | -1.01, 0.28 | 53 | 0.1 | 0.4 |
| | | | Lower income status | 11 | 4507 | 4376 | -0.08 | -0.27, 0.12 | 0 | 0.5 | |
| SES by country income status | Higher SES | Lower SES | Higher income status | 2 | 751 | 970 | -0.03 | -0.41, 0.36 | 0 | 0.9 | 0.8 |
| | | | Lower income status | 5 | 1067 | 1812 | -0.09 | -0.45, 0.27 | 0 | 0.9 | |
| Education by country income status | Higher education | Lower education | Higher income status | 1 | 395 | 665 | 0.15 | -0.68, 0.98 | n/a | n/a | 0.1 |
| | | | Lower income status | 1 | 491 | 236 | 2.28 | 0.27, 4.29 | n/a | n/a | |

Abbreviations: SES = socioeconomic status; Ptts = participants.
*Examines difference between the study subgroups, from test for subgroup differences based on random-effects meta-analysis

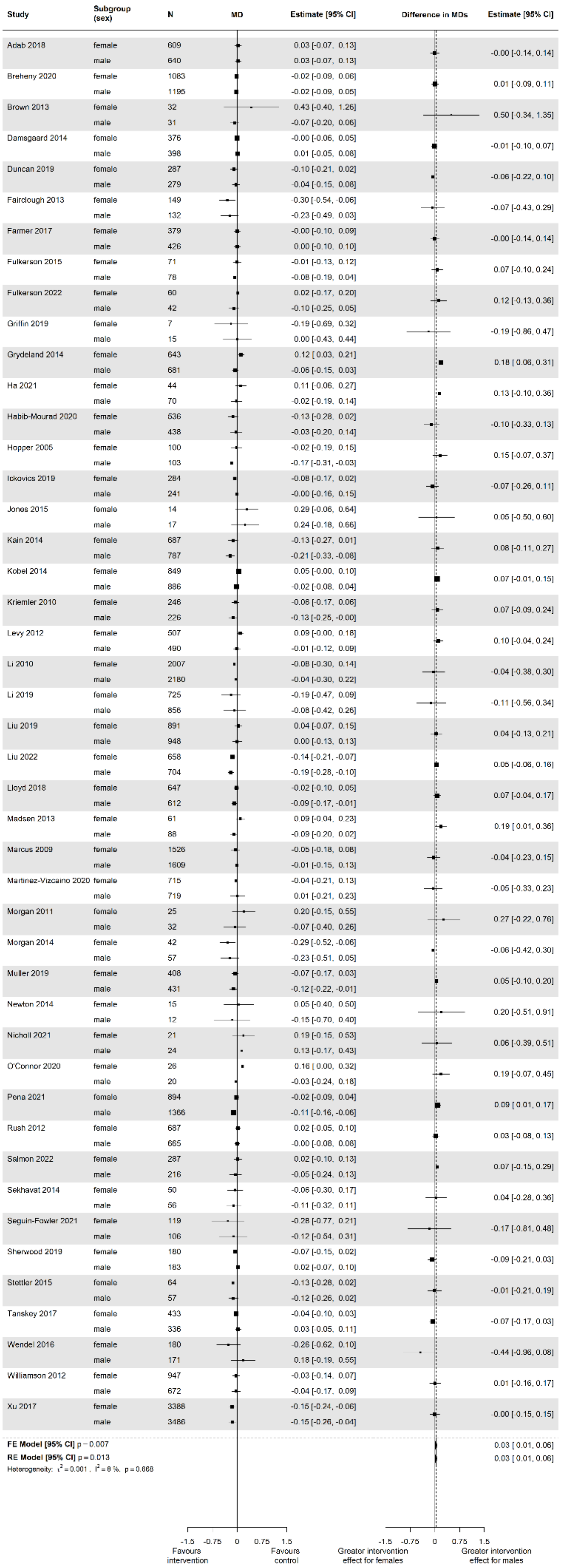

Supplementary Figure 1: Estimates of intervention effect for separate subgroups (left) and differences in intervention effect between subgroups (interactions; right) for factor **gender/sex** and outcome **zBMI** in the **younger age group** (5-11 years)

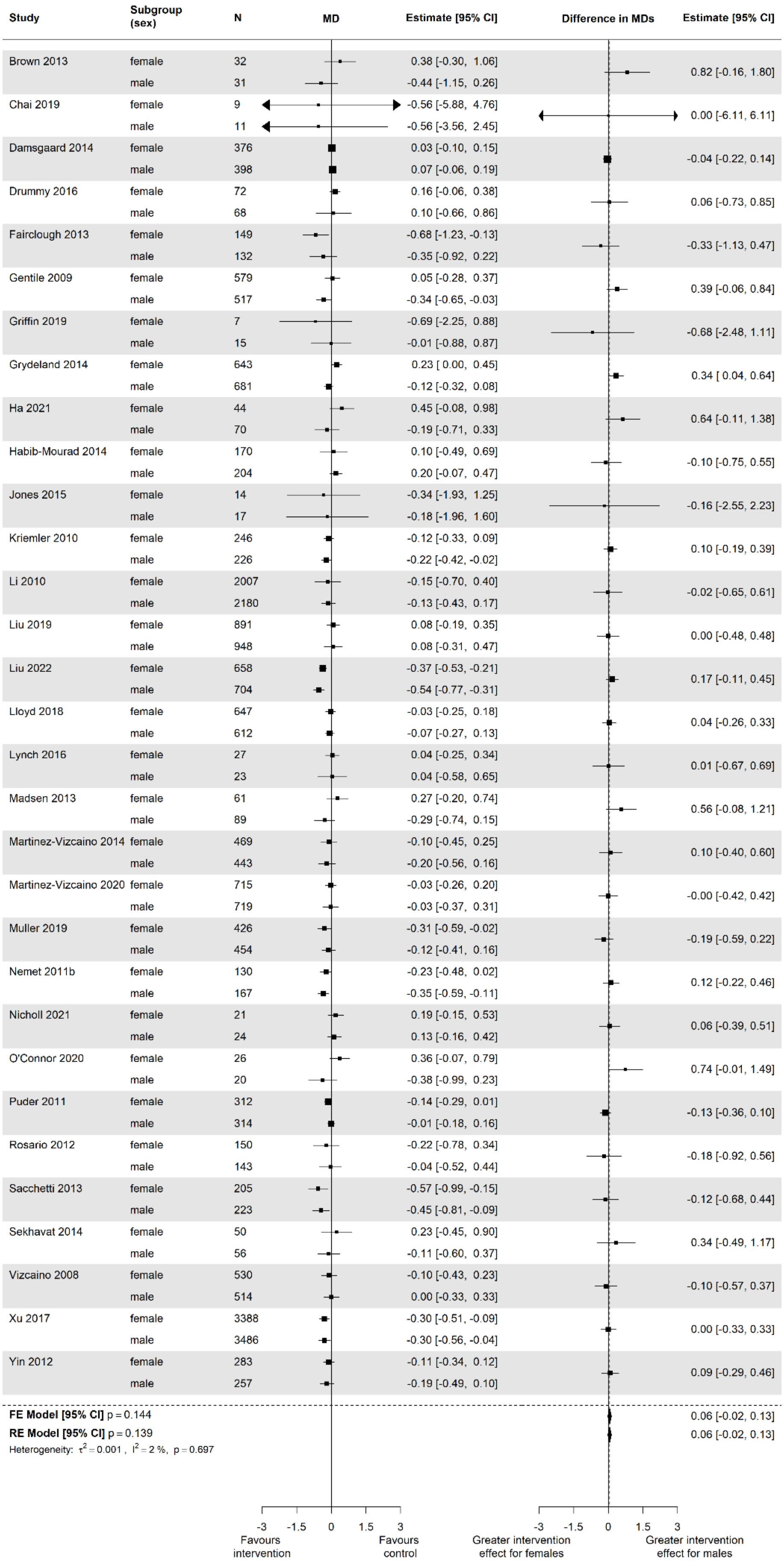

Supplementary Figure 2: Estimates of intervention effect for separate subgroups (left) and differences in intervention effect between subgroups (interactions; right) for factor **gender/sex** and outcome **BMI** in the **younger age group** (5-11 years)

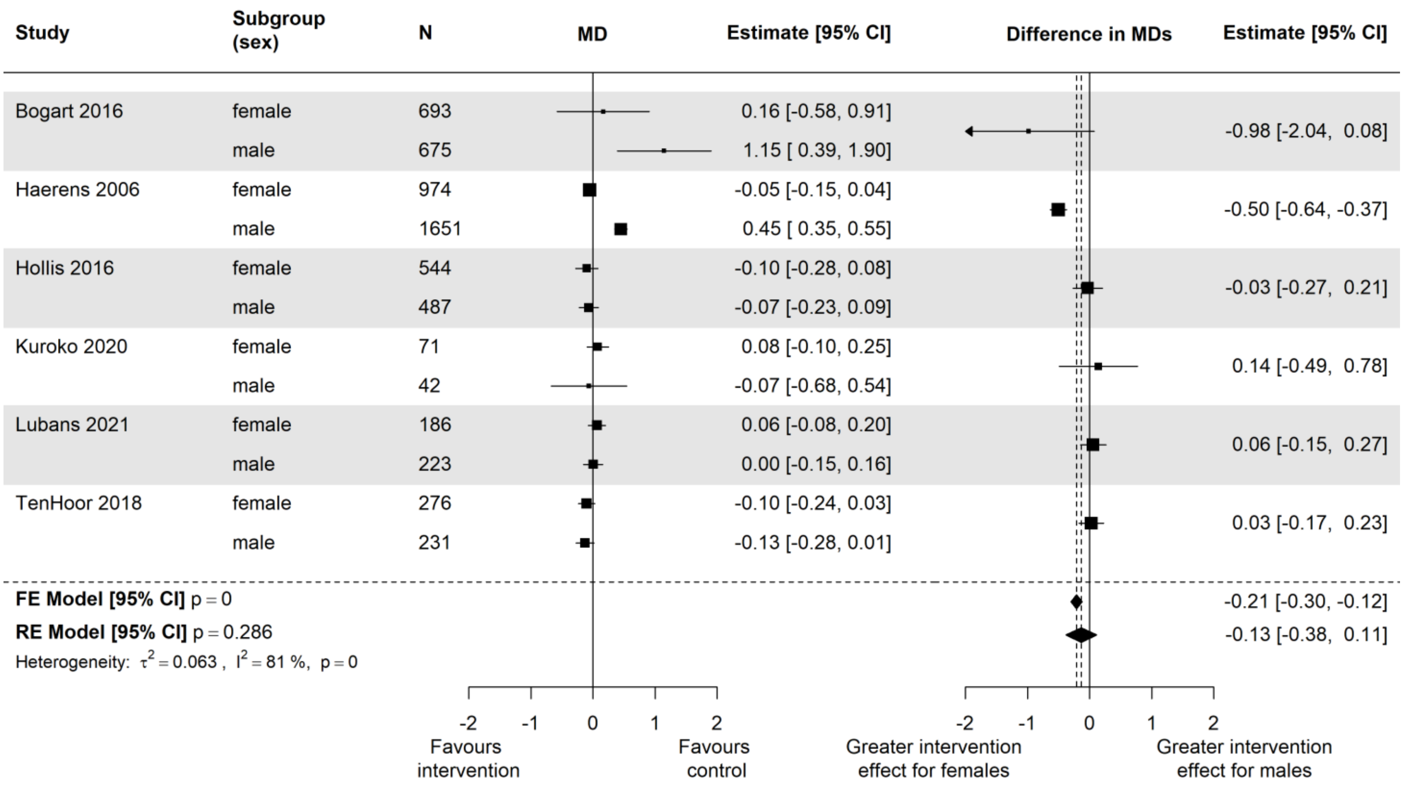

Supplementary Figure 3: Estimates of intervention effect for separate subgroups (left) and differences in intervention effect between subgroups (interactions; right) for factor **gender/sex** and outcome **zBMI** in the **older age group** (12-18 years)

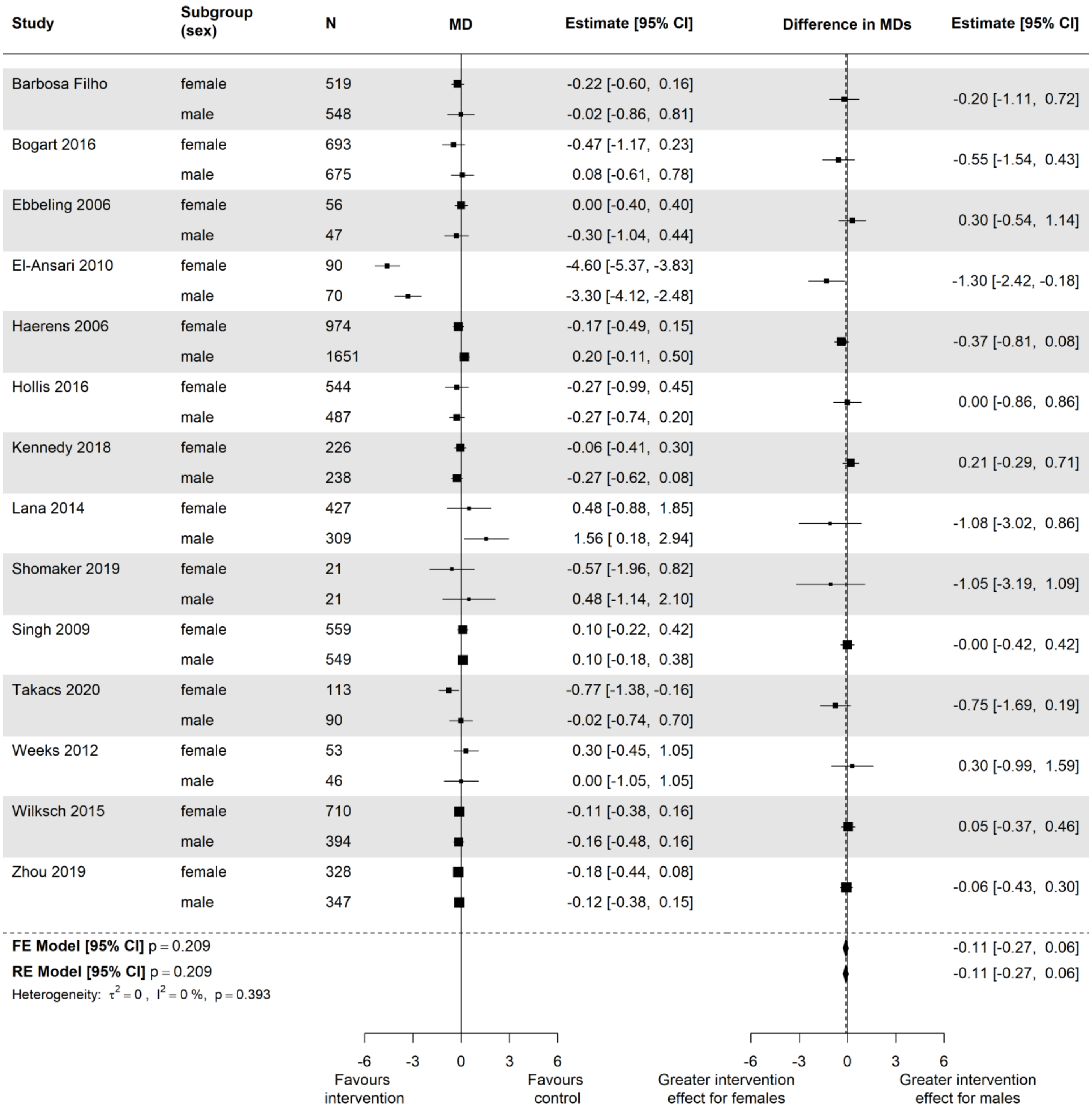

Supplementary Figure 4: Estimates of intervention effect for separate subgroups (left) and differences in intervention effect between subgroups (interactions; right) for factor **gender/sex** and outcome **BMI** in the **older age group** (12-18 years)

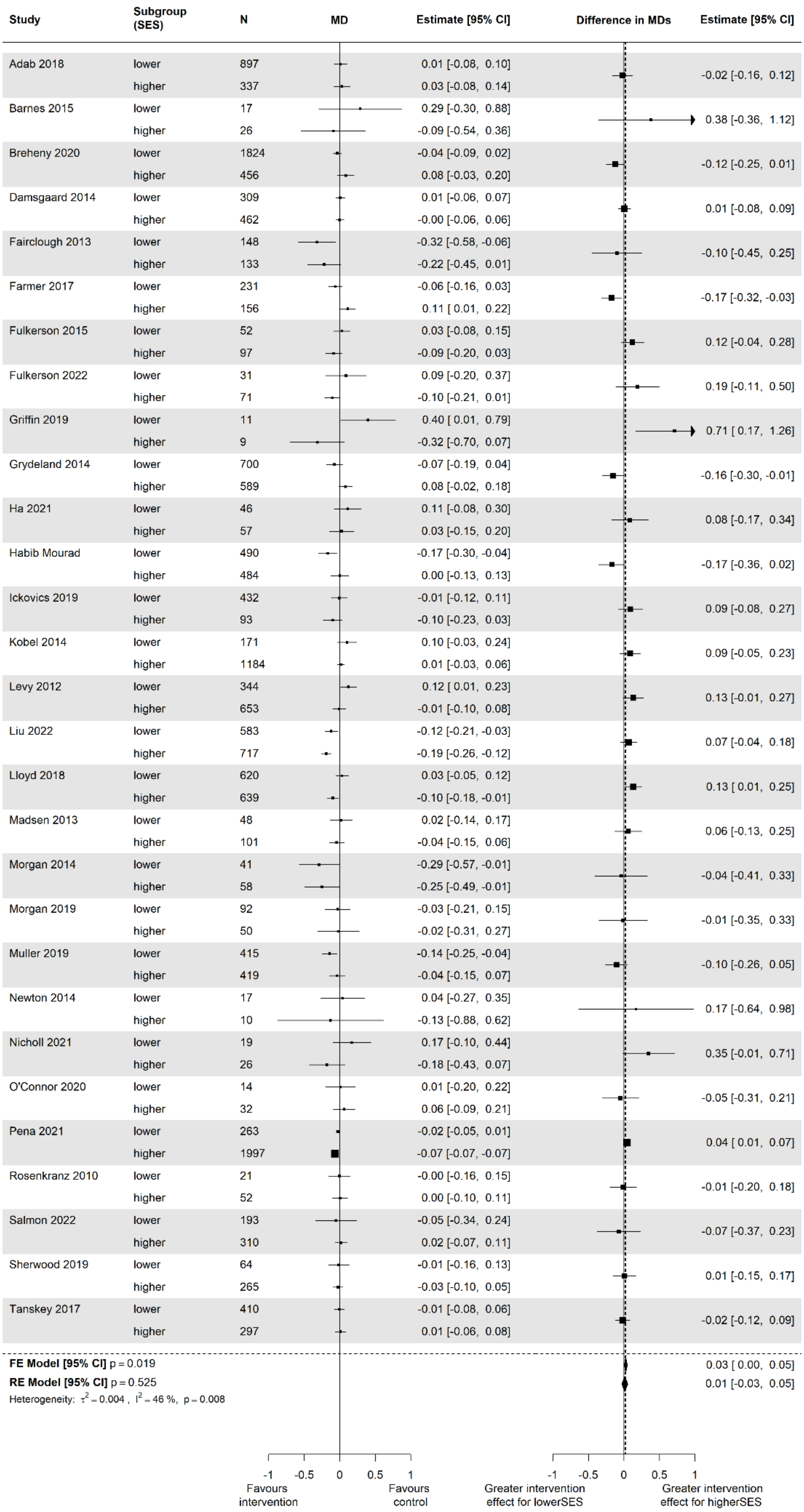
Study | Subgroup (SES) | N | MD | Estimate [95% CI] | Difference in MDs | Estimate [95% CI]
Adab 2018 | lower | 897 | 0.01 [-0.08, 0.10]
higher | 337 | 0.03 [-0.08, 0.14] | -0.02 [-0.16, 0.12]
Barnes 2015 | lower | 17 | 0.29 [-0.30, 0.88]
higher | 26 | -0.09 [-0.54, 0.36] | 0.38 [-0.36, 1.12]
Breheny 2020 | lower | 1824 | -0.04 [-0.09, 0.02]
higher | 456 | 0.08 [-0.03, 0.20] | -0.12 [-0.25, 0.01]
Damsgaard 2014 | lower | 309 | 0.01 [-0.06, 0.07]
higher | 462 | -0.00 [-0.06, 0.06] | 0.01 [-0.08, 0.09]
Fairclough 2013 | lower | 148 | -0.32 [-0.58, -0.06]
higher | 133 | -0.22 [-0.45, 0.01] | -0.10 [-0.45, 0.25]
Farmer 2017 | lower | 231 | -0.06 [-0.16, 0.03]
higher | 156 | 0.11 [ 0.01, 0.22] | -0.17 [-0.32, -0.03]
Fulkerson 2015 | lower | 52 | 0.03 [-0.08, 0.15]
higher | 97 | -0.09 [-0.20, 0.03] | 0.12 [-0.04, 0.28]
Fulkerson 2022 | lower | 31 | 0.09 [-0.20, 0.37]
higher | 71 | -0.10 [-0.21, 0.01] | 0.19 [-0.11, 0.50]
Griffin 2019 | lower | 11 | 0.40 [ 0.01, 0.79]
higher | 9 | -0.32 [-0.70, 0.07] | 0.71 [ 0.17, 1.26]
Grydeland 2014 | lower | 700 | -0.07 [-0.19, 0.04]
higher | 589 | 0.08 [-0.02, 0.18] | -0.16 [-0.30, -0.01]
Ha 2021 | lower | 46 | 0.11 [-0.08, 0.30]
higher | 57 | 0.03 [-0.15, 0.20] | 0.08 [-0.17, 0.34]
Habib Mourad | lower | 490 | -0.17 [-0.30, -0.04]
higher | 484 | 0.00 [-0.13, 0.13] | -0.17 [-0.36, 0.02]
Ickovics 2019 | lower | 432 | -0.01 [-0.12, 0.11]
higher | 93 | -0.10 [-0.23, 0.03] | 0.09 [-0.08, 0.27]
Kobel 2014 | lower | 171 | 0.10 [-0.03, 0.24]
higher | 1184 | 0.01 [-0.03, 0.06] | 0.09 [-0.05, 0.23]
Levy 2012 | lower | 344 | 0.12 [ 0.01, 0.23]
higher | 653 | -0.01 [-0.10, 0.08] | 0.13 [-0.01, 0.27]
Liu 2022 | lower | 583 | -0.12 [-0.21, -0.03]
higher | 717 | -0.19 [-0.26, -0.12] | 0.07 [-0.04, 0.18]
Lloyd 2018 | lower | 620 | 0.03 [-0.05, 0.12]
higher | 639 | -0.10 [-0.18, -0.01] | 0.13 [ 0.01, 0.25]
Madsen 2013 | lower | 48 | 0.02 [-0.14, 0.17]
higher | 101 | -0.04 [-0.15, 0.06] | 0.06 [-0.13, 0.25]
Morgan 2014 | lower | 41 | -0.29 [-0.57, -0.01]
higher | 58 | -0.25 [-0.49, -0.01] | -0.04 [-0.41, 0.33]
Morgan 2019 | lower | 92 | -0.03 [-0.21, 0.15]
higher | 50 | -0.02 [-0.31, 0.27] | -0.01 [-0.35, 0.33]
Muller 2019 | lower | 415 | -0.14 [-0.25, -0.04]
higher | 419 | -0.04 [-0.15, 0.07] | -0.10 [-0.26, 0.05]
Newton 2014 | lower | 17 | 0.04 [-0.27, 0.35]
higher | 10 | -0.13 [-0.88, 0.62] | 0.17 [-0.64, 0.98]
Nicholl 2021 | lower | 19 | 0.17 [-0.10, 0.44]
higher | 26 | -0.18 [-0.43, 0.07] | 0.35 [-0.01, 0.71]
O'Connor 2020 | lower | 14 | 0.01 [-0.20, 0.22]
higher | 32 | 0.06 [-0.09, 0.21] | -0.05 [-0.31, 0.21]
Pena 2021 | lower | 263 | -0.02 [-0.05, 0.01]
higher | 1997 | -0.07 [-0.07, -0.07] | 0.04 [ 0.01, 0.07]
Rosenkranz 2010 | lower | 21 | -0.00 [-0.16, 0.15]
higher | 52 | 0.00 [-0.10, 0.11] | -0.01 [-0.20, 0.18]
Salmon 2022 | lower | 193 | -0.05 [-0.34, 0.24]
higher | 310 | 0.02 [-0.07, 0.11] | -0.07 [-0.37, 0.23]
Sherwood 2019 | lower | 64 | -0.01 [-0.16, 0.13]
higher | 265 | -0.03 [-0.10, 0.05] | 0.01 [-0.15, 0.17]
Tanskey 2017 | lower | 410 | -0.01 [-0.08, 0.06]
higher | 297 | 0.01 [-0.06, 0.08] | -0.02 [-0.12, 0.09]
FE Model [95% CI] p = 0.019 | 0.03 [ 0.00, 0.05]
RE Model [95% CI] p = 0.525 | 0.01 [-0.03, 0.05]
Heterogeneity: τ² = 0.004 , I² = 46 %, p = 0.008
-1 -0.5 0 0.5 1
Favours intervention
Favours control
-1 -0.5 0 0.5 1
Greater intervention effect for lowerSES
Greater intervention effect for higherSES

Supplementary Figure 5: Estimates of intervention effect for separate subgroups (left) and differences in intervention effect between subgroups (interactions; right) for factor **socioeconomic status** and outcome **zBMI** in the **younger age group** (5-11 years)

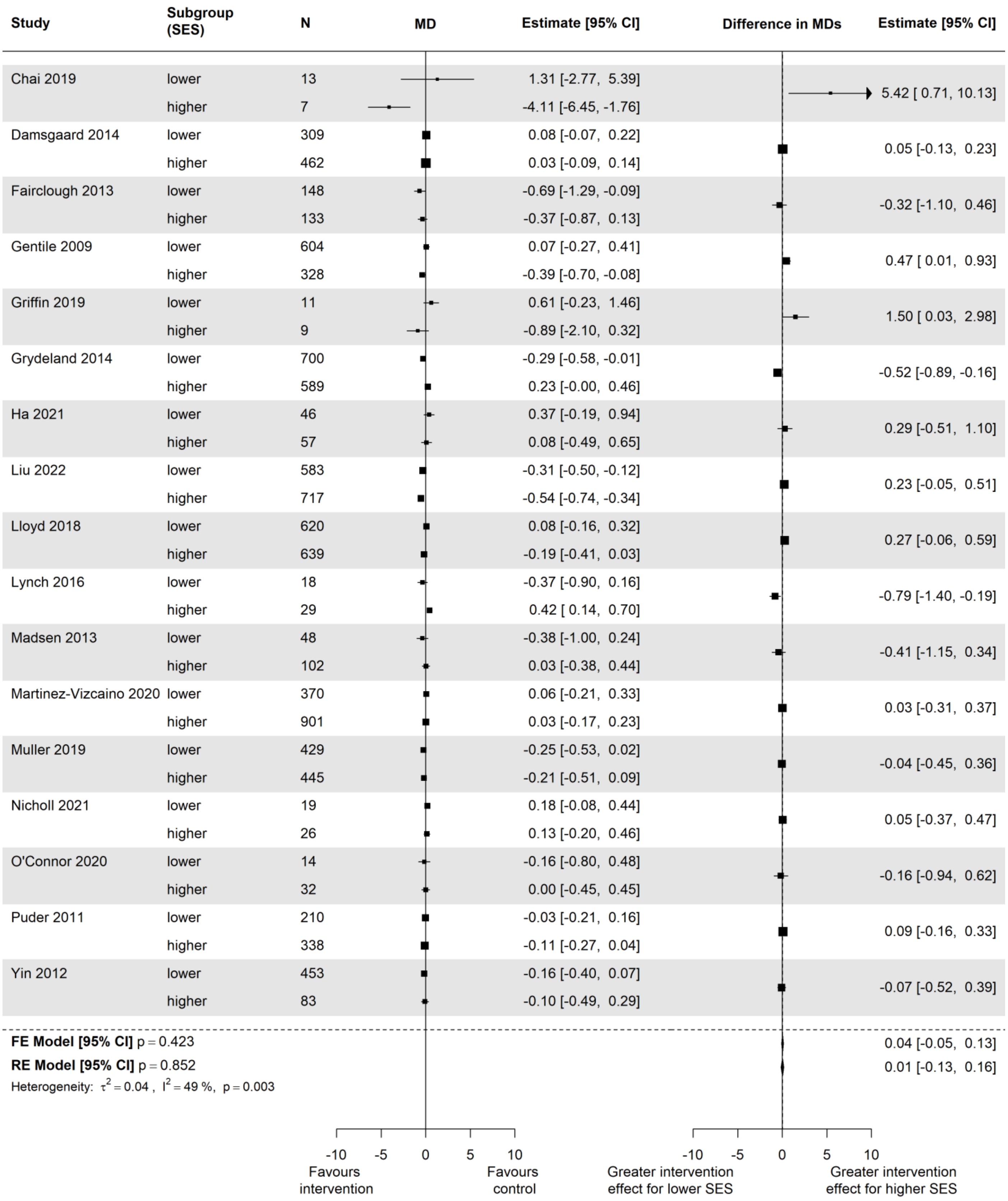

Supplementary Figure 6: Estimates of intervention effect for separate subgroups (left) and differences in intervention effect between subgroups (interactions; right) for factor **socioeconomic status** and outcome **BMI** in the **younger age group** (5-11 years)

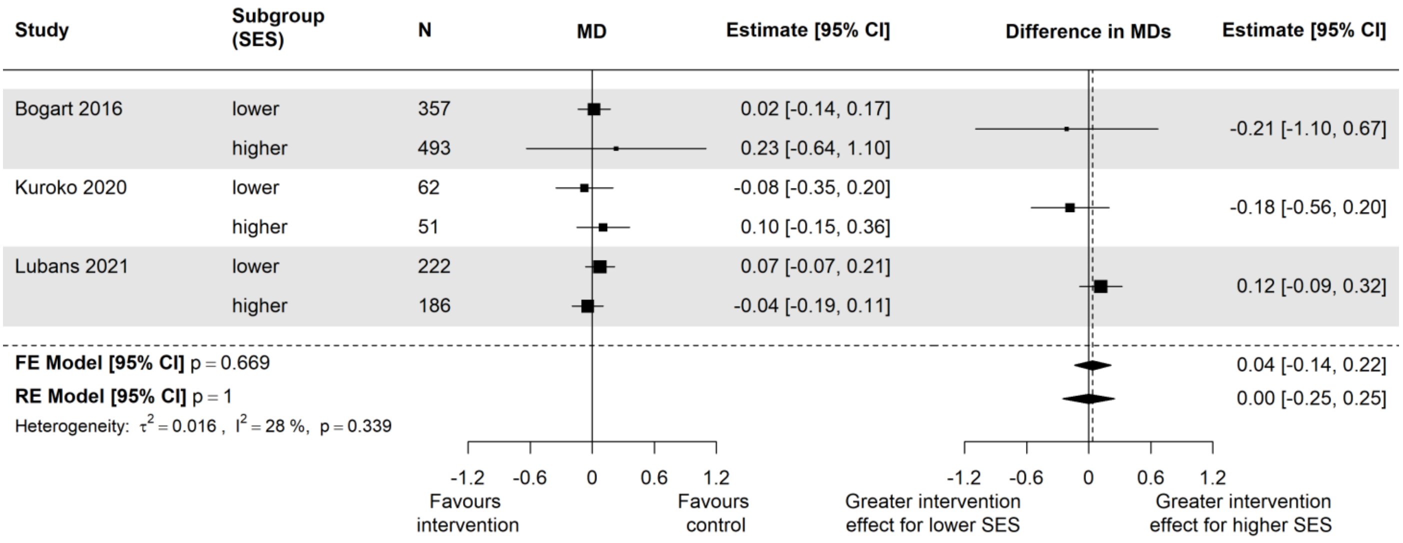

Supplementary Figure 7: Estimates of intervention effect for separate subgroups (left) and differences in intervention effect between subgroups (interactions; right) for factor **socioeconomic status** and outcome **zBMI** in the **older age group** (12-18 years)

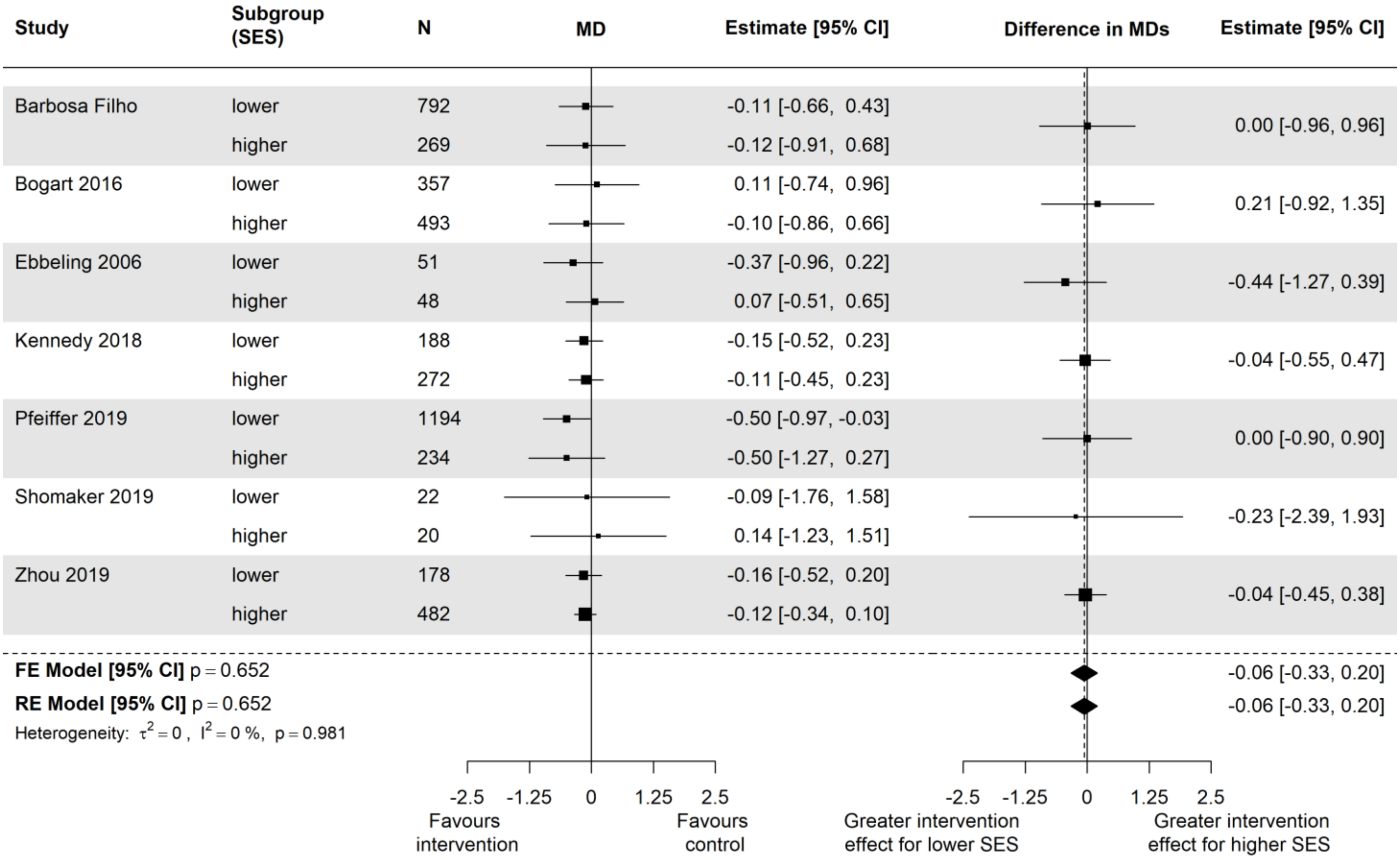

Supplementary Figure 8: Estimates of intervention effect for separate subgroups (left) and differences in intervention effect between subgroups (interactions; right) for factor **socioeconomic status** and outcome **BMI** in the **older age group** (12-18 years)

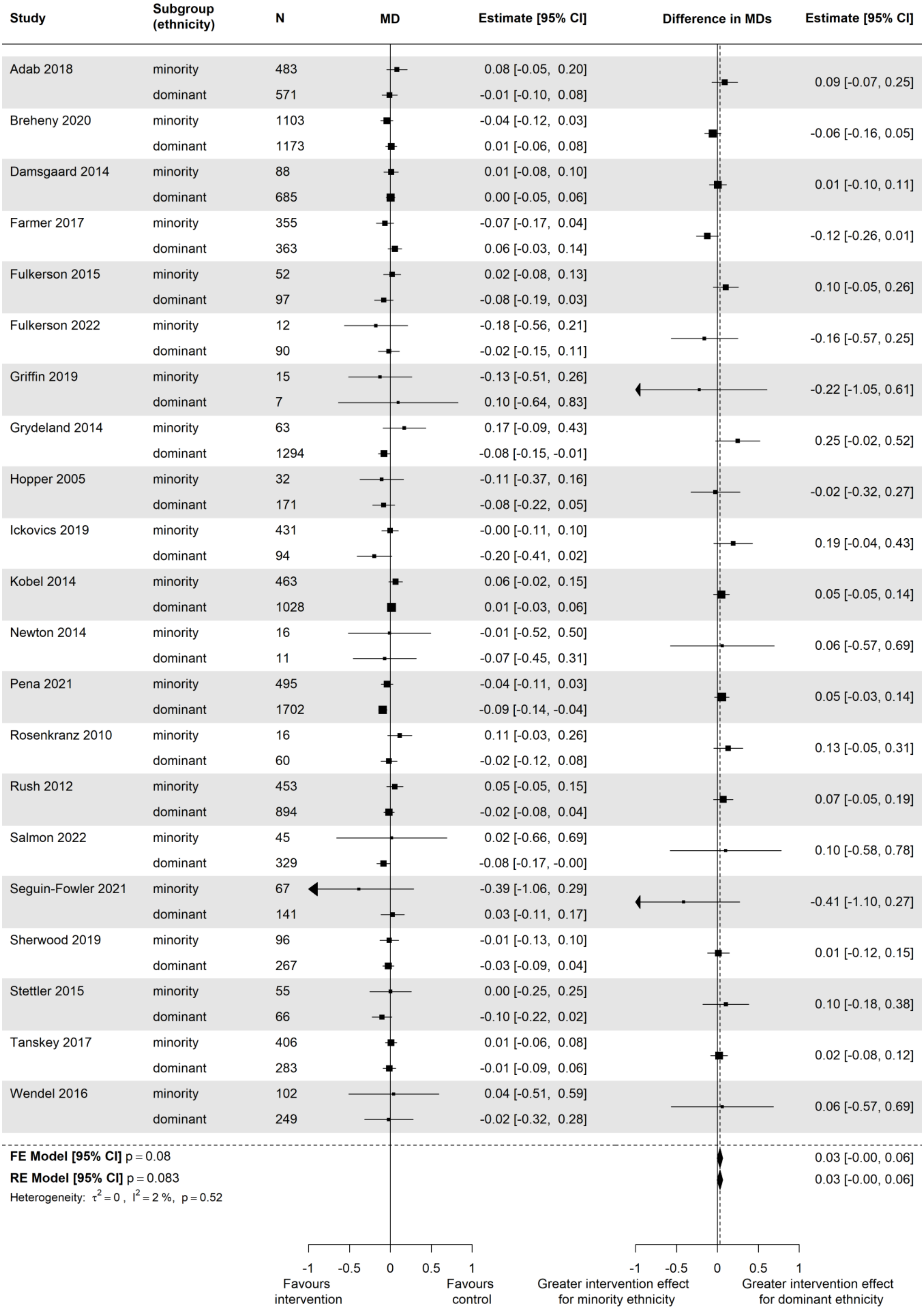
Study
Subgroup (ethnicity)
N
MD
Estimate [95% CI]
Difference in MDs
Estimate [95% CI]
Adab 2018 minority 483 0.08 [-0.05, 0.20]
dominant 571 -0.01 [-0.10, 0.08]
0.09 [-0.07, 0.25]
Breheny 2020 minority 1103 -0.04 [-0.12, 0.03]
dominant 1173 0.01 [-0.06, 0.08]
-0.06 [-0.16, 0.05]
Damsgaard 2014 minority 88 0.01 [-0.08, 0.10]
dominant 685 0.00 [-0.05, 0.06]
0.01 [-0.10, 0.11]
Farmer 2017 minority 355 -0.07 [-0.17, 0.04]
dominant 363 0.06 [-0.03, 0.14]
-0.12 [-0.26, 0.01]
Fulkerson 2015 minority 52 0.02 [-0.08, 0.13]
dominant 97 -0.08 [-0.19, 0.03]
0.10 [-0.05, 0.26]
Fulkerson 2022 minority 12 -0.18 [-0.56, 0.21]
dominant 90 -0.02 [-0.15, 0.11]
-0.16 [-0.57, 0.25]
Griffin 2019 minority 15 -0.13 [-0.51, 0.26]
dominant 7 0.10 [-0.64, 0.83]
-0.22 [-1.05, 0.61]
Grydeland 2014 minority 63 0.17 [-0.09, 0.43]
dominant 1294 -0.08 [-0.15, -0.01]
0.25 [-0.02, 0.52]
Hopper 2005 minority 32 -0.11 [-0.37, 0.16]
dominant 171 -0.08 [-0.22, 0.05]
-0.02 [-0.32, 0.27]
Ickovics 2019 minority 431 -0.00 [-0.11, 0.10]
dominant 94 -0.20 [-0.41, 0.02]
0.19 [-0.04, 0.43]
Kobel 2014 minority 463 0.06 [-0.02, 0.15]
dominant 1028 0.01 [-0.03, 0.06]
0.05 [-0.05, 0.14]
Newton 2014 minority 16 -0.01 [-0.52, 0.50]
dominant 11 -0.07 [-0.45, 0.31]
0.06 [-0.57, 0.69]
Pena 2021 minority 495 -0.04 [-0.11, 0.03]
dominant 1702 -0.09 [-0.14, -0.04]
0.05 [-0.03, 0.14]
Rosenkranz 2010 minority 16 0.11 [-0.03, 0.26]
dominant 60 -0.02 [-0.12, 0.08]
0.13 [-0.05, 0.31]
Rush 2012 minority 453 0.05 [-0.05, 0.15]
dominant 894 -0.02 [-0.08, 0.04]
0.07 [-0.05, 0.19]
Salmon 2022 minority 45 0.02 [-0.66, 0.69]
dominant 329 -0.08 [-0.17, -0.00]
0.10 [-0.58, 0.78]
Seguin-Fowler 2021 minority 67 -0.39 [-1.06, 0.29]
dominant 141 0.03 [-0.11, 0.17]
-0.41 [-1.10, 0.27]
Sherwood 2019 minority 96 -0.01 [-0.13, 0.10]
dominant 267 -0.03 [-0.09, 0.04]
0.01 [-0.12, 0.15]
Stettler 2015 minority 55 0.00 [-0.25, 0.25]
dominant 66 -0.10 [-0.22, 0.02]
0.10 [-0.18, 0.38]
Tanskey 2017 minority 406 0.01 [-0.06, 0.08]
dominant 283 -0.01 [-0.09, 0.06]
0.02 [-0.08, 0.12]
Wendel 2016 minority 102 0.04 [-0.51, 0.59]
dominant 249 -0.02 [-0.32, 0.28]
0.06 [-0.57, 0.69]
FE Model [95% CI] p = 0.08 0.03 [-0.00, 0.06]
RE Model [95% CI] p = 0.083 0.03 [-0.00, 0.06]
Heterogeneity: τ² = 0 , I² = 2 %, p = 0.52
-1 -0.5 0 0.5 1
Favours intervention
Favours control
-1 -0.5 0 0.5 1
Greater intervention effect for minority ethnicity
Greater intervention effect for dominant ethnicity

Supplementary Figure 9: Estimates of intervention effect for separate subgroups (left) and differences in intervention effect between subgroups (interactions; right) for factor **ethnicity** and outcome **zBMI** in the **younger age group** (5-11 years)

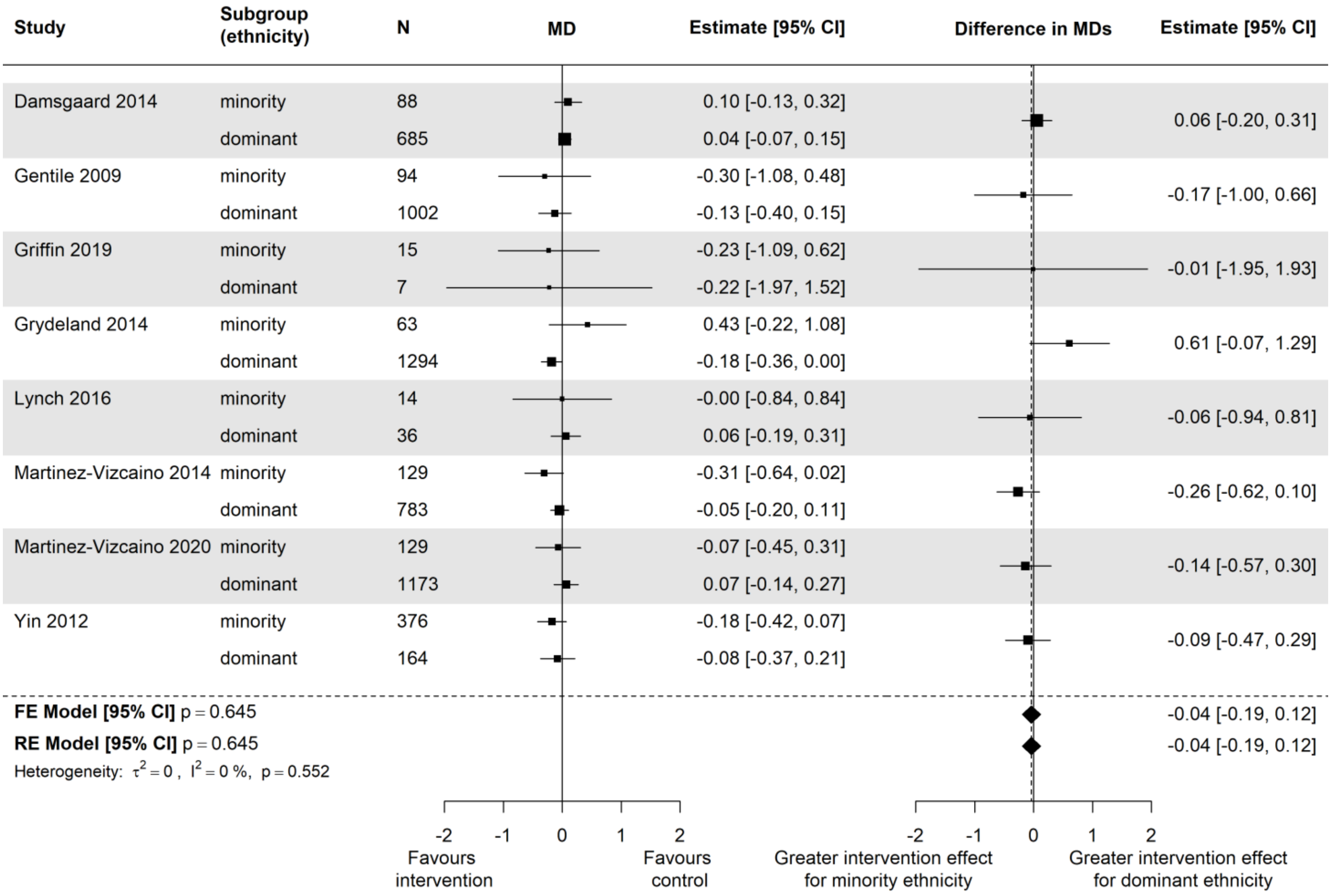

Supplementary Figure 10: Estimates of intervention effect for separate subgroups (left) and differences in intervention effect between subgroups (interactions; right) for factor **ethnicity** and outcome **BMI** in the **younger age group** (5-11 years)

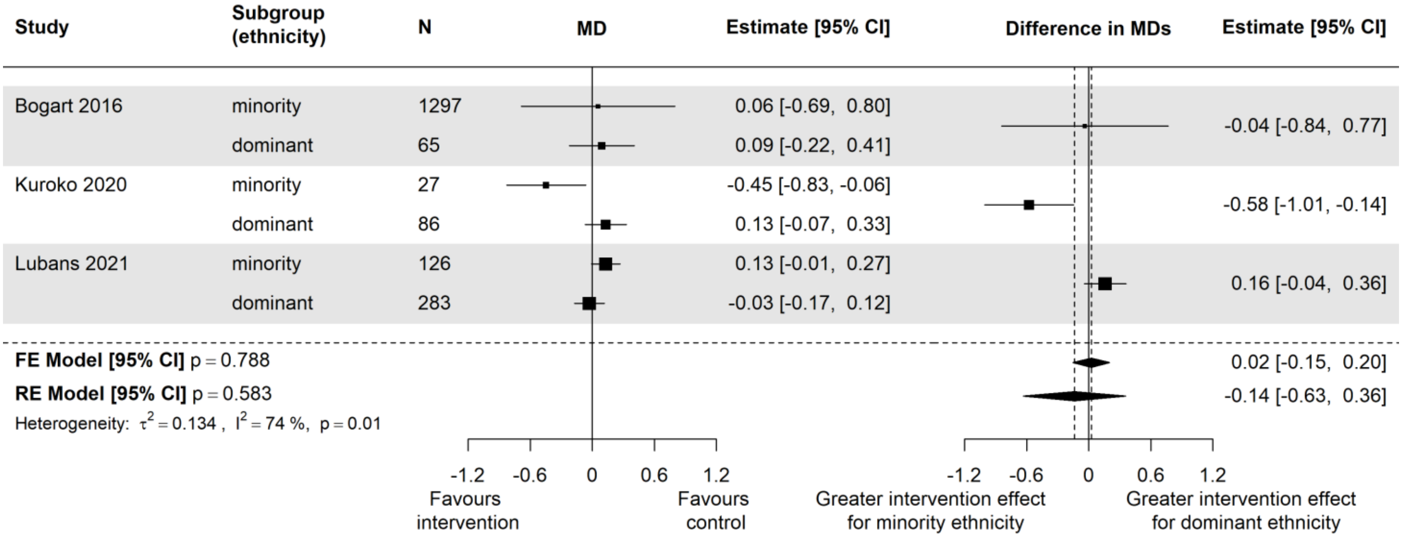

Supplementary Figure 11: Estimates of intervention effect for separate subgroups (left) and differences in intervention effect between subgroups (interactions; right) for factor **ethnicity** and outcome **zBMI** in the **older age group** (12-18 years)

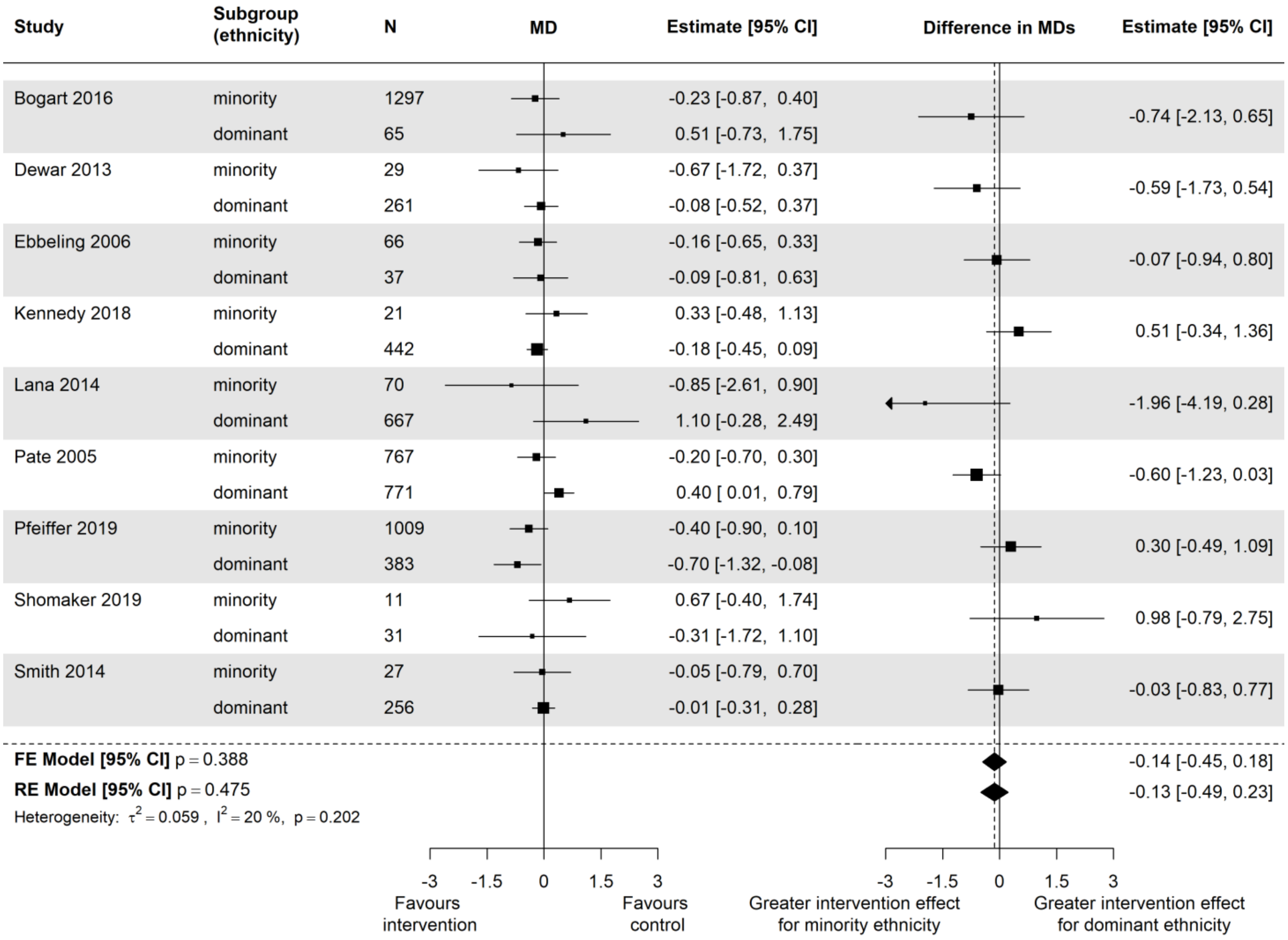

Supplementary Figure 12: Estimates of intervention effect for separate subgroups (left) and differences in intervention effect between subgroups (interactions; right) for factor **ethnicity** and outcome **BMI** in the **older age group** (12-18 years)

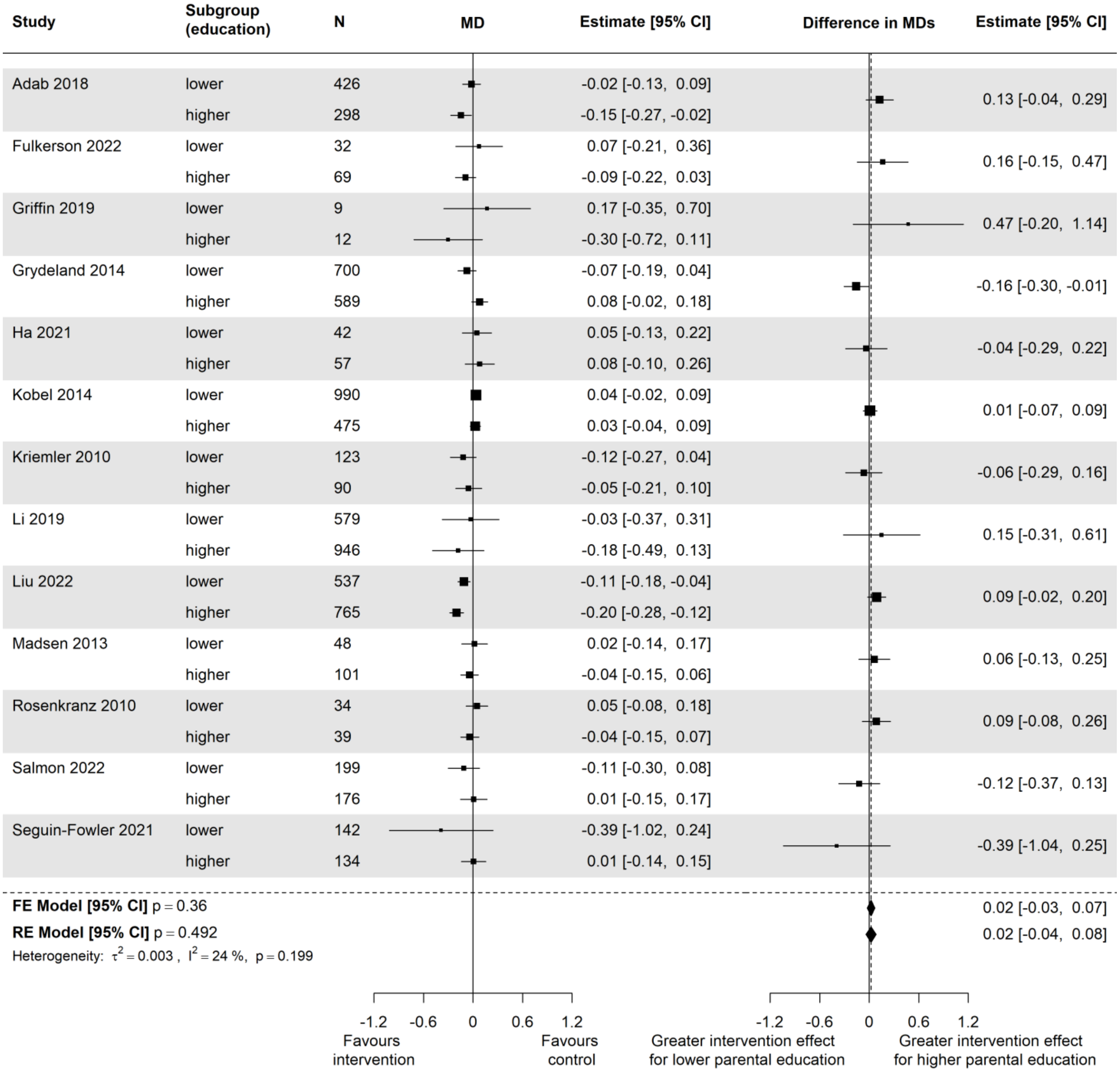

Supplementary Figure 13: Estimates of intervention effect for separate subgroups (left) and differences in intervention effect between subgroups (interactions; right) for factor **(parental) education** and outcome **zBMI** in the **younger age group** (5-11 years)

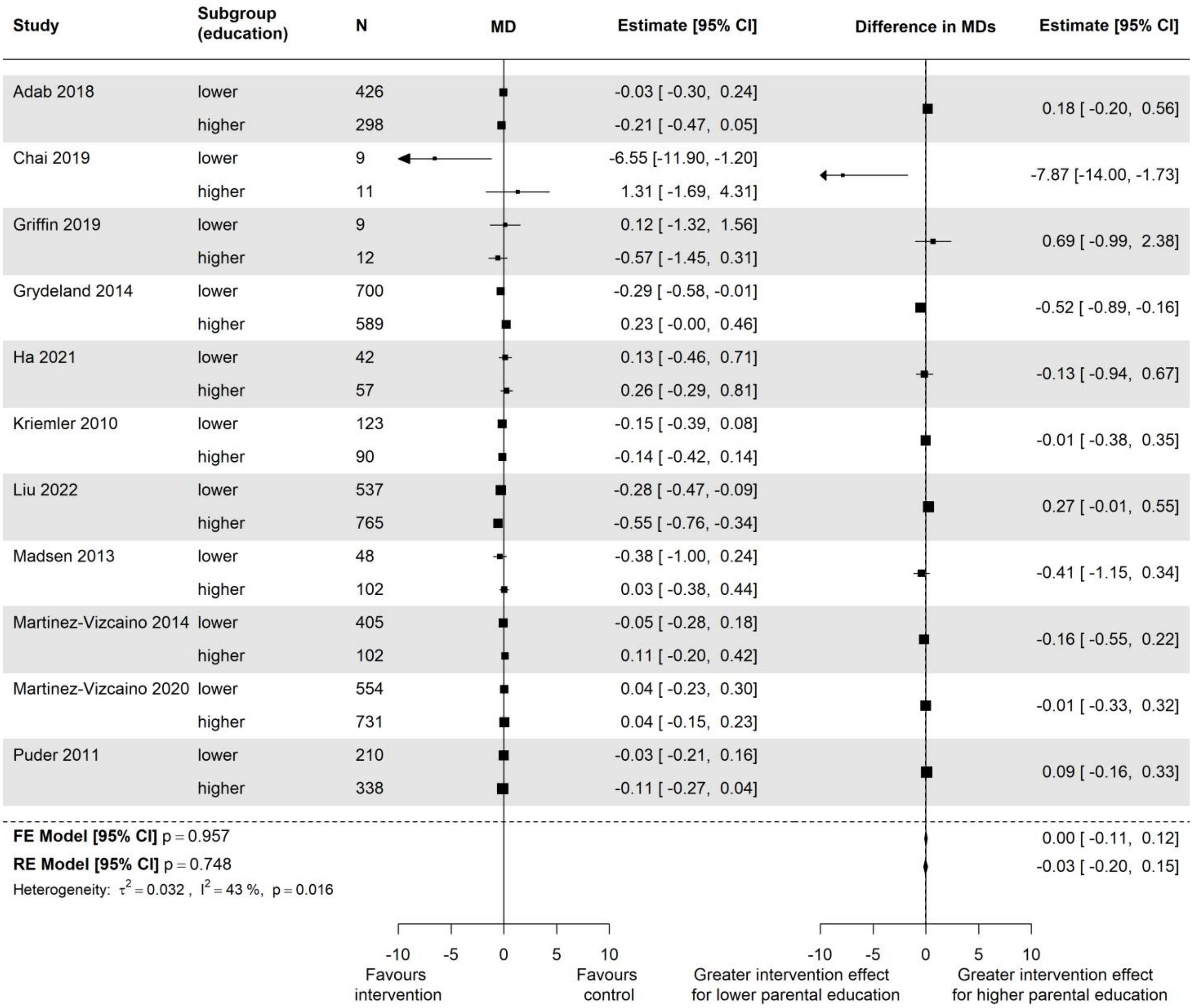

Supplementary Figure 14: Estimates of intervention effect for separate subgroups (left) and differences in intervention effect between subgroups (interactions; right) for factor **(parental) education** and outcome **BMI** in the **younger age group** (5-11 years)

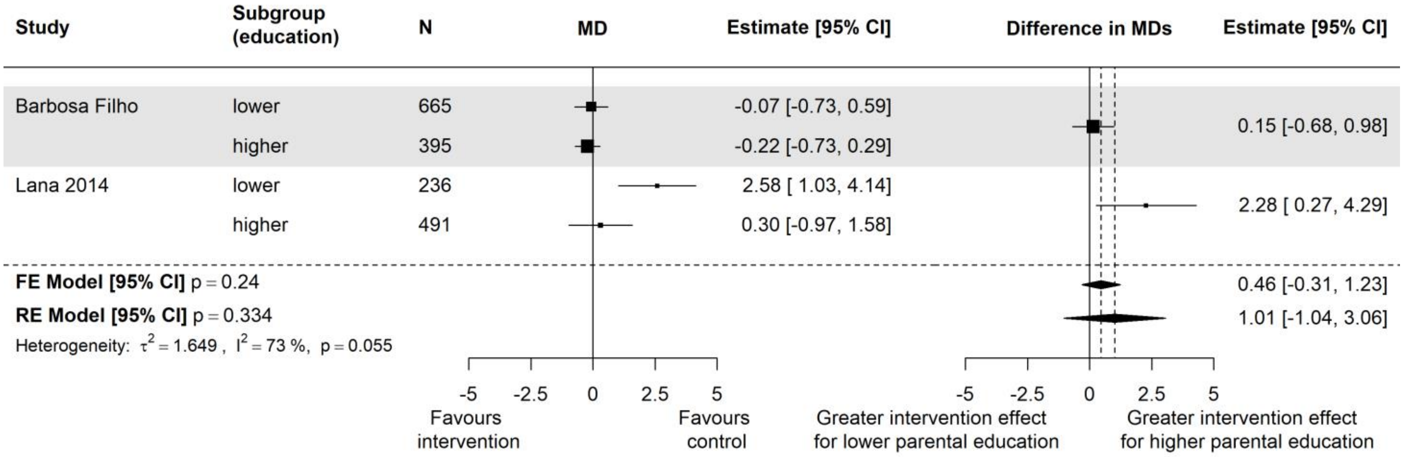

Supplementary Figure 15: Estimates of intervention effect for separate subgroups (left) and differences in intervention effect between subgroups (interactions; right) for factor **(parental) education** and outcome **BMI** in the **older age group** (12-18 years)

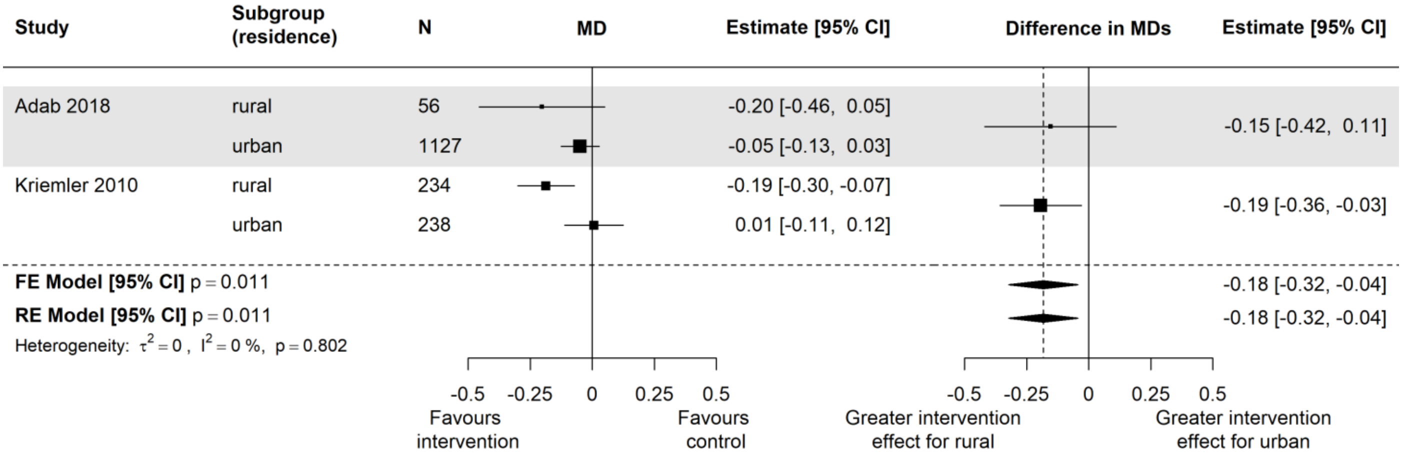

Supplementary Figure 16: Estimates of intervention effect for separate subgroups (left) and differences in intervention effect between subgroups (interactions; right) for factor **place of residence** and outcome **zBMI** in the **younger age group** (5-11 years)

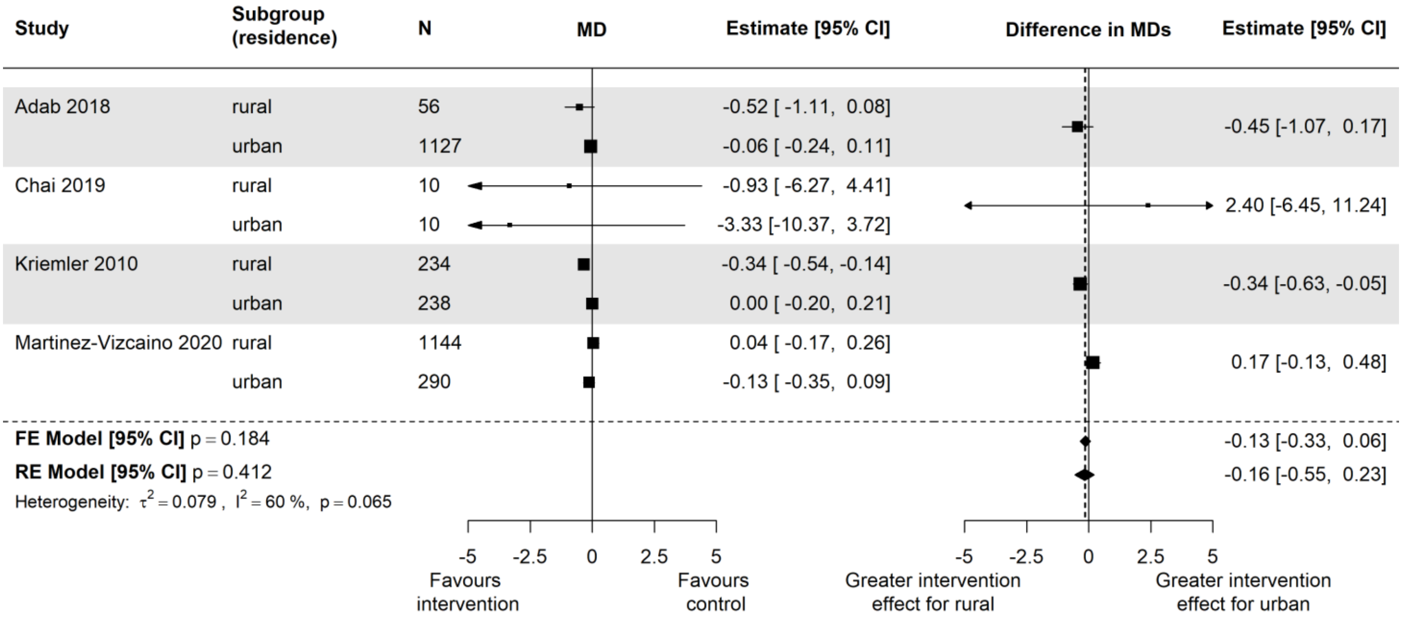

Supplementary Figure 17: Estimates of intervention effect for separate subgroups (left) and differences in intervention effect between subgroups (interactions; right) for factor **place of residence** and outcome **BMI** in the **younger age group** (5-11 years)

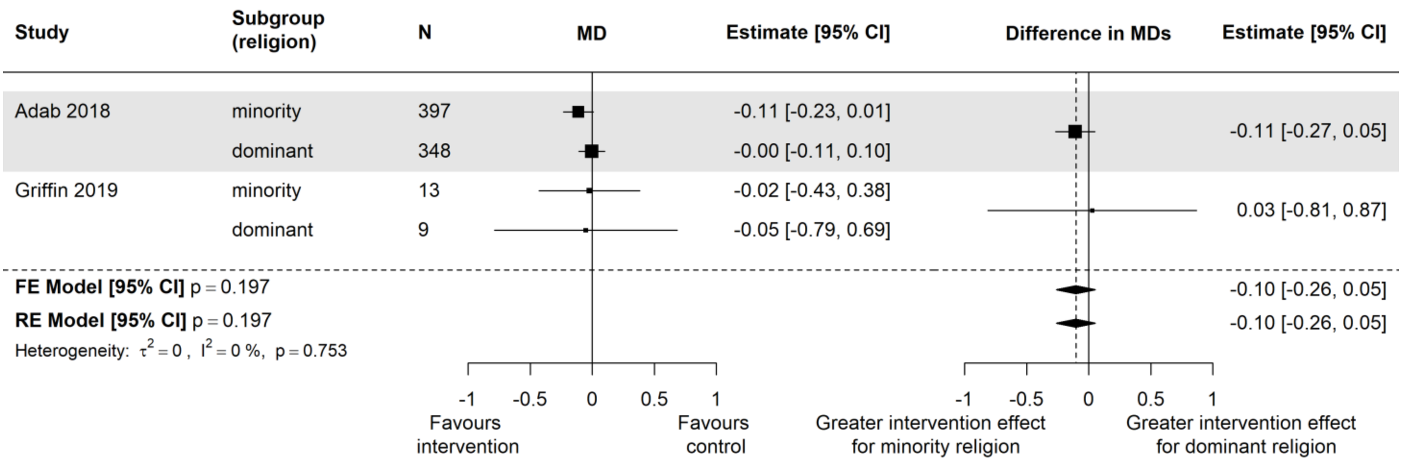

Supplementary Figure 18: Estimates of intervention effect for separate subgroups (left) and differences in intervention effect between subgroups (interactions; right) for factor **religion** and outcome **zBMI** in the **younger age group** (5-11 years)

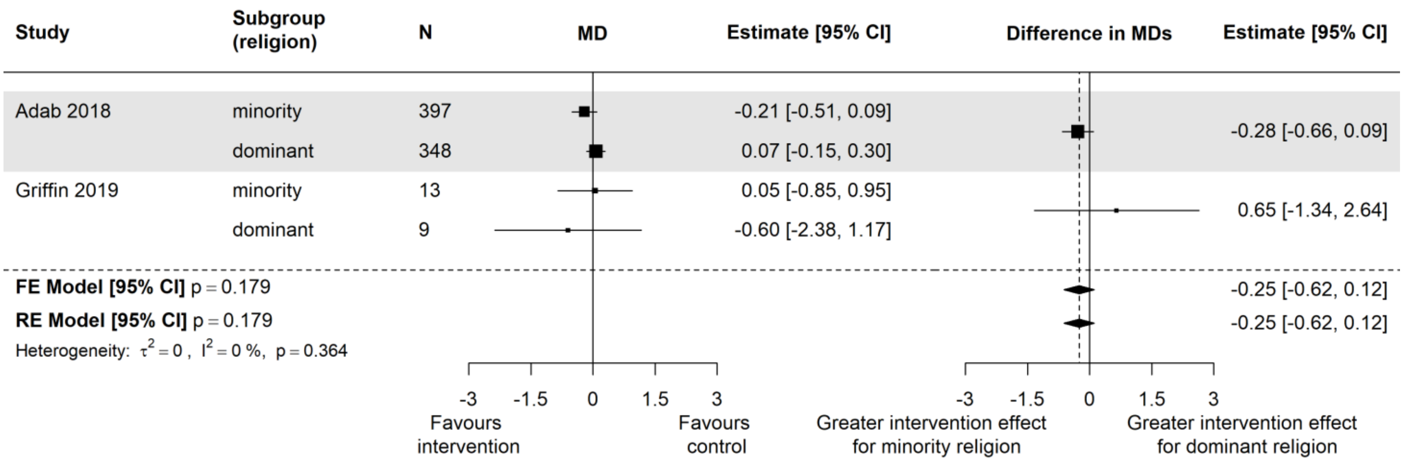

Supplementary Figure 19: Estimates of intervention effect for separate subgroups (left) and differences in intervention effect between subgroups (interactions; right) for factor **religion** and outcome **BMI** in the **younger age group** (5-11 years)

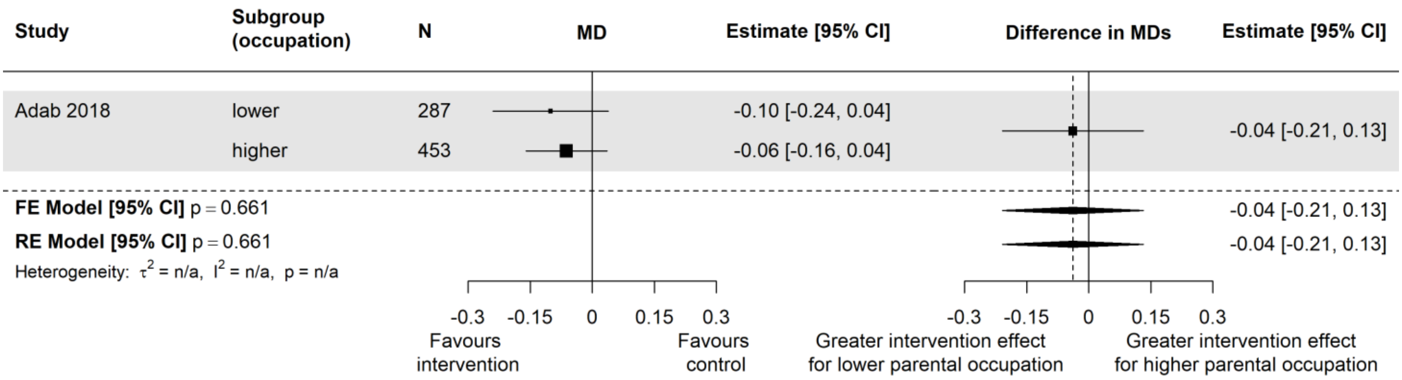

Supplementary Figure 20: Estimates of intervention effect for separate subgroups (left) and differences in intervention effect between subgroups (interactions; right) for factor **(parental) occupation** and outcome **zBMI** in the **younger age group** (5-11 years)

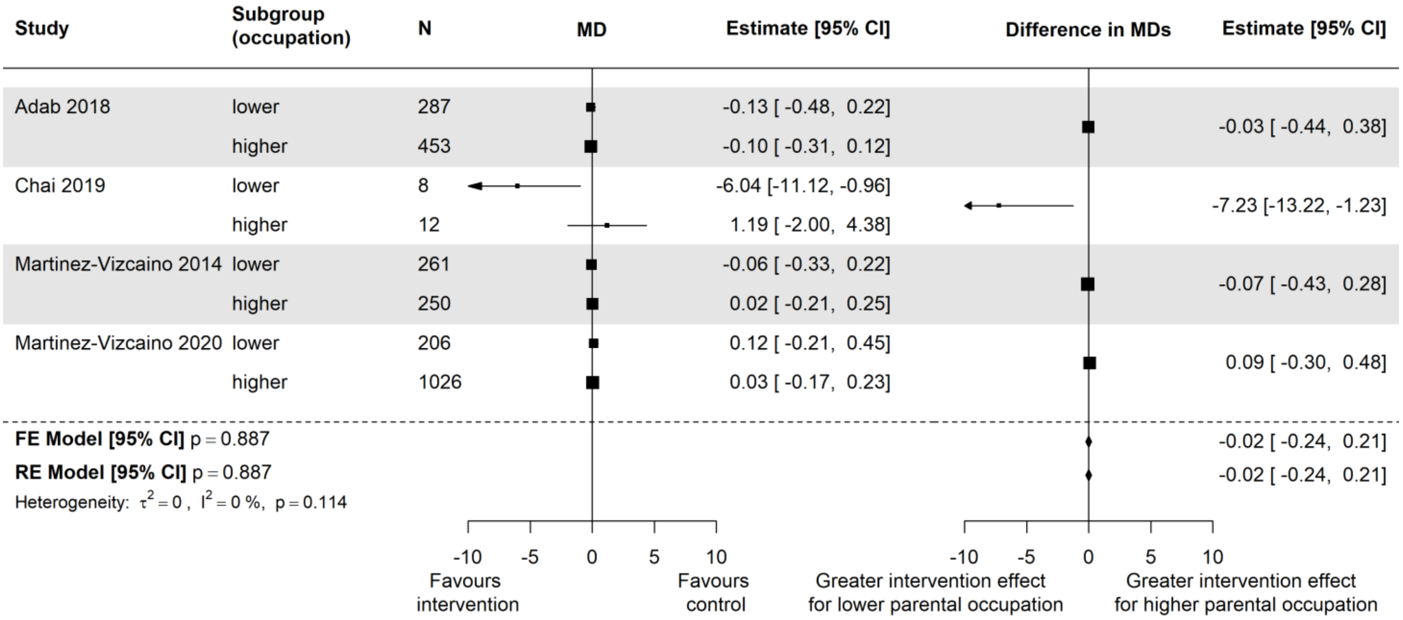

Supplementary Figure 21: Estimates of intervention effect for separate subgroups (left) and differences in intervention effect between subgroups (interactions; right) for factor **(parental) occupation** and outcome **BMI** in the **younger age group** (5-11 years)

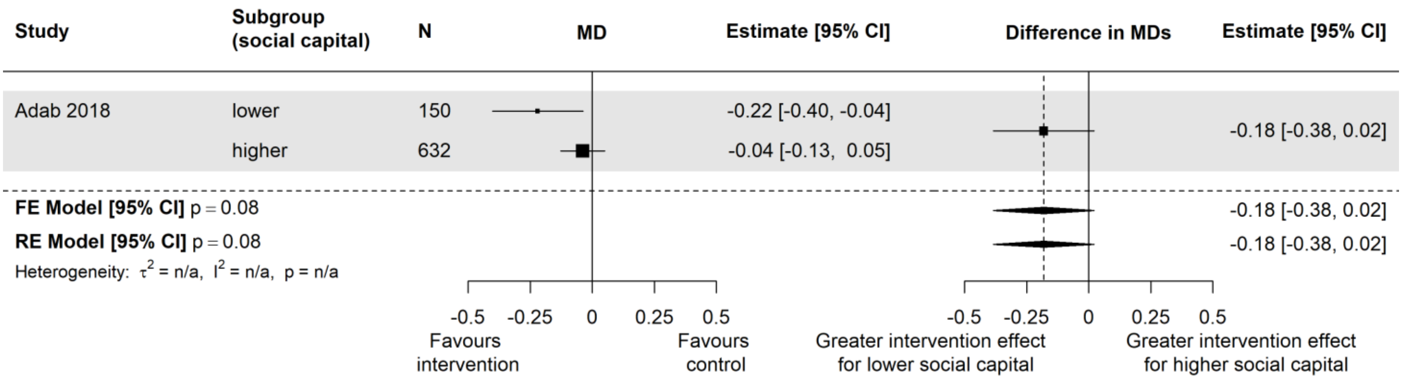

Supplementary Figure 22: Estimates of intervention effect for separate subgroups (left) and differences in intervention effect between subgroups (interactions; right) for factor **social capital** and outcome **zBMI** in the **younger age group** (5-11 years)

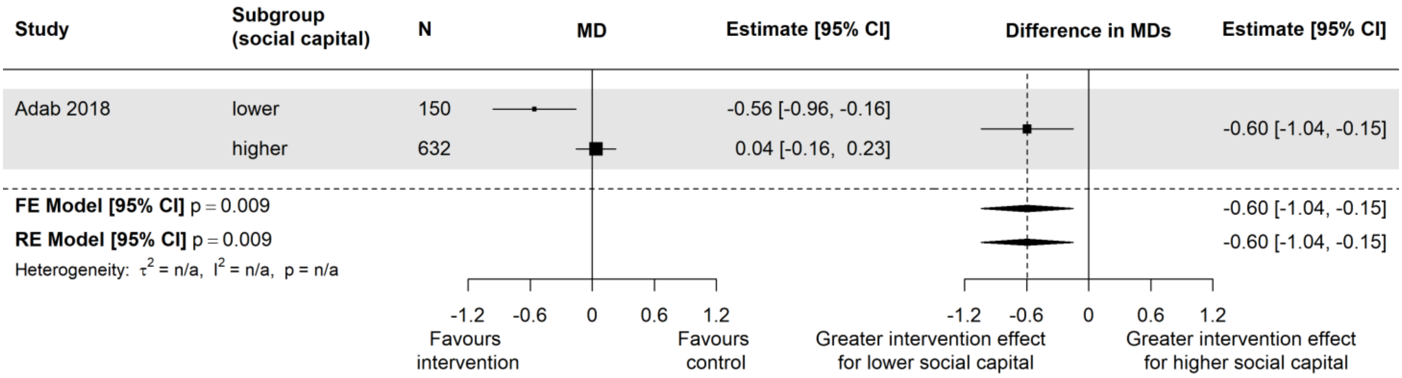

Supplementary Figure 23: Estimates of intervention effect for separate subgroups (left) and differences in intervention effect between subgroups (interactions; right) for factor **social capital** and outcome **BMI** in the **younger age group** (5-11 years)
